# Multinational attitudes towards AI in healthcare and diagnostics among hospital patients

**DOI:** 10.1101/2024.09.01.24312016

**Authors:** Felix Busch, Lena Hoffmann, Lina Xu, Longjiang Zhang, Bin Hu, Ignacio García-Juárez, Liz N Toapanta-Yanchapaxi, Natalia Gorelik, Valérie Gorelik, Gaston A Rodriguez-Granillo, Carlos Ferrarotti, Nguyen N Cuong, Chau AP Thi, Murat Tuncel, Gürsan Kaya, Sergio M Solis-Barquero, Maria C Mendez Avila, Nevena G Ivanova, Felipe C Kitamura, Karina YI Hayama, Monserrat L Puntunet Bates, Pedro Iturralde Torres, Esteban Ortiz-Prado, Juan S Izquierdo-Condoy, Gilbert M Schwarz, Jochen G Hofstaetter, Michihiro Hide, Konagi Takeda, Barbara Perić, Gašper Pilko, Hans O Thulesius, Thomas A Lindow, Israel K Kolawole, Samuel Adegboyega Olatoke, Andrzej Grzybowski, Alexandru Corlateanu, Oana-Simina Iaconi, Ting Li, Izabela Domitrz, Katarzyna Kępczyńska, Matúš Mihalčin, Lenka Fašaneková, Tomasz Zatoński, Katarzyna Fułek, András Molnár, Stefani Maihoub, Zenewton A da Silva Gama, Luca Saba, Petros Sountoulides, Marcus R Makowski, Hugo JWL Aerts, Lisa C Adams, Keno K Bressem, COMFORT consortium

## Abstract

The successful implementation of artificial intelligence (AI) in healthcare is dependent upon the acceptance of this technology by key stakeholders, particularly patients, who are the primary beneficiaries of AI-driven outcomes. This international, multicenter, cross-sectional study assessed the attitudes of hospital patients towards AI in healthcare across 43 countries. A total of 13806 patients at 74 hospitals were surveyed between February and November 2023, with 64.8% from the Global North and 35.2% from the Global South. The findings indicate a predominantly favorable general view of AI in healthcare, with 57.6% of respondents expressing a positive attitude. However, attitudes exhibited notable variation based on demographic characteristics, health status, and technological literacy. Female respondents and those with poorer health status exhibited fewer positive attitudes towards AI use in medicine. Conversely, higher levels of AI knowledge and frequent use of technology devices were associated with more positive attitudes. It is noteworthy that less than half of the participants expressed positive attitudes regarding all items pertaining to trust in AI. The lowest level of trust was observed for the accuracy of AI in providing information regarding treatment responses. Patients exhibited a strong preference for explainable AI and physician-led decision-making, even if it meant slightly compromised accuracy. This large-scale, multinational study provides a comprehensive perspective on patient attitudes towards AI in healthcare across six continents. Findings suggest a need for tailored AI implementation strategies that consider patient demographics, health status, and preferences for explainable AI and physician oversight. All study data has been made publicly available to encourage replication and further investigation.

## Introduction

Artificial Intelligence (AI) has become increasingly prevalent in various industries and public sectors, including healthcare.^1,2^ In particular, the development of large language models (LLMs) such as ChatGPT has intensified public discourse on the potential impact of AI, especially in healthcare.^3–5^

AI technologies offer promising solutions to pressing healthcare challenges, including staff shortages, high administrative costs, and economic constraints.^6^ In clinical practice, AI applications range from assisting with image-based diagnoses to personalizing treatment strategies and predicting risk factors and therapy responses.^7–9^ Beyond direct patient care, AI facilitates drug discovery and development while streamlining administrative tasks such as data extraction, curation, and report structuring.^10,11^

The economic implications are substantial, with projections suggesting that AI technologies could reduce healthcare spending in the United States by five to ten percent, potentially yielding annual savings of $200 to $360 billion.^12^ The rapid integration of AI in healthcare is further evidenced by the Food and Drug Administration’s approval of 692 AI- and machine learning-enabled medical devices through July 2023, with 478 (69.1%) approved in just the past three years.^13^

Despite this rapid growth, the benefits of AI applications to patient care are not always clear.^14^ While patient acceptance is important for the sustainable adoption of AI, patients may not always have the opportunity to consent to its use.^15,16^ To address this challenge, adopting biopsychosocial perspectives that recognize patients’ unique experiences, beliefs, and values in health maintenance can help steer AI towards patient-centered care.^15–21^ Moreover, fostering patient trust in AI is vital, as it may positively influence adherence to AI-assisted care and related health outcomes, as demonstrated in conditions such as diabetes management.^22,23^

Exploring patient perspectives can, therefore, be highly beneficial in ensuring the successful integration of AI in healthcare. Patients whose health is directly affected by AI – either through improved treatment and diagnosis or by potential consequences of immature AI – may hold views that diverge significantly from those of clinicians. However, a notable knowledge gap exists regarding patient attitudes, particularly on a large, international scale. Existing studies are limited to data from one or at most two countries, failing to capture the likely variations in patient attitudes across different sociodemographic contexts.^24–31^

To address these challenges, we conducted the first large-scale, international, multicenter survey of hospital patients to determine 1) patients’ trust, concerns, and preferences towards AI in healthcare and diagnostics and 2) factors that influence patient attitudes. By focusing on the voices of patients from diverse global contexts, including from the Global North and South, this study aims to provide a comprehensive, global perspective on patient attitudes towards AI in healthcare, thereby contributing to the development of patient-centered AI applications.

## Methods

### Study Design

This multicenter, international, cross-sectional study was conducted in accordance with the Strengthening the Reporting of Observational studies in Epidemiology (STROBE) statement (see eTable 13) and American Association for Public Opinion Research best practices for survey research. Ethical approval was obtained from the Charité – Universitätsmedizin Berlin (EA4/213/22), which served as the lead institution, and from all other participating hospitals according to their institutional policies (see eTable 14). Given the unsupervised and anonymous design of the instrument, informed consent was waived to preserve participant anonymity.

### Setting and Participants

The survey was administered to a non-probability convenience sample at 74 COMFORT network hospitals across 43 countries. Local staff disseminated the surveys, which were also displayed in prominent areas such as waiting rooms, from February 1, 2023, to November 1, 2023. We targeted radiology departments as the primary site for the survey because of the high turnover of patients with a wide range of conditions. Participants could submit their responses through drop boxes or directly to staff. Collected data from all participating sites were then centrally analyzed at Charité Berlin. The sample size was determined using Cochran’s formula. Assuming a 50% response distribution (which was chosen because it allows for the most conservative estimate), a 95% confidence level, and a five percent margin of error, we determined a minimum sample size of 385 respondents. Inclusion criteria were patients aged 18 years or older who attended a participating department during the study period, agreed to participate in the survey voluntarily, and were able to complete the questionnaire independently in one of 26 local languages (Azerbaijani, Bahasa Indonesia, Bulgarian, Croatian, Czech, Dutch, English, French, German, Greek, Hindi, Hungarian, Italian, Japanese, Macedonian, Malayalam, Mandarin, Polish, Portuguese, Romanian, Slovenian, Spanish, Swedish, Thai, Turkish, Vietnamese). Patients who did not complete any items or only items capturing variables for sample stratification were excluded.

### Survey Development and Design

To inform the survey construct for our measures on patient attitudes towards the use of AI in healthcare and diagnostics, we followed the systematic review by Young et al., synthesizing 23 qualitative, quantitative, or mixed-method original articles on patient and public attitudes toward clinical AI.^24^ In addition, a sample of ten voluntary patients who visited the Department of Radiology at Charité – Universitätsmedizin Berlin in January 2023 were interviewed to explore how patients understand and conceptualize the construct, starting with an unprompted discussion followed by focused questions on our measures. Based on the systematic review and semi-structured interviews, a multidisciplinary expert panel from the COMFORT consortium, including patient representatives, radiologists, urologists, medical faculty members and educators, AI researchers and developers, and biomedical ethicists and statisticians from seven countries, developed a 26-item survey. The comprehensibility and overall length of our instrument were evaluated in cognitive interviews with ten patients at the Department of Radiology, Charité Berlin, followed by a pilot study to test the internal reliability, consistency, and unidimensionality.

The pilot study group consisted of 100 patients visiting the Department of Radiology, Charité – University Medicine Berlin in January 2023. Psychometric validation of the questionnaire was performed using “R” version 4.2.2, including the packages “tidyverse” (1.3.2), “lavaan” (0.6-13), and “psych” (2.2.9).^32–34^ Baseline characteristics of the pilot study group are displayed in eTable 15.

The internal consistency of the questionnaire was assessed using Cronbach’s alpha. The following scales with six items each were evaluated: “Trust in AI,” “AI and Diagnosis,” and “Preferences and Concerns Towards AI.” The “Trust in AI” scale demonstrated excellent internal consistency (*α* = .94), while the “AI and Diagnosis” and “Preferences and Concerns Towards AI” scales showed good consistency (*α* = .80 and *α* = .86, respectively). The Kaiser-Meyer-Olkin (KMO) measure and Bartlett’s test of sphericity further validated the appropriateness of the data for factor analysis. The KMO measure was .93, indicating that sampling was adequate and the data were suitable for factor analysis. Bartlett’s Test of Sphericity yielded a significant result (*P* < .001), confirming that the variables were sufficiently correlated for factor analysis. Confirmatory factor analysis was used to assess the construct validity of the questionnaire. Model fit indices indicated a reasonable fit to the data, with a Comparative Fit Index of 0.956 and a Tucker-Lewis Index of 0.949, both above the recommended threshold of 0.9. The Root Mean Square Error of Approximation was .066, and the Standardized Root Mean Square Residual was .055, indicating a good model fit. Factor loadings were significant, indicating strong relationships between items and their respective latent constructs.

### Variables

The instrument consisted of three dimensions: “Trust in AI,” “AI and Diagnosis,” and “Preferences and Concerns Towards AI,” each with six items, complemented by a general data section with eight items (self-reported gender, age, highest educational level, weekly use of technological devices, health status, AI knowledge, and general attitudes towards AI in medicine and healthcare). We have chosen to collect data on gender rather than sex to allow for a more inclusive data collection that recognizes diverse individuals and reflects social identities, roles, and experiences that are not captured by biological characteristics.

“Trust in AI” measured confidence in AI improving healthcare, trust in AI providing information about health, diagnosis, response to treatment and making vital decisions, and agreement with the use of AI depending on disease severity using four- and five-point Likert scale items. “AI and Diagnosis” assessed attitudes towards AI analyzing X-rays and cancer, its role as a second opinion for physicians, trade-offs in diagnostic accuracy, and diagnostic preferences if AI and physicians would have equal accuracy using four- and five-point Likert scale and two multinomial items. “Preferences and Concerns Towards AI” assessed attitudes on the use of AI in healthcare facilities, preference for visiting such facilities, and concerns about the impact of AI on cost, data security, physician-patient interaction, and replacement of human physicians using four- and five-point Likert scale items.

### Statistical Analysis

Statistical analyses were conducted using ’R’ version 4.3.1, employing the packages ’tidyverse’ (2.0.0), ’ordinal’ (2023.12-4), and ’lme4’ (1.1-35-1) for data manipulation and modeling.^32,35,36^ Survey results were summarized using frequencies and percentages for the total cohort, the Global North and Global South based on the definitions of the United Nations Finance Center for South-South Cooperation, and by continent according to the location of the hospital visited using the United Nations Geoscheme.^37,38^ To assess differences between patient groups (categorized by gender, age, highest educational level, number of technical devices used weekly, AI knowledge, and health status), we employed cumulative link mixed models (CLMMs) and binary mixed-effects models. CLMMs were used for ordinal response items to account for the ordered nature of the responses while considering both the grouping factors as fixed effects and the collection site as a random effect to control for site-specific clustering. For questions with categorical outcomes, binomial logistic regression models were fitted, utilizing a one-vs-rest strategy. Cases with missing data were excluded from the respective analyses. Adjusted *P* values were calculated using a Bonferroni correction to address the issue of multiple comparisons. An adjusted *P* value of <.05 was considered statistically significant.

## Data Availability Statement

The full dataset and data dictionary are publicly available under CC-BY 4.0 international license at figshare: https://doi.org/10.6084/m9.figshare.24964488

## Code Availability Statement

The code for all statistical analyses is publicly available at: https://github.com/kbressem/ai-survey

## Ethics and Inclusion Statement

This study focused on capturing and comparing a wide range of hospital patient attitudes towards AI worldwide, using a non-probability convenience sample of patients from network hospitals of the COMFORT project, funded by the European Union’s Horizon Europe program and led by Charité Berlin. Each site obtained Institutional Review Board approval to conduct the survey at their institution in accordance with local guidelines. At least one principal investigator from each site was involved in the design and implementation of the survey, including substantial contributions to survey development, planning, translation (if applicable), data collection, interpretation, and validation of results. The questionnaires were completely anonymous. If participants added personal identifiers, such as names and contact information, these questionnaires were discarded to prevent the identification of individuals. In addition, all sites agreed that the original questionnaires would never be published or shared with unauthorized individuals or organizations. All participating sites had full access to the survey data and agreed that the aggregated electronic results would be made publicly available under the international CC-BY 4.0 license with publication. All consortium members have critically reviewed the manuscript, agreed to the final version for publication, and take responsibility for the accuracy and integrity of the work. Where available, we have discussed local and regional research relevant to our findings, although, at the time of manuscript preparation, all previous studies capturing patient attitudes towards AI were based in the Global North or China.^24–31^

## Results

### Participants

A total of 13955 surveys were collected, of which 1.1% (n=149/13955) were excluded from analysis due to no response to any item (0.1%, n=12/13955) or only to items in the general data section (1%, n=137/13955); see characteristics of excluded patients in eTable 1. Of the 13806 patients included, most surveys were collected in radiology departments (51.3%, n=7081/13806), followed by gastroenterology (7.9%, n=1098/13806), cardiology (5.4%, n=743/13806), and 21 other specialty departments (35.4%, n=4884/13806). Most patients (64.8%, n=8951/13806) visited hospitals in the Global North. Europe accounted for 41.7% (n=5764/13806) of patients, followed by Asia (25.2%, n=3473/13806), North America (16.5%, n=2284/13806), South America (9.7%, n=1336/13806), Africa (5.3%, n=728/13806), and Oceania (1.6%, n=221/13806). Figure 1 illustrates the geographical distribution of participating institutions. A detailed list of participating institutions and corresponding departments and patient numbers can be found in eTable 2.

**Fig. 1.**
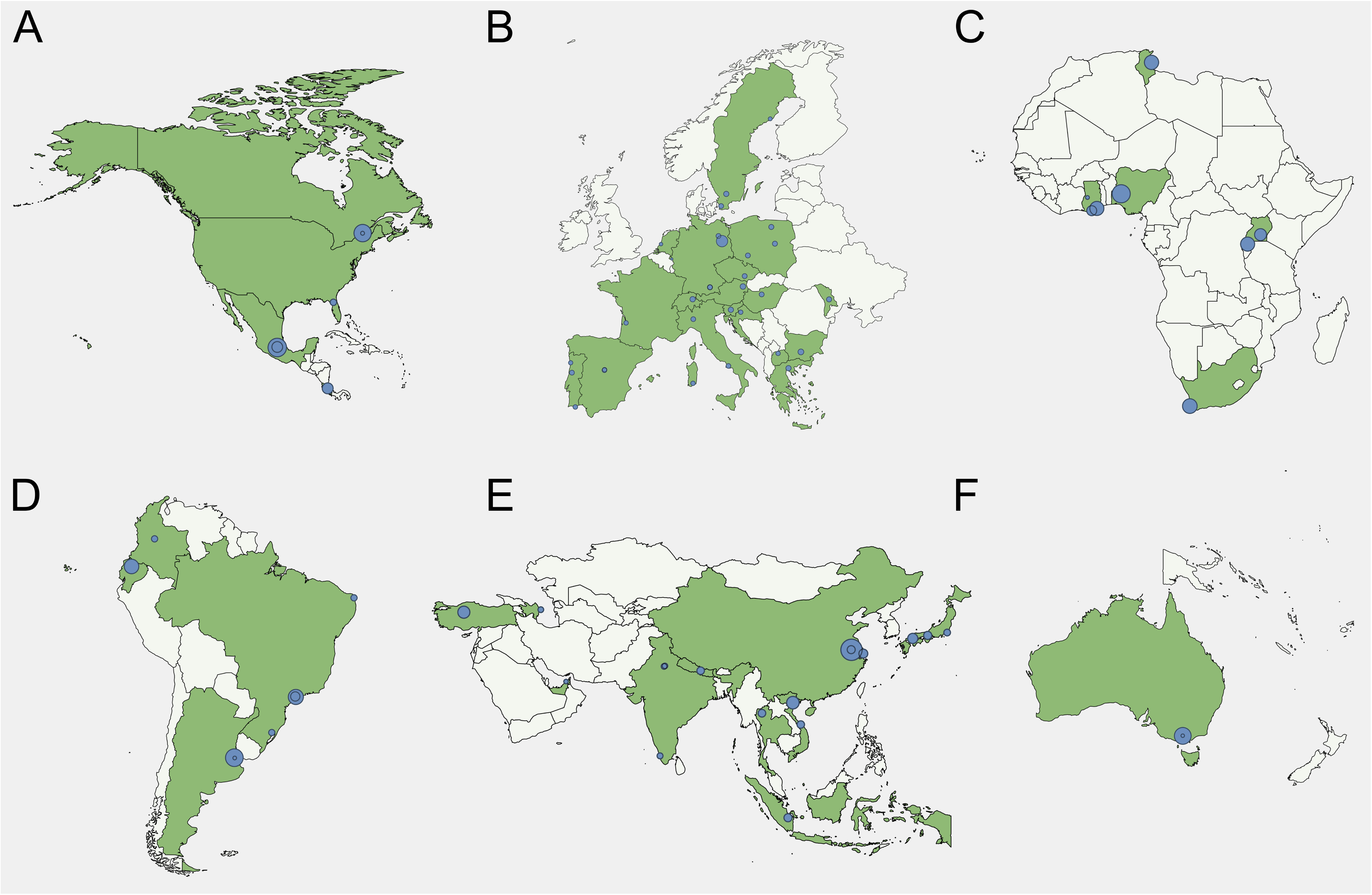
Geographical distribution of participating institutions (blue dots) on a world map. Legend: The size of the blue dots refers to the proportion of respondents per institution relative to the total number of respondents. Countries with at least one participating institution are highlighted in dark green. A, North America; B, Europe; C, Africa; D, South America; E, Asia; F, Australia.

Sociodemographic profiles and technology literacy, health status, and AI knowledge are shown in Table 1 for the overall study population and the region-specific subgroups. Gantt diagrams depicting the responses for each survey item for the total study cohort can be viewed in Figure 2. Regional breakdowns for each item response are presented in Table 2.

**Fig. 2.**
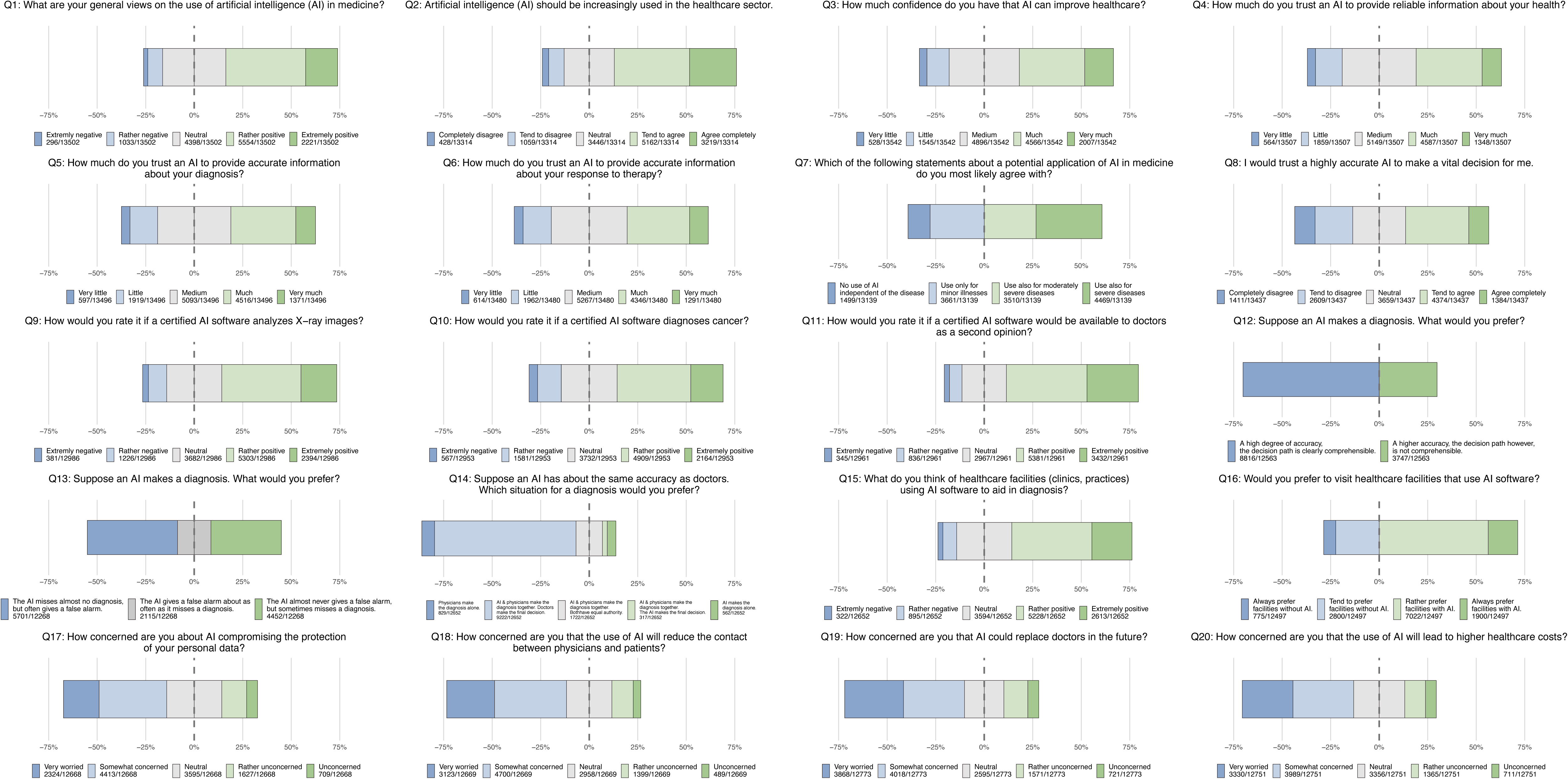
Gantt diagrams depicting the results for each item for the total study cohort. Legend: Colors represent the response options indicated below each Gantt chart, including the corresponding numerator and denominator.

**Table 1.**
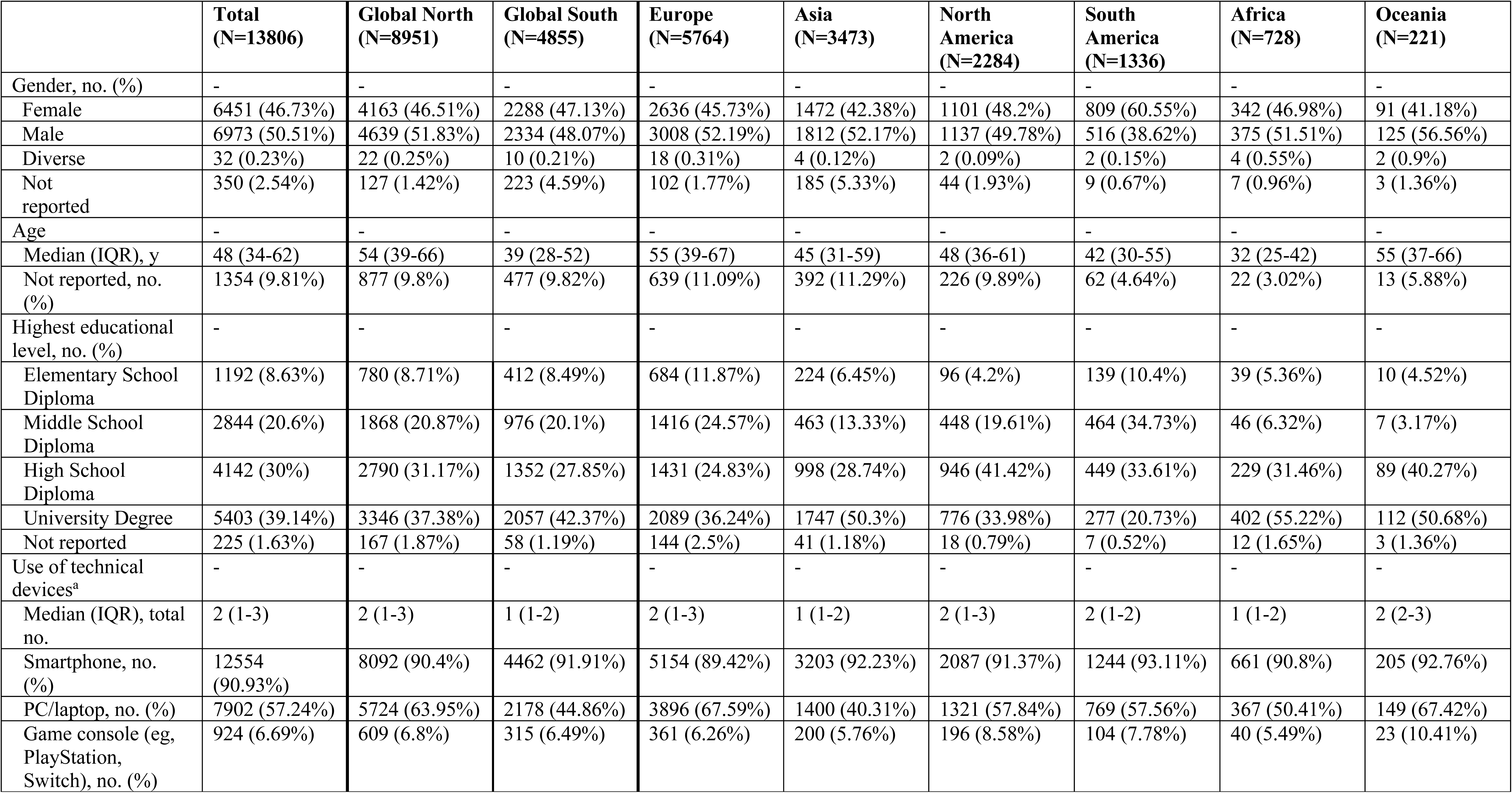
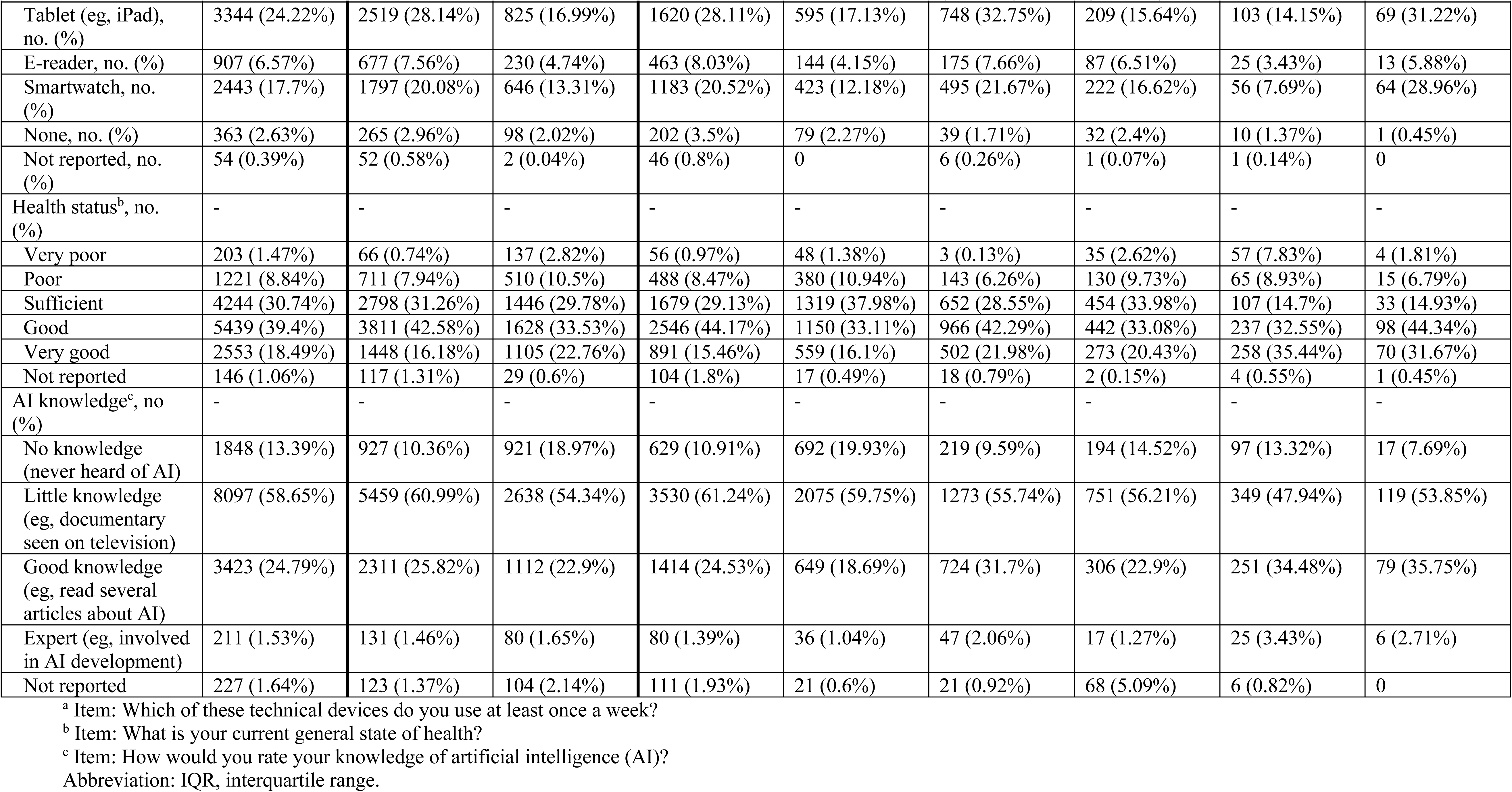
Sociodemographic data and scores on technological literacy, health status, and AI knowledge items for the total study population and regional breakdowns by Global North/South and continents.

**Table 2.** Absolute survey results and regional breakdowns for each item.

|  | **Total**<br>**(N=13806)** | **Global North**<br>**(N=8951)** | **Global South**<br>**(N=4855)** | **Europe**<br>**(N=5764)** | **Asia**<br>**(N=3473)** | **North**<br>**America**<br>**(N=2284)** | **South**<br>**America**<br>**(N=1336)** | **Africa**<br>**(N=728)** | **Oceania**<br>**(N=221)** |
| --- | --- | --- | --- | --- | --- | --- | --- | --- | --- |
| Q1, no. (%) | 13502<br>(97.8%) | 8787<br>(98.17%) | 4715<br>(97.12%) | 5617<br>(97.45%) | 3427<br>(98.68%) | 2260<br>(98.95%) | 1261<br>(94.39%) | 721 (99.04%) | 216 (97.74%) |
| Extremely negative | 296 (2.19%) | 137 (1.56%) | 159 (3.37%) | 86 (1.53%) | 78 (2.28%) | 33 (1.46%) | 31 (2.46%) | 63 (8.74%) | 5 (2.31%) |
| Rather negative | 1033 (7.65%) | 657 (7.48%) | 376 (7.97%) | 448 (7.98%) | 246 (7.18%) | 142 (6.28%) | 108 (8.56%) | 68 (9.43%) | 21 (9.27%) |
| Neutral | 4398<br>(32.57%) | 3019<br>(34.36%) | 1379<br>(29.25%) | 1980<br>(35.25%) | 987 (28.8%) | 663 (29.34%) | 457 (36.24%) | 225 (31.21%) | 86 (39.81%) |
| Rather positive | 5554<br>(41.13%) | 3787 (43.1%) | 1767<br>(37.48%) | 2389<br>(42.53%) | 1384<br>(40.39%) | 1023<br>(45.27%) | 458 (36.32%) | 218 (30.24%) | 82 (37.96%) |
| Extremely positive | 2221<br>(16.45%) | 1187<br>(13.51%) | 1034<br>(21.93%) | 714 (12.71%) | 732 (21.36%) | 399 (17.65%) | 207 (16.42%) | 147 (20.39%) | 22 (10.19%) |
| Q2, no. (%) | 13314<br>(96.44%) | 8627<br>(96.38%) | 4687<br>(96.54%) | 5477<br>(95.02%) | 3416<br>(98.36%) | 2167<br>(94.88%) | 1319<br>(98.73%) | 717 (98.49%) | 218 (98.64%) |
| Completely disagree | 428 (3.21%) | 207 (2.4%) | 221 (4.72%) | 118 (2.15%) | 112 (3.28%) | 55 (2.54%) | 73 (5.53%) | 61 (8.51%) | 9 (4.13%) |
| Tend to disagree | 1059 (7.95%) | 645 (7.48%) | 414 (8.83%) | 459 (8.38%) | 243 (7.11%) | 119 (5.49%) | 129 (9.78%) | 85 (11.85%) | 24 (11.01%) |
| Neutral | 3446<br>(25.88%) | 2480<br>(28.75%) | 966 (20.61%) | 1668<br>(30.45%) | 804 (23.54%) | 495 (22.84%) | 248 (18.8%) | 154 (21.48%) | 77 (35.32%) |
| Tend to agree | 5162<br>(38.77%) | 3482<br>(40.36%) | 1680<br>(35.84%) | 2200<br>(40.17%) | 1303<br>(38.14%) | 835 (38.53%) | 532 (40.33%) | 209 (29.15%) | 83 (38.07%) |
| Agree completely | 3219<br>(24.18%) | 1813<br>(21.02%) | 1406 (30%) | 1032<br>(18.84%) | 954 (27.93%) | 663 (30.6%) | 337 (25.55%) | 208 (29.01%) | 25 (11.47%) |
| Q3, no. (%) | 13542<br>(98.09%) | 8753<br>(97.79%) | 4789<br>(98.64%) | 5593<br>(97.03%) | 3423<br>(98.56%) | 2252 (98.6%) | 1326<br>(99.25%) | 727 (99.86%) | 221 (100%) |
| Very little | 528 (3.9%) | 323 (3.69%) | 205 (4.28%) | 202 (3.61%) | 115 (3.36%) | 75 (3.33%) | 47 (3.54%) | 76 (10.45%) | 13 (5.88%) |
| Little | 1545<br>(11.41%) | 902 (10.31%) | 643 (13.43%) | 577 (10.32%) | 374 (10.93%) | 228 (10.12%) | 232 (17.5%) | 106 (14.58%) | 28 (12.67%) |
| Medium | 4896<br>(36.15%) | 3346<br>(38.23%) | 1550<br>(32.37%) | 2200<br>(39.33%) | 1277<br>(37.31%) | 682 (30.28%) | 469 (35.37%) | 186 (25.58%) | 82 (37.1%) |
| Much | 4566<br>(33.72%) | 3104<br>(35.46%) | 1462<br>(30.53%) | 1993<br>(35.63%) | 987 (28.83%) | 895 (39.74%) | 422 (31.83%) | 213 (29.3%) | 56 (25.34%) |
| Very much | 2007<br>(14.82%) | 1078<br>(12.32%) | 929 (19.4%) | 621 (11.1%) | 670 (19.57%) | 372 (16.52%) | 156 (11.76%) | 146 (20.08%) | 42 (19%) |
| Q4, no. (%) | 13507<br>(97.83%) | 8730<br>(97.53%) | 4777<br>(98.39%) | 5573<br>(96.69%) | 3420<br>(98.47%) | 2245<br>(98.29%) | 1323<br>(99.03%) | 726 (99.73%) | 220 (99.55%) |

**Table 2. Absolute survey results and regional breakdowns for each item.**
|  | **Total**<br>**(N=13806)** | **Global North**<br>**(N=8951)** | **Global South**<br>**(N=4855)** | **Europe**<br>**(N=5764)** | **Asia**<br>**(N=3473)** | **North**<br>**America**<br>**(N=2284)** | **South**<br>**America**<br>**(N=1336)** | **Africa**<br>**(N=728)** | **Oceania**<br>**(N=221)** |
| --- | --- | --- | --- | --- | --- | --- | --- | --- | --- |
| Very little | 564 (4.18%) | 359 (4.11%) | 205 (4.29%) | 219 (3.93%) | 97 (2.84%) | 98 (4.37%) | 58 (4.38%) | 74 (10.19%) | 18 (8.18%) |
| Little | 1859<br>(13.76%) | 1083<br>(12.41%) | 776 (16.24%) | 719 (12.9%) | 478 (13.98%) | 275 (12.25%) | 249 (18.82%) | 104 (14.33%) | 34 (15.45%) |
| Medium | 5149<br>(38.12%) | 3569<br>(40.88%) | 1580<br>(33.08%) | 2455<br>(44.05%) | 1158<br>(33.86%) | 721 (32.12%) | 501 (37.87%) | 227 (31.27%) | 87 (39.55%) |
| Much | 4587<br>(33.96%) | 3075<br>(35.22%) | 1512<br>(31.65%) | 1804<br>(32.37%) | 1196<br>(34.97%) | 906 (40.36%) | 399 (30.16%) | 218 (30.03%) | 64 (29.09%) |
| Very much | 1348 (9.98%) | 644 (7.38%) | 704 (14.74%) | 376 (6.75%) | 491 (14.36%) | 245 (10.91%) | 116 (8.77%) | 103 (14.19%) | 17 (7.73%) |
| Q5, no. (%) | 13496<br>(97.75%) | 8716<br>(97.37%) | 4780<br>(98.46%) | 5561<br>(96.48%) | 3426<br>(98.65%) | 2242<br>(98.16%) | 1322<br>(98.95%) | 726 (99.73%) | 219 (99.1%) |
| Very little | 597 (4.42%) | 401 (4.6%) | 196 (4.1%) | 236 (4.24%) | 106 (3.09%) | 112 (5%) | 61 (4.61%) | 60 (8.26%) | 22 (10.05%) |
| Little | 1919<br>(14.22%) | 1110<br>(12.74%) | 809 (16.92%) | 750 (13.49%) | 473 (13.81%) | 282 (12.58%) | 253 (19.14%) | 134 (18.46%) | 27 (12.33%) |
| Medium | 5093<br>(37.74%) | 3514<br>(40.32%) | 1579<br>(33.03%) | 2395<br>(43.07%) | 1156<br>(33.74%) | 746 (33.27%) | 483 (36.54%) | 225 (30.99%) | 88 (40.18%) |
| Much | 4516<br>(33.46%) | 3027<br>(34.73%) | 1489<br>(31.15%) | 1796 (32.3%) | 1216<br>(35.49%) | 846 (37.73%) | 392 (29.65%) | 206 (28.37%) | 60 (27.4%) |
| Very much | 1371<br>(10.16%) | 664 (7.62%) | 707 (14.79%) | 384 (6.91%) | 475 (13.86%) | 256 (11.42%) | 133 (10.06%) | 101 (13.91%) | 22 (10.05%) |
| Q6, no. (%) | 13480<br>(97.64%) | 8704<br>(97.24%) | 4776<br>(98.37%) | 5551 (96.3%) | 3423<br>(98.56%) | 2238<br>(97.99%) | 1322<br>(98.95%) | 726 (99.73%) | 220 (99.55%) |
| Very little | 614 (4.55%) | 424 (4.87%) | 190 (3.98%) | 261 (4.7%) | 105 (3.07%) | 113 (5.05%) | 50 (3.78%) | 63 (8.68%) | 22 (10%) |
| Little | 1962<br>(14.55%) | 1183<br>(13.59%) | 779 (16.31%) | 777 (14%) | 459 (13.41%) | 308 (13.76%) | 252 (19.06%) | 123 (16.94%) | 43 (19.55%) |
| Medium | 5267<br>(39.07%) | 3601<br>(41.37%) | 1666<br>(34.88%) | 2493<br>(44.91%) | 1209<br>(35.32%) | 723 (32.31%) | 522 (39.49%) | 243 (33.47%) | 77 (35%) |
| Much | 4346<br>(32.24%) | 2894<br>(33.25%) | 1452 (30.4%) | 1675<br>(30.17%) | 1187<br>(34.68%) | 853 (38.11%) | 381 (28.82%) | 191 (26.31%) | 59 (26.82%) |
| Very much | 1291 (9.58%) | 602 (6.92%) | 689 (14.43%) | 345 (6.22%) | 463 (13.53%) | 241 (10.77%) | 117 (8.85%) | 106 (14.6%) | 19 (8.64%) |
| Q7, no. (%) | 13139<br>(95.17%) | 8439<br>(94.28%) | 4700<br>(96.81%) | 5361<br>(93.01%) | 3381<br>(97.35%) | 2179 (95.4%) | 1298<br>(97.16%) | 711 (97.66%) | 209 (94.57%) |
| No use of AI<br>independent of the<br>disease. | 1499<br>(11.41%) | 876 (10.38%) | 623 (13.26%) | 546 (10.18%) | 388 (11.48%) | 237 (10.88%) | 174 (13.41%) | 131 (18.42%) | 23 (11%) |

**Table 2. Absolute survey results and regional breakdowns for each item.**
|  | **Total**<br>**(N=13806)** | **Global North**<br>**(N=8951)** | **Global South**<br>**(N=4855)** | **Europe**<br>**(N=5764)** | **Asia**<br>**(N=3473)** | **North**<br>**America**<br>**(N=2284)** | **South**<br>**America**<br>**(N=1336)** | **Africa**<br>**(N=728)** | **Oceania**<br>**(N=221)** |
| --- | --- | --- | --- | --- | --- | --- | --- | --- | --- |
| Use only for minor illnesses (eg, cold). | 3661<br>(27.86%) | 2101 (24.9%) | 1560<br>(33.19%) | 1366<br>(25.48%) | 1094<br>(32.36%) | 509 (23.36%) | 444 (34.21%) | 198 (27.85%) | 50 (23.92%) |
| Use also for moderately severe diseases (eg, appendicitis). | 3510<br>(26.71%) | 2336<br>(27.68%) | 1174<br>(24.98%) | 1444<br>(26.94%) | 947 (28.01%) | 643 (29.51%) | 247 (19.03%) | 168 (23.63%) | 61 (29.19%) |
| Use also for severe diseases (eg, cancer, traffic accidents). | 4469<br>(34.01%) | 3126<br>(37.04%) | 1343<br>(28.57%) | 2005 (37.4%) | 952 (28.16%) | 790 (36.26%) | 433 (33.36%) | 214 (30.1%) | 75 (35.89%) |
| Q8, no. (%) | 13437<br>(97.33%) | 8668<br>(96.84%) | 4769<br>(98.23%) | 5522 (95.8%) | 3423<br>(98.56%) | 2225<br>(97.42%) | 1323<br>(99.03%) | 725 (99.59%) | 219 (99.1%) |
| Completely disagree | 1411 (10.5%) | 984 (11.35%) | 427 (8.95%) | 677 (12.26%) | 123 (3.59%) | 236 (10.61%) | 258 (19.5%) | 80 (11.03%) | 37 (16.89%) |
| Tend to disagree | 2609<br>(19.42%) | 1829 (21.1%) | 780 (16.36%) | 1235<br>(22.37%) | 443 (12.94%) | 475 (21.35%) | 281 (21.24%) | 136 (18.76%) | 39 (17.81%) |
| Neutral | 3659<br>(27.23%) | 2502<br>(28.86%) | 1157<br>(24.26%) | 1630<br>(29.52%) | 988 (28.86%) | 511 (22.97%) | 267 (20.18%) | 201 (27.72%) | 62 (28.31%) |
| Tend to agree | 4374<br>(32.55%) | 2716<br>(31.33%) | 1658<br>(34.77%) | 1590<br>(28.79%) | 1339<br>(39.12%) | 776 (34.88%) | 388 (29.33%) | 212 (29.24%) | 69 (31.51%) |
| Agree completely | 1384 (10.3%) | 637 (7.35%) | 747 (15.66%) | 390 (7.06%) | 530 (15.48%) | 227 (10.2%) | 129 (9.75%) | 96 (13.24%) | 12 (5.48%) |
| Q9, no. (%) | 12986<br>(94.06%) | 8243<br>(92.09%) | 4743<br>(97.69%) | 5134<br>(89.07%) | 3401<br>(97.93%) | 2196<br>(96.15%) | 1317<br>(98.58%) | 722 (99.18%) | 216 (97.74%) |
| Extremely negative | 381 (2.93%) | 191 (2.32%) | 190 (4.01%) | 105 (2.05%) | 105 (3.09%) | 54 (2.46%) | 52 (3.95%) | 57 (7.89%) | 8 (3.7%) |
| Rather negative | 1226 (9.44%) | 685 (8.31%) | 541 (11.41%) | 470 (9.15%) | 268 (7.88%) | 162 (7.38%) | 237 (18%) | 69 (9.56%) | 20 (9.26%) |
| Neutral | 3682<br>(28.35%) | 2318<br>(28.12%) | 1364<br>(28.76%) | 1475<br>(28.73%) | 1093<br>(32.14%) | 528 (24.04%) | 336 (25.51%) | 190 (26.32%) | 60 (27.78%) |
| Rather positive | 5303<br>(40.84%) | 3726 (45.2%) | 1577<br>(33.25%) | 2292<br>(44.64%) | 1223<br>(35.96%) | 970 (44.17%) | 475 (36.07%) | 242 (33.52%) | 101 (46.76%) |
| Extremely positive | 2394<br>(18.44%) | 1323<br>(16.05%) | 1071<br>(22.58%) | 792 (15.43%) | 712 (20.94%) | 482 (21.95%) | 217 (16.48%) | 164 (22.71%) | 27 (12.5%) |
| Q10, no. (%) | 12953<br>(93.82%) | 8219<br>(91.82%) | 4734<br>(97.51%) | 5110<br>(88.65%) | 3399<br>(97.87%) | 2192<br>(95.97%) | 1314<br>(98.35%) | 721 (99.04%) | 217 (98.19%) |
| Extremely negative | 567 (4.38%) | 323 (3.93%) | 244 (5.15%) | 211 (4.13%) | 118 (3.47%) | 79 (3.6%) | 84 (6.39%) | 63 (8.74%) | 12 (5.53%) |
| Rather negative | 1581<br>(12.21%) | 868 (10.56%) | 713 (15.06%) | 597 (11.68%) | 433 (12.74%) | 194 (8.85%) | 248 (18.87%) | 71 (9.85%) | 38 (17.51%) |

**Table 2. Absolute survey results and regional breakdowns for each item.**
|  | **Total**<br>**(N=13806)** | **Global North**<br>**(N=8951)** | **Global South**<br>**(N=4855)** | **Europe**<br>**(N=5764)** | **Asia**<br>**(N=3473)** | **North**<br>**America**<br>**(N=2284)** | **South**<br>**America**<br>**(N=1336)** | **Africa**<br>**(N=728)** | **Oceania**<br>**(N=221)** |
| --- | --- | --- | --- | --- | --- | --- | --- | --- | --- |
| Neutral | 3732<br>(28.81%) | 2354<br>(28.64%) | 1378<br>(29.11%) | 1477 (28.9%) | 1037<br>(30.51%) | 584 (26.64%) | 351 (26.71%) | 222 (30.79%) | 61 (28.11%) |
| Rather positive | 4909 (37.9%) | 3431<br>(41.74%) | 1478<br>(31.22%) | 2091<br>(40.92%) | 1190<br>(35.01%) | 895 (40.83%) | 438 (33.33%) | 219 (30.37%) | 76 (35.02%) |
| Extremely positive | 2164<br>(16.71%) | 1243<br>(15.12%) | 921 (19.46%) | 734 (14.36%) | 621 (18.27%) | 440 (20.07%) | 193 (14.69%) | 146 (20.25%) | 30 (13.82%) |
| Q11, no. (%) | 12961<br>(93.88%) | 8232<br>(91.97%) | 4729 (97.4%) | 5122<br>(89.86%) | 3395<br>(97.75%) | 2188 (95.8%) | 1316 (98.5%) | 723 (99.31%) | 217 (98.19%) |
| Extremely negative | 345 (2.66%) | 161 (1.96%) | 184 (3.89%) | 82 (1.6%) | 110 (3.24%) | 50 (2.29%) | 43 (3.27%) | 51 (7.05%) | 9 (4.15%) |
| Rather negative | 836 (6.45%) | 392 (4.76%) | 444 (9.39%) | 255 (4.98%) | 280 (8.25%) | 79 (3.61%) | 112 (8.51%) | 93 (12.86%) | 17 (7.83%) |
| Neutral | 2967<br>(22.89%) | 1716<br>(20.85%) | 1251<br>(26.45%) | 1073<br>(20.95%) | 1006<br>(29.63%) | 391 (17.87%) | 295 (22.42%) | 161 (22.27%) | 41 (18.89%) |
| Rather positive | 5381<br>(41.52%) | 3672<br>(44.61%) | 1709<br>(36.14%) | 2238<br>(43.69%) | 1246 (36.7%) | 976 (44.61%) | 594 (45.14%) | 232 (32.09%) | 95 (43.78%) |
| Extremely positive | 3432<br>(26.48%) | 2291<br>(27.83%) | 1141<br>(24.13%) | 1474<br>(28.78%) | 753 (22.18%) | 692 (31.63%) | 272 (20.67%) | 186 (25.73%) | 55 (25.35%) |
| Q12, no. (%) | 12563 (91%) | 7931 (88.6%) | 4632<br>(95.41%) | 4887<br>(84.78%) | 3331<br>(95.91%) | 2123<br>(92.95%) | 1294<br>(96.86%) | 721 (99.04%) | 207 (93.67%) |
| A high degree of accuracy, the decision path is clearly comprehensible (explainable AI). | 8816<br>(70.17%) | 5599 (70.6%) | 3217<br>(69.45%) | 3304<br>(67.61%) | 2091<br>(62.77%) | 1704<br>(80.26%) | 1045<br>(80.76%) | 520 (72.12%) | 152 (73.43%) |
| A higher accuracy, the decision path however, is not comprehensible. | 3747<br>(29.83%) | 2332 (29.4%) | 1415<br>(30.55%) | 1583<br>(32.39%) | 1240<br>(37.23%) | 419 (19.74%) | 249 (19.24%) | 201 (27.88%) | 55 (26.57%) |
| Q13, no. (%) | 12268<br>(88.86%) | 7671 (85.7%) | 4597<br>(94.69%) | 4680<br>(81.19%) | 3301<br>(95.05%) | 2086<br>(91.33%) | 1285<br>(96.18%) | 710 (97.53%) | 206 (93.21%) |
| The AI misses almost no diagnosis, but often gives a false alarm. | 5701<br>(46.47%) | 3725<br>(48.56%) | 1976<br>(42.98%) | 2224<br>(47.52%) | 1323<br>(40.08%) | 1025<br>(49.14%) | 673 (52.37%) | 336 (47.32%) | 120 (58.25%) |

**Table 2. Absolute survey results and regional breakdowns for each item.**
|  | **Total**<br>**(N=13806)** | **Global North**<br>**(N=8951)** | **Global South**<br>**(N=4855)** | **Europe**<br>**(N=5764)** | **Asia**<br>**(N=3473)** | **North**<br>**America**<br>**(N=2284)** | **South**<br>**America**<br>**(N=1336)** | **Africa**<br>**(N=728)** | **Oceania**<br>**(N=221)** |
| --- | --- | --- | --- | --- | --- | --- | --- | --- | --- |
| The AI almost never gives a false alarm, but sometimes misses a diagnosis. | 4452<br>(36.29%) | 2663<br>(34.72%) | 1789<br>(38.92%) | 1668<br>(35.64%) | 1296<br>(39.26%) | 732 (35.09%) | 456 (35.49%) | 243 (34.23%) | 57 (27.67%) |
| The AI gives a false alarm about as often as it misses a diagnosis. | 2115<br>(17.24%) | 1283<br>(16.73%) | 832 (18.1%) | 788 (16.84%) | 682 (20.66%) | 329 (15.77%) | 156 (12.14%) | 131 (18.45%) | 29 (14.08%) |
| Q14, no. (%) | 12652<br>(91.64%) | 8089<br>(90.37%) | 4563<br>(93.99%) | 5012<br>(86.95%) | 3305<br>(95.16%) | 2157<br>(94.44%) | 1244<br>(93.11%) | 720 (98.9%) | 214 (96.83%) |
| Physicians make the diagnosis alone. | 829 (6.55%) | 570 (7.05%) | 259 (5.68%) | 345 (6.88%) | 198 (5.99%) | 111 (5.15%) | 113 (9.08%) | 51 (7.08%) | 11 (5.14%) |
| AI and physicians make the diagnosis together. Doctors make the final decision. | 9222<br>(72.89%) | 6092<br>(75.31%) | 3130 (68.6%) | 3853<br>(76.88%) | 2212<br>(66.93%) | 1594 (73.9%) | 896 (72.03%) | 496 (68.89%) | 171 (79.91%) |
| AI and physicians make the diagnosis together. Both have equal authority. | 1722<br>(13.61%) | 1063<br>(13.14%) | 659 (14.44%) | 574 (11.45%) | 563 (17.03%) | 298 (13.82%) | 152 (12.22%) | 114 (15.83%) | 21 (9.81%) |
| AI and physicians make the diagnosis together. The AI makes the final decision. | 317 (2.51%) | 153 (1.89%) | 164 (3.59%) | 111 (2.21%) | 98 (2.97%) | 30 (1.39%) | 40 (3.22%) | 34 (4.72%) | 4 (1.87%) |
| AI makes the diagnosis alone. | 562 (4.44%) | 211 (2.61%) | 351 (7.69%) | 129 (2.57%) | 234 (7.08%) | 124 (5.75%) | 43 (3.46%) | 25 (3.47%) | 7 (3.27%) |
| Q15, no. (%) | 12652<br>(91.64%) | 8077<br>(90.24%) | 4575<br>(94.23%) | 4992<br>(86.61%) | 3244<br>(93.41%) | 2167<br>(94.88%) | 1312 (98.2%) | 723 (99.31%) | 214 (96.83%) |
| Extremely negative | 322 (2.55%) | 138 (1.71%) | 184 (4.02%) | 69 (1.38%) | 93 (2.87%) | 39 (1.8%) | 38 (2.9%) | 76 (10.51%) | 7 (3.27%) |
| Rather negative | 895 (7.07%) | 419 (5.19%) | 476 (10.4%) | 274 (5.49%) | 338 (10.42%) | 89 (4.11%) | 103 (7.85%) | 75 (10.37%) | 16 (7.48%) |
| Neutral | 3594<br>(28.41%) | 2381<br>(29.48%) | 1213<br>(26.51%) | 1555<br>(31.15%) | 890 (27.44%) | 502 (23.17%) | 434 (33.08%) | 159 (21.99%) | 54 (25.23%) |

**Table 2. Absolute survey results and regional breakdowns for each item.**
|  | **Total**<br>**(N=13806)** | **Global North**<br>**(N=8951)** | **Global South**<br>**(N=4855)** | **Europe**<br>**(N=5764)** | **Asia**<br>**(N=3473)** | **North**<br>**America**<br>**(N=2284)** | **South**<br>**America**<br>**(N=1336)** | **Africa**<br>**(N=728)** | **Oceania**<br>**(N=221)** |
| --- | --- | --- | --- | --- | --- | --- | --- | --- | --- |
| Rather positive | 5228<br>(41.32%) | 3617<br>(44.78%) | 1611<br>(35.21%) | 2184<br>(43.75%) | 1219<br>(37.58%) | 990 (45.69%) | 494 (37.65%) | 242 (33.47%) | 99 (46.26%) |
| Extremely positive | 2613<br>(20.65%) | 1522<br>(18.84%) | 1091<br>(23.85%) | 910 (18.23%) | 704 (21.7%) | 547 (25.24%) | 243 (18.52%) | 171 (23.65%) | 38 (17.76%) |
| Q16, no. (%) | 12497<br>(90.52%) | 7947<br>(88.78%) | 4523<br>(93.16%) | 4905 (85.1%) | 3214<br>(92.54%) | 2150<br>(94.13%) | 1298<br>(97.16%) | 715 (98.21%) | 215 (97.29%) |
| Always prefer<br>facilities without<br>AI. | 775 (6.2%) | 426 (5.36%) | 349 (7.72%) | 299 (6.1%) | 213 (6.63%) | 78 (3.63%) | 130 (10.02%) | 38 (5.31%) | 17 (7.91%) |
| Tend to prefer<br>facilities without<br>AI. | 2800<br>(22.41%) | 1855<br>(23.34%) | 945 (20.89%) | 1183<br>(24.12%) | 645 (20.07%) | 426 (19.81%) | 355 (27.35%) | 126 (17.62%) | 65 (30.23%) |
| Rather prefer<br>facilities with AI. | 7022<br>(56.19%) | 4750<br>(59.77%) | 2272<br>(50.23%) | 2841<br>(57.92%) | 1808<br>(56.25%) | 1287<br>(59.86%) | 656 (50.54%) | 314 (43.92%) | 116 (53.95%) |
| Always prefer<br>facilities with AI. | 1900 (15.2%) | 943 (11.87%) | 957 (21.16%) | 582 (11.87%) | 548 (17.05%) | 359 (16.7%) | 157 (12.1%) | 237 (33.15%) | 17 (7.91%) |
| Q17, no. (%) | 12668<br>(91.76%) | 8104<br>(90.54%) | 4564<br>(94.01%) | 5019<br>(87.07%) | 3239<br>(93.26%) | 2161<br>(94.61%) | 1311<br>(98.13%) | 722 (99.18%) | 216 (97.74) |
| Very worried | 2324<br>(18.35%) | 1497<br>(18.47%) | 827 (18.12%) | 718 (14.31%) | 441 (13.62%) | 703 (32.53%) | 267 (20.37%) | 152 (21.05%) | 43 (19.91%) |
| Somewhat<br>concerned | 4413<br>(34.84%) | 2830<br>(34.92%) | 1583<br>(34.68%) | 1673<br>(33.33%) | 1233<br>(38.07%) | 727 (33.64%) | 441 (33.64%) | 252 (34.9%) | 87 (40.28%) |
| Neutral | 3595<br>(28.38%) | 2220<br>(27.39%) | 1375<br>(30.13%) | 1513<br>(30.15%) | 1008<br>(31.12%) | 453 (20.96%) | 350 (26.7%) | 213 (29.5%) | 58 (26.85%) |
| Rather unconcerned | 1627<br>(12.84%) | 1115<br>(13.76%) | 512 (11.22%) | 775 (15.44%) | 394 (12.16%) | 219 (10.13%) | 151 (11.52%) | 67 (9.28%) | 21 (9.72%) |
| Unconcerned | 709 (5.6%) | 442 (5.45%) | 267 (5.85%) | 340 (6.77%) | 163 (5.03%) | 59 (2.73%) | 102 (7.78%) | 38 (5.26%) | 7 (3.24%) |
| Q18, no. (%) | 12669<br>(91.76%) | 8109<br>(90.59%) | 4560<br>(93.92%) | 5025<br>(87.18%) | 3237 (93.2%) | 2160<br>(94.59%) | 1312 (98.2%) | 719 (98.76%) | 216 (97.74%) |
| Very worried | 3123<br>(24.65%) | 2150<br>(26.51%) | 973 (21.34%) | 1206 (24%) | 516 (15.94%) | 786 (36.39%) | 385 (29.34%) | 177 (24.62%) | 53 (24.54%) |
| Somewhat<br>concerned | 4700 (37.1%) | 3174<br>(39.14%) | 1526<br>(33.46%) | 2023<br>(40.26%) | 1265<br>(39.08%) | 698 (32.31%) | 375 (28.58%) | 241 (33.52%) | 98 (45.37%) |

**Table 2. Absolute survey results and regional breakdowns for each item.**
|  | **Total**<br>**(N=13806)** | **Global North**<br>**(N=8951)** | **Global South**<br>**(N=4855)** | **Europe**<br>**(N=5764)** | **Asia**<br>**(N=3473)** | **North**<br>**America**<br>**(N=2284)** | **South**<br>**America**<br>**(N=1336)** | **Africa**<br>**(N=728)** | **Oceania**<br>**(N=221)** |
| --- | --- | --- | --- | --- | --- | --- | --- | --- | --- |
| Neutral | 2958<br>(23.35%) | 1699<br>(20.95%) | 1259<br>(27.61%) | 1044<br>(20.78%) | 882 (27.25%) | 467 (21.62%) | 338 (25.76%) | 183 (25.45%) | 44 (20.37%) |
| Rather unconcerned | 1399<br>(11.04%) | 862 (10.63%) | 537 (11.78%) | 593 (11.8%) | 403 (12.45%) | 178 (8.24%) | 133 (10.14%) | 76 (10.57%) | 16 (7.41%) |
| Unconcerned | 489 (3.86%) | 224 (2.76%) | 265 (5.81%) | 159 (3.16%) | 171 (5.28%) | 31 (1.44%) | 81 (6.17%) | 42 (5.84%) | 5 (2.31%) |
| Q19, no. (%) | 12773<br>(92.52%) | 8105<br>(90.55%) | 4668<br>(96.15%) | 5025<br>(87.18%) | 3341<br>(96.20%) | 2158<br>(94.48%) | 1311<br>(98.13%) | 722 (99.18%) | 216 (97.74%) |
| Very worried | 3868<br>(30.28%) | 2724<br>(33.61%) | 1144<br>(24.51%) | 1580<br>(31.44%) | 593 (17.75%) | 918 (42.54%) | 466 (35.55%) | 230 (31.86%) | 81 (37.5%) |
| Somewhat concerned | 4018<br>(31.46%) | 2743<br>(33.84%) | 1275<br>(27.31%) | 1717<br>(34.17%) | 1102<br>(32.98%) | 647 (29.98%) | 293 (22.35%) | 187 (25.9%) | 72 (33.33%) |
| Neutral | 2595<br>(20.32%) | 1409<br>(17.38%) | 1186<br>(25.41%) | 841 (16.74%) | 867 (25.95%) | 375 (17.38%) | 334 (25.48%) | 143 (19.81%) | 35 (16.2%) |
| Rather unconcerned | 1571 (12.3%) | 910 (11.23%) | 661 (14.16%) | 647 (12.88%) | 501 (15%) | 180 (8.34%) | 131 (9.99%) | 89 (12.33%) | 23 (10.65%) |
| Unconcerned | 721 (5.64%) | 319 (3.94%) | 402 (8.61%) | 240 (4.78%) | 278 (8.32%) | 38 (1.76%) | 87 (6.64%) | 73 (10.11%) | 5 (2.31%) |
| Q20, no. (%) | 12751<br>(92.36%) | 8091<br>(90.39%) | 4660<br>(95.98%) | 5009 (86.9%) | 3334 (96%) | 2159<br>(94.53%) | 1310<br>(98.05%) | 723 (99.31%) | 216 (97.74%) |
| Very worried | 3330<br>(26.12%) | 2151<br>(26.59%) | 1179 (25.3%) | 1093<br>(21.82%) | 676 (20.28%) | 845 (39.14%) | 421 (32.14%) | 245 (33.89%) | 50 (23.15%) |
| Somewhat concerned | 3989<br>(31.28%) | 2439<br>(30.14%) | 1550<br>(33.26%) | 1408<br>(28.11%) | 1241<br>(37.22%) | 688 (31.87%) | 362 (27.63%) | 215 (29.74%) | 75 (34.72%) |
| Neutral | 3356<br>(26.32%) | 2205<br>(27.25%) | 1151 (24.7%) | 1518<br>(30.31%) | 872 (26.15%) | 444 (20.57%) | 327 (24.96%) | 132 (18.26%) | 63 (29.17%) |
| Rather unconcerned | 1365<br>(10.71%) | 885 (10.94%) | 480 (10.3%) | 683 (13.64%) | 359 (10.77%) | 128 (5.93%) | 127 (9.69%) | 51 (7.05%) | 17 (7.87%) |
| Unconcerned | 711 (5.58%) | 411 (5.08%) | 300 (6.44%) | 307 (6.13%) | 186 (5.58%) | 54 (2.5%) | 73 (5.57%) | 80 (11.07%) | 11 (5.09%) |

Abbreviations: Q1, What are your general views on the use of artificial intelligence (AI) in medicine?; Q2, Artificial intelligence (AI) should be increasingly used in the healthcare sector.; Q3, How much confidence do you have that AI can improve healthcare?; Q4, How much do you trust an AI to provide reliable information about your health?; Q5, How much do you trust an AI to provide accurate information about your diagnosis?; Q6, How much do you trust an AI to provide accurate information about your response to therapy?; Q7, Which of the following statements about a potential application of AI in medicine do you most likely agree with?; Q8, I would trust a highly accurate AI to make a vital decision for me.; Q9, How would you rate it if a certified AI software analyzes X-ray images?; Q10, How would you rate it if a certified AI software diagnoses cancer?; Q11, How would you rate it if a certified AI software would be available to doctors as a second opinion?; Q12/Q13, Suppose an AI makes a diagnosis. What would you prefer?; Q14, Suppose an AI has about the same accuracy as doctors. Which situation for a diagnosis would you prefer?; Q15, What do you think of healthcare facilities (clinics, practices) using AI software to aid in diagnosis?; Q16, Would you prefer to visit healthcare facilities that use AI software? Q17, How concerned are you about AI compromising
the protection of your personal data?; Q18, How concerned are you that the use of AI will reduce the contact between physicians and patients?; Q19, How concerned are you that AI could replace human doctors in the future?; Q20, How concerned are you that the use of AI will lead to higher healthcare costs?.

### General attitudes towards AI

Most patients were positive about the general use of AI in medicine (question (Q) 1; 57.6%, n=7775/13502) and favored its increasing application in healthcare (Q2; 63%, n=8381/13314). Female respondents tended to be slightly less positive about the general use of AI in medicine than males, with 55.6% (n=3511/6318) having “rather positive” or “extremely positive” views on AI compared to 59.1% (n=4057/6864) of males (Q1; adjusted odds ratio (AOR): 0.84, 95% confidence interval (CI): 0.78 to 0.9; see eTables 3 and 8). Patients tended to be more dismissive towards AI if they reported worse overall health status. Of the patients with very poor health status, 26.3% (n=53/199) had “extremely negative” and 29.2% (n=58/199) “rather negative” views on AI. In comparison, only 1.3% (n=33/2538) and 5.3% (n=134/2538) of patients with very good health shared those views (see eTable 9). This trend was also reflected in the AOR, which increased with higher self-reported health status, ranging from 0.15 (95% CI: 0.11 to 0.21) for very poor to 0.71 (95% CI: 0.64 to 0.78) for good, compared with those who indicated very good health (Q1; see eTable 4).

Similar observations were made for higher AI knowledge, where 83.3% (n=175/210) of self-reported AI experts had “rather positive” or “extremely positive” views, compared to 38% (n=667/1755) of those with no AI knowledge (Q1; see eTable 10). For this question, AORs ranged from 1.75 (95% CI: 1.56 to 1.96) for little knowledge to 7.11 (95% CI: 5.19 to 9.74) for expert compared to no knowledge (Q1; see eTable 5).

Patients with a higher technological affinity or literacy, measured by the number of technology devices used weekly, also showed a higher tendency to express positive views on AI (Q1; AOR: 1.17, 95% CI: 1.13 to 1.21; see eTable 6). Age and level of education did not significantly influence general attitudes (Q1/Q2; see eTables 6 and 7). Absolute survey results for each item stratified by gender, education level, health status, AI knowledge, age, and weekly use of technological devices are presented in eTables 8-12.

### Trust in AI

Less than half of the respondents indicated a positive attitude towards the items related to trust in AI. Overall, 48.5% (n=6573/13542) of patients surveyed were confident that AI would improve healthcare (Q3), 43.9% (n=5935/13507) trusted AI to provide reliable health information (Q4), 43.6% (n=5887/13496) trusted AI to provide accurate information about their diagnosis (Q5), and 41.8% (n=5637/13480) trusted AI to provide accurate information about their response to therapy (Q6).

While the majority of female patients responded positively towards AI for all items on trust, they were slightly less favorable than male patients, reflected in the lower AORs compared to males, ranging from 0.76 (Q5 and Q6; 95% CIs: 0.71 to 0.8 and 0.71 to 0.81) to 0.8 (Q4; 95% CIs: 0.74 to 0.85; see eTable 3). For instance, in Q3, 45% (n=2862/6357) of female respondents had “much” or “very much” confidence that AI can improve healthcare, compared to 51.4% (n=3526/6861) of male respondents (see eTable 8).

For Q3 to Q6, patients with expert knowledge consistently expressed higher tendencies to answer more favorably towards AI with AORs ranging from 3.26 (Q4; 95% CI: 2.41 to 4.41) to 5.11 (Q3; 95% CI: 3.76 to 6.94; see eTable 5). For example, in Q3, 77.3% (n=163/211) of self-reported AI experts had “much” or “very much” confidence in AI improving healthcare, compared to only 35.9% (n=630/1756) of those with no AI knowledge (see eTable 10). Similarly, AORs for Q3 to Q6 increased with better self-reported health status, with the reference group of patients reporting very good health consistently demonstrating the highest AORs (see eTable 4).

Only 11.4% (n=1499/13139) of patients were against using AI regardless of the disease (Q7). In contrast, 27.9% (n=3661/13139) preferred to use AI only for minor conditions such as the common cold, 26.7% (n=3510/13139) accepted AI for moderate conditions such as appendicitis, and 34% (n=4469/13139) were open to using AI for severe conditions such as traffic accidents. Notably, 42.9% (n=5758/13437) of patients trusted a highly accurate AI to make vital health decisions on their behalf (Q8). However, this trust varied significantly in terms of both health status and AI knowledge. 50.4% (n=1273/2526) of patients with very good health trusted AI for vital decisions, compared to only 26% (n=51/196) of those with very poor health, reflected in an AOR of 0.45 (95% CI: 0.33-0.62; see eTables 4 and 9). 91.4% (n=192/210) of self-reported AI experts trusted or strongly trusted AI for vital decisions, compared to only 36% (n=628/1745) of those with no AI knowledge, with AORs ranging from 1.18 (95% CI: 1.06-1.32) for little knowledge to 2.06 (95% CI: 1.52-2.8) for expert knowledge (see eTables 5 and 10).

### Preferences towards AI applications in diagnostics and healthcare facilities

The majority of patients preferred healthcare facilities that use AI software to assist in diagnosis (Q15), with 62% (n=7841/12652) expressing a positive attitude. Similarly, most patients indicated that they would often or always prefer facilities that use AI (Q16; 71.4%, n=8922/12497). Female patients tended to be less positive than males for both items (Q15: 59.9% (n=3585/5990) versus 63.9% (n=4041/6329); Q16: 68.7% (n=4061/5912) versus 73.7% (n=4618/6263)) with AORs of 0.79 (95% CI: 0.78 to 0.79) for Q15 and 0.81 (95% CI: 0.75 to 0.88) for Q16 (see eTables 3 and 8). AI knowledge also significantly influenced these preferences, with AORs for expert knowledge of 4.77 (95% CI: 3.51 to 6.48) for Q15 and 3.18 (95% CI: 2.29 to 4.42) for Q16, compared to no knowledge. For instance, 79.7% (n=161/202) of self-reported AI experts had positive attitudes towards healthcare facilities using AI compared to 46.2% (n=747/1616) of those with no knowledge (Q15; see eTables 5 and 9). Younger age and higher technological literacy were only associated with a more positive attitude for Q15 (see eTable 6).

The use of AI was viewed positively in various medical scenarios: 59.3% (n=7697/12986) supported the use of AI for X-ray analysis (Q9), 54.6% (n=7073/12953) for cancer diagnosis (Q10), and 68% (n=8804/12961) for availability as a second opinion for physicians (Q11). Notably, 70.2% (n=8816/12563) of patients preferred explainable AI (Q12), even if this meant a trade-off in accuracy compared to black-box models. This observation was consistent across subgroups with small and mostly non-significant differences.

Regarding diagnostic accuracy (Q13), 46.5% (n=5701/12268) preferred AI with higher sensitivity compared to 36.3% (n=4452/12268) who preferred AI with higher specificity. When asked about joint diagnosis by physicians and AI when both have the same accuracy (Q14), the majority (72.9%, n=9222/12652) preferred a collaborative diagnostic approach where physicians make the final decision. Only a small proportion (4.4%, n=562/12652) supported the idea of fully autonomous AI in diagnosis, while 6.6% (n=829/12652) favored physicians making the diagnosis independently of AI.

### Concerns towards AI

Concerns about data protection were expressed by 53.2% (n=6737/12668) of patients (Q17). Even more participants were concerned about AI’s potential impact on healthcare delivery: 61.8% (n=7823/12669) feared that AI could reduce doctor-patient interaction (Q18), while 61.7% (n=7886/12773) were concerned that AI could replace human doctors (Q19). The expectation that AI will lead to increased healthcare costs was a concern for 57.4% (n=7319/12751) of patients (Q20).

Younger age was associated with lower concerns for all items (Q17 to Q20; AORs, 95% CIs ranging from 0.36, 0.29 to 0.46 (Q18) to 0.62, 0.49 to 0.79 (Q17); see eTable 6). However, absolute differences were small, with 53.3% of patients ≤48 years (n=3167/5938) vs. 53.4% (n=2984/5593) of patients >48 years expressing concerns in Q17 or 57.4% (n=3412/5940) of patients ≤48 years vs. 66.7% (n=3731/5592) of patients >48 years expressing concerns in Q18 (eTable 11).

Notably, higher self-reported AI knowledge was associated with lower concerns about the replacement of human doctors (Q19; 50.2% (n=107/205) of experts vs. 62.2% (n=1037/1668) with no knowledge, AOR expert: 1.84, 95% CI: 1.35 to 2.5) and increased healthcare costs (Q20; 51.7% of experts versus 62.6% with no knowledge, AOR expert: 1.83, 95% CI: 1.34 to 2.5; see eTables 5 and 10).

## Discussion

This multinational study represents the most extensive and comprehensive survey to date of patient attitudes toward AI in healthcare worldwide. With 13806 participants from 43 countries, our findings provide a multifaceted understanding of patients’ preferences, trust, and concerns about AI in healthcare. The results illustrate a nuanced landscape of attitudes, with most patients expressing support for the use of AI in healthcare while also articulating concerns about its implementation.

Previous studies examining patient attitudes towards AI in healthcare have been limited to individual countries or specific clinical areas. For example, positive attitudes towards the use of AI in healthcare ranged from 53% in a German tertiary referral hospital to 94% in a German radiology patient study.^26,29,31,39^ Although this overall trend is also reflected in our findings, with 57.6% of respondents expressing a generally positive view of the use of AI in healthcare, our study provides a more comprehensive and granular understanding of patient attitudes. Notably, 71.4% of patients indicated a preference for healthcare facilities that utilize AI software (Q16). Interestingly, the preference for facilities utilizing AI was higher than the percentage of patients expressing a generally positive view of AI (57.6%, Q1) or favoring an increase in AI use in healthcare (63%, Q2). One potential explanation for this discrepancy is that the use of AI is perceived as a marker of modern technology, and patients anticipate that other aspects of the hospital may also be more modern. This perception may provide a rationale for healthcare providers to allocate greater resources toward AI solutions, particularly in contexts where the private healthcare sector is prominent.

Our study also uniquely demonstrates how attitudes towards AI vary significantly based on demographic factors and health status. Young, healthy males tend to view AI most positively, while older patients and those with poorer health express more reservations. This gradient of acceptance suggests that as the likelihood of AI being applied to one’s own care increases, patients become more cautious in their outlook. Similar trends were also observed for the use of AI depending on disease severity, where patients were less likely to accept AI for more severe conditions. This finding is also supported by a recent study by Khullar et al., who found that 31% of respondents to an online survey agreed with the use of AI in cancer diagnosis, compared to 55% for chest X-rays.^29^ However, it is important to note that their study allowed respondents to agree with AI use in multiple scenarios and focused on AI potentially replacing doctors in these activities. In contrast, our study examined the acceptance of AI application across a spectrum of disease severities, regardless of whether it was supplementing or replacing human doctors. In contrast, a study by Robertson et al. involving 2675 patients showed no significant preference for AI use based on disease severity.^25^

A noteworthy finding of our study is the pronounced inclination towards explainable AI, which was observed to be independent of demographic characteristics. Approximately 70% of patients indicated a preference for AI with transparent decision-making processes, even if this entailed a slight compromise in accuracy. This preference for explainability is considerably higher than that reported in a previous US study, which found that only 42% of patients felt uncomfortable with highly accurate AI diagnoses that lacked explainability.^29^ Our study demonstrates a global desire for transparency in AI-driven healthcare decisions, which has significant implications for the development and implementation of AI in medical settings.

Moreover, our results reinforce the importance of maintaining human oversight in AI-assisted healthcare. Despite the ongoing debate about the use of autonomous AI in healthcare,^40–42^ our findings indicate that the majority of patients prefer physicians to retain control when utilizing AI in clinical settings. Notably, only 4% of patients preferred fully autonomous AI. Supporting these findings, previous studies have reported that 67% to 96% of patients would prefer physician-led diagnoses to AI recommendations.^26,31,43,44^ On the other hand, the preference for physicians to make diagnoses without the assistance of AI was expressed by only 6.6% of patients, suggesting a substantial endorsement of AI among the survey participants.

Despite the generally favorable views on AI, we also observed multiple concerns among respondents. Over half voiced apprehensions about data security, reduced doctor-patient interaction, and potential increases in healthcare costs. These findings underscore the need for a balanced approach to AI implementation in healthcare, one that addresses patient concerns while leveraging the potential benefits of AI technology.

This study has limitations. The non-probability convenience sampling likely resulted in low response rates and may have introduced selection and noncoverage bias, affecting data representativeness. Despite these issues, the sampling method enabled the collection of diverse patient attitudes across various countries and healthcare settings. The uncertain selection probabilities and unsupervised survey administration may limit the robustness of inferences. To address site-specific clustering and stratification variations, we used mixed models for subgroup analysis. While not fully generalizable to all hospital populations, the findings offer valuable insights into multinational patient attitudes towards AI in healthcare and can inform future research. The authors encourage replication and extension, particularly in underrepresented populations, and have made study materials available to support this.

In conclusion, this global survey, which includes patients across six continents, provides the most comprehensive snapshot of patient attitudes toward AI in healthcare to date. Our findings reveal a nuanced landscape. While patients generally favor AI-equipped healthcare facilities and recognize AI’s potential, they strongly prefer explainable AI systems and physician-led decision-making. Furthermore, attitudes vary significantly based on demographics and health status. These insights underscore the critical need for healthcare providers and AI developers to prioritize transparency, maintain human oversight, and tailor AI implementation to patient characteristics.

## Supporting information

eTables 1-15

## Data Availability

The full dataset and data dictionary are publicly available under CC-BY 4.0 international license at figshare.

https://doi.org/10.6084/m9.figshare.24964488

## Acknowledgments

The authors thank: at the College of Pharmacy and Health Sciences, Ajman University, United Arab Emirates, Ms Dania S Rammal, Ms Aya Mutasem Baradie, and Ms Farrah E Elsubeihi for supporting the data collection; at the Department of Medical Oncology, Dr BRA Institute Rotary Cancer Hospital, Ms Ritika Arora (supporting the data collection), Ms Swetambri Sharma (supporting the translation), Ms Mamta Kumari (supporting the translation), Ms Ayushi Bansal (supporting the translation), and Ms Vasudha V (supporting the translation) for supporting the data collection and questionnaire translation into Hindi; at the Diagnostic and Imaging Services Department, SSD Infectious Diseases 3 - Ultrasound, Fondazione IRCCS Policlinico San Matteo, Pavia, Italy, Ms Nadia Locatelli for supporting the data collection; at the Department of Radiology, Alfred Health, Melbourne, Victoria, Australia, Mr Dominic Buensalido and Ms Christina Tasiopoulos for supporting the data collection and Mr Adil Zia for supporting the ethics submission and data collection; at the Department of Radiology, Jinling Hospital, Affiliated Hospital of Medical School, Nanjing University, Nanjing, China, Mr Jian Zhong for supporting the data collection; at the Department of Gastroenterology, Lakeshore Hospital, Kochi, India, Mr Binimol Shaji for supporting the data collection; at the Department of Radiology, Ramón y Cajal University Hospital, Madrid, Spain, Dr Ana M Ayala Carbonero and Dr María J Carrillo Guivernau for supporting the data collection; at the Department of Imaging, A.C.Camargo Cancer Center, São Paulo, Brazil, Dr Paula NVP Barbosa for supporting the data collection; at the Department of Radiology, Kantonsspital Glarus, Glarus, Switzerland, Mr Christian Weiser for supporting the data collection; all hospital and administration staff at each location who made this study possible. This research is funded by the European Union (COMFORT (Computational Models FOR patienT stratification in urologic cancers – Creating robust and trustworthy multimodal AI for health care), project number: 101079894, authors involved: Petros Sountoulides, Renato Cuocolo, Virginia Dignum, Guillermo de Velasco, Alessa Hering, Lili Jiang, George Kolostoumpis, Alexander Loeser, principal investigator: Keno K Bressem, sponsors’ website: https://www.comfort-ai.eu). Views and opinions expressed are, however, those of the authors only and do not necessarily reflect those of the European Union. The European Union cannot be held responsible for them. The funding had no role in the study design, data collection and analysis, manuscript preparation, or decision to publish.

## Author Contributions

Conceptualization: Felix Busch, Lena Hoffmann, Lina Xu, Zenewton A da Silva Gama, Petros Sountoulides, Lisa C Adams, Keno K Bressem, Renato Cuocolo, Virginia Dignum, Guillermo de Velasco, Alessa Hering, Lili Jiang, George Kolostoumpis, Alexander Loeser, Marcus R Makowski, Daniel Truhn; Project administration: Felix Busch, Lena Hoffmann, Lisa C Adams, Keno K Bressem; Resources: All COMFORT consortium members; Software: Felix Busch, Lena Hoffmann, Lisa C Adams, Keno K Bressem; Data curation: Felix Busch, Lena Hoffmann, Lisa C Adams, Keno K Bressem; Formal analysis: Felix Busch, Lena Hoffmann, Lisa C Adams, Keno K Bressem; Funding acquisition: Felix Busch, Lena Hoffmann, Lina Xu, Petros Sountoulides, Lisa C Adams, Keno K Bressem, Renato Cuocolo, Virginia Dignum, Guillermo de Velasco, Alessa Hering, Lili Jiang, George Kolostoumpis, Alexander Loeser, Marcus R Makowski; Investigation: All COMFORT consortium members; Methodology: Felix Busch, Lena Hoffmann, Lisa C Adams, Keno K Bressem; Supervision: Felix Busch, Lisa C Adams, Keno K Bressem; Validation: All COMFORT consortium members; Visualization: Felix Busch, Esteban Ortiz-Prado, Keno K Bressem; Writing – original draft preparation: Felix Busch, Lisa C Adams, Keno K Bressem; Writing – review & editing: All COMFORT consortium members.

### Table of COMFORT consortium members (ordered alphabetically by surname)

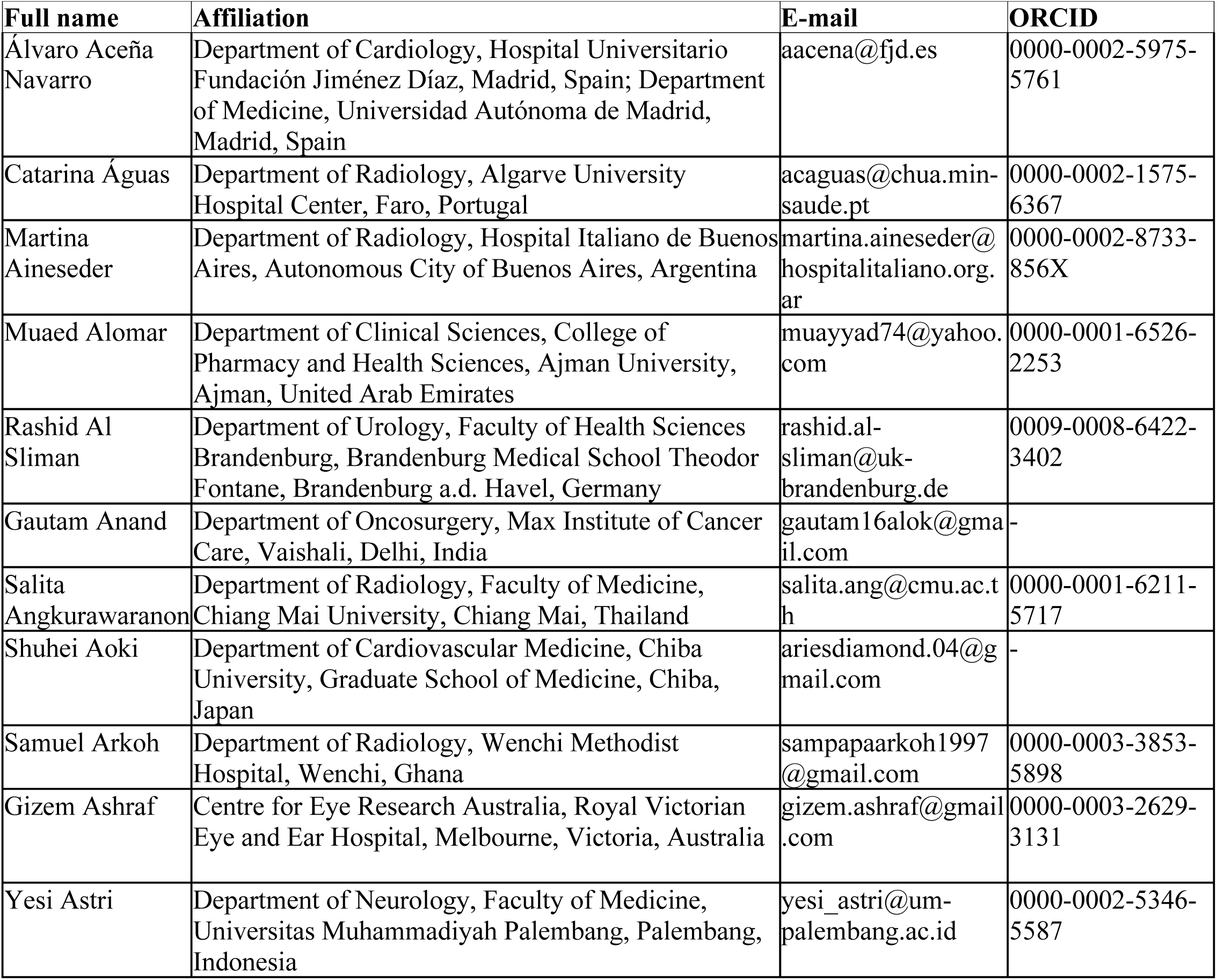
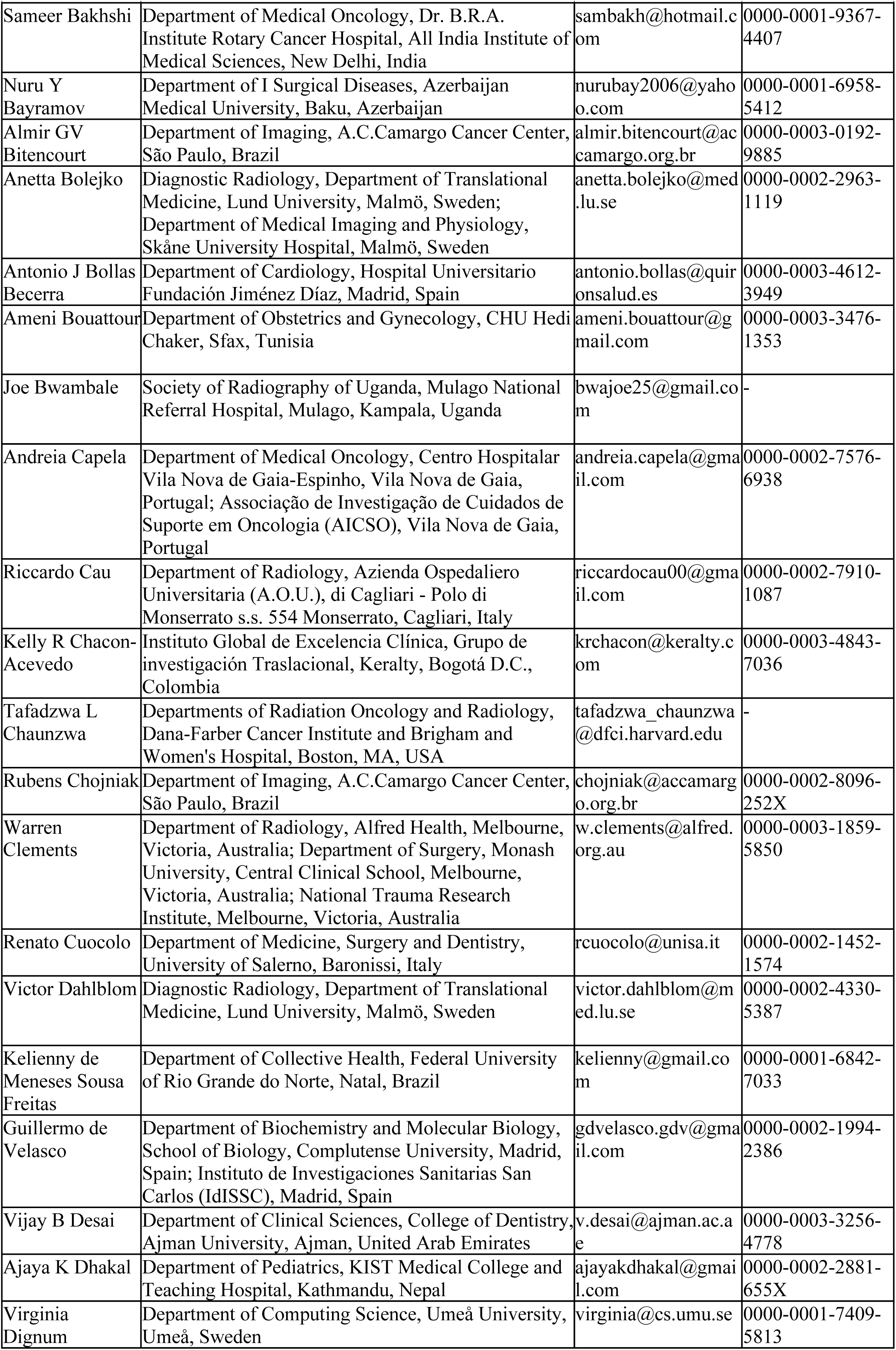
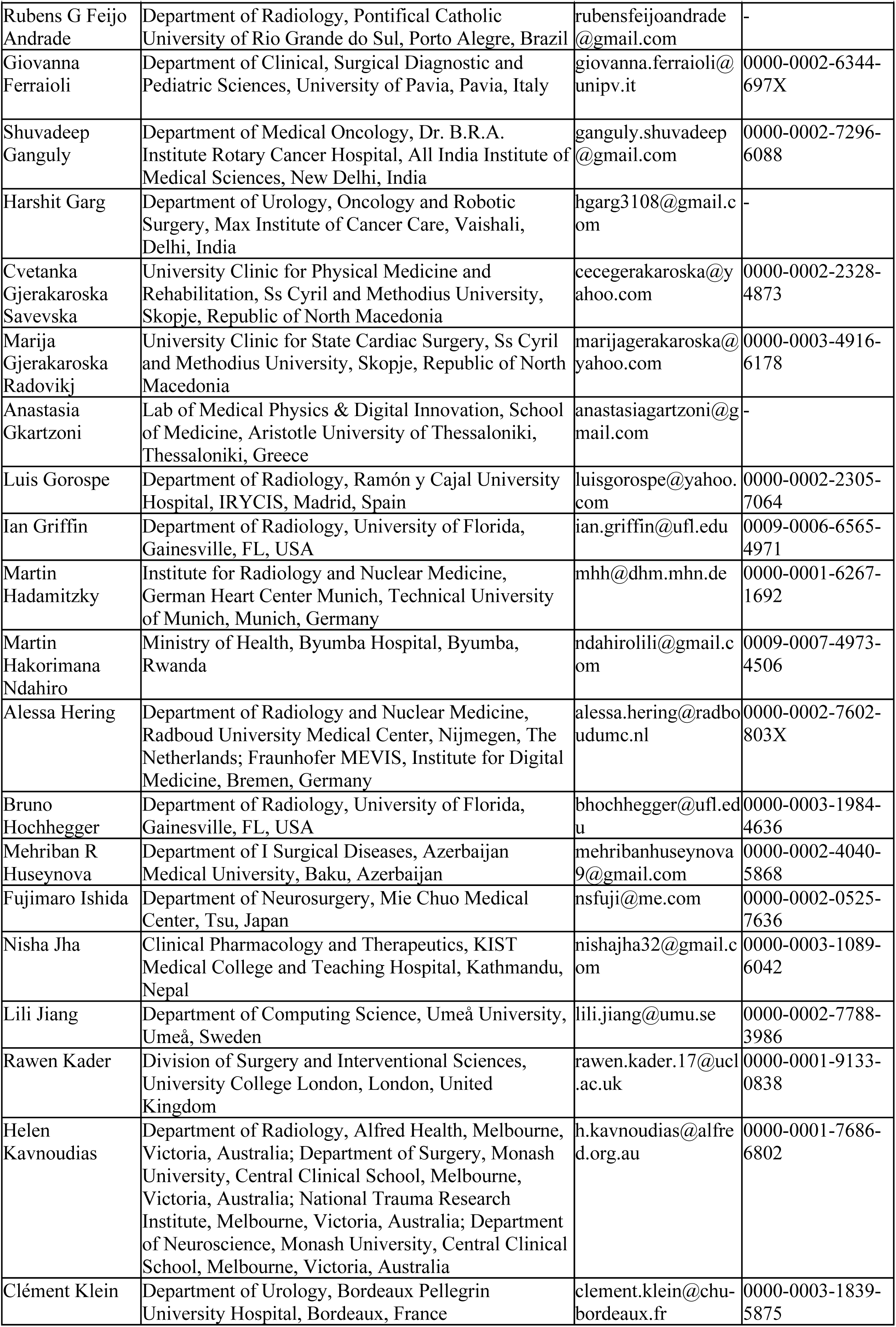
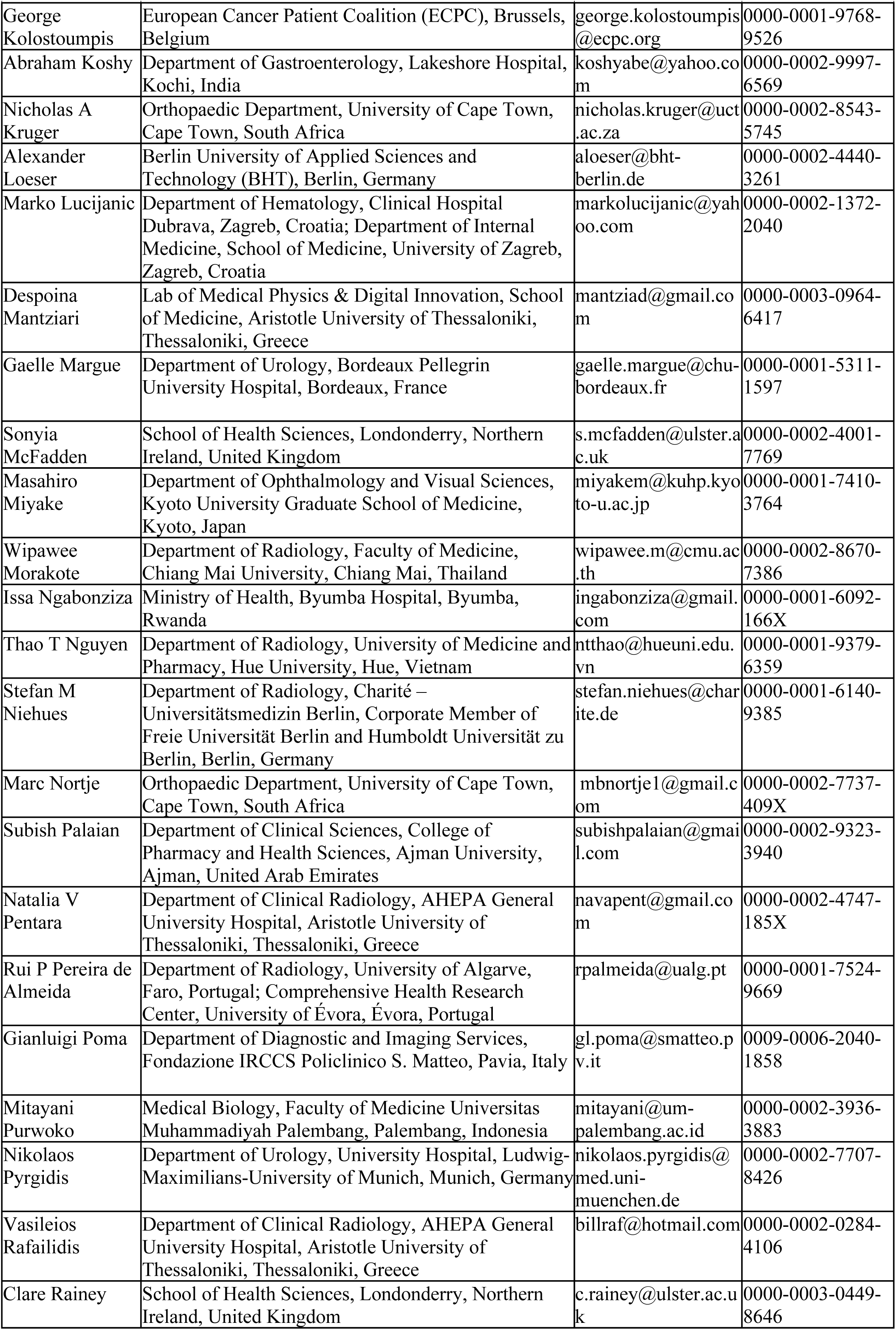
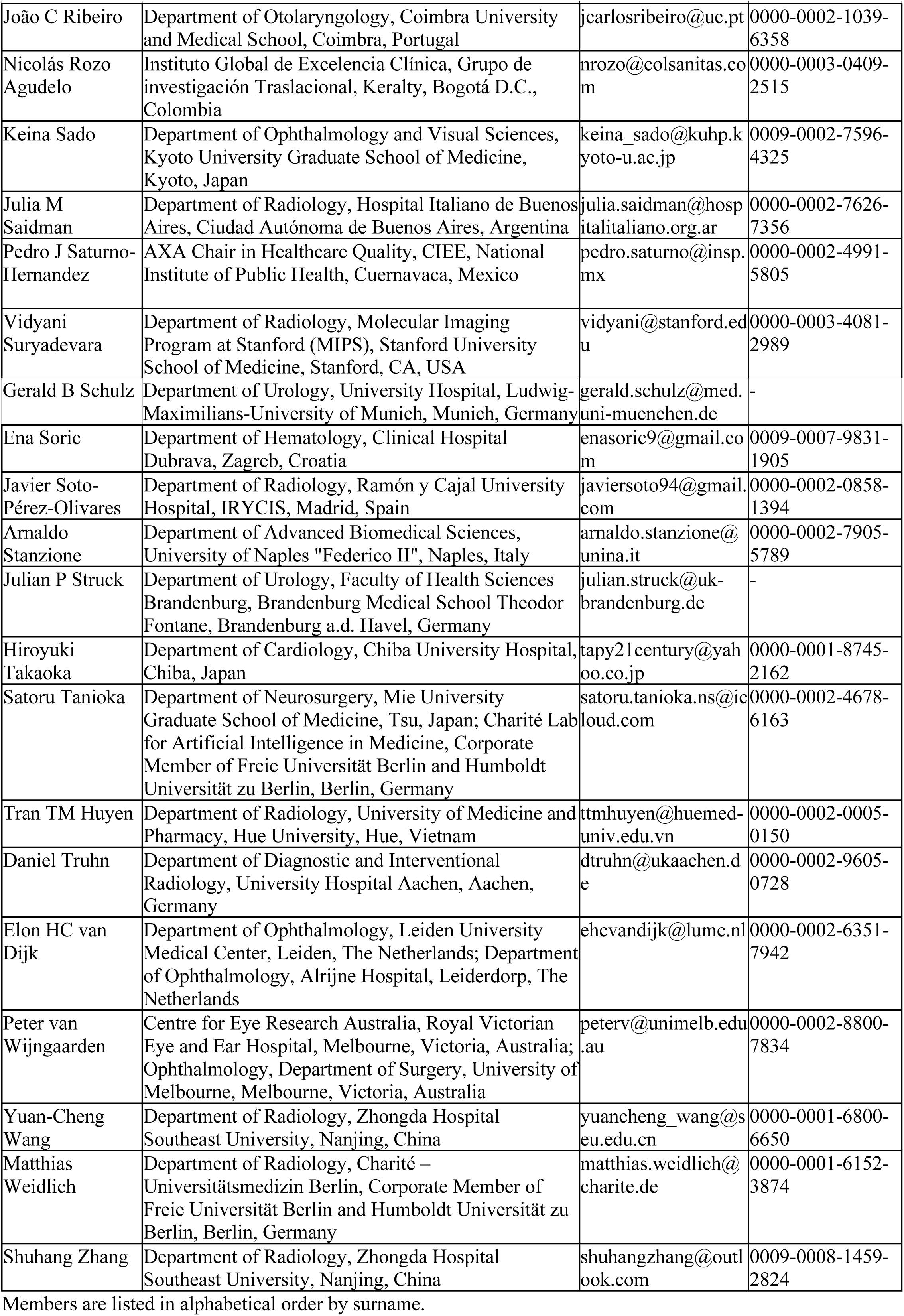

