## Supplementary material for "Multinational attitudes towards AI in healthcare and diagnostics among hospital patients": eTables 1-15

#### eTable 1. Baseline characteristics of excluded patients.

| | Total (N=149) |
|---|---|
| Institution, no. (%) | |
| Charité – University Medicine Berlin, Campus Benjamin Franklin | 127 (85.23%) |
| University of Warmia and Mazury | 8 (5.37%) |
| Faculty of Health Sciences Brandenburg, Brandenburg Medical School Theodor Fontane | 3 (2.01%) |
| Jinling Hospital, Affiliated Hospital of Medical School, Nanjing University | 3 (2.01%) |
| Coimbra University and Medical School | 2 (1.34%) |
| Butabika National Referral Mental Hospital | 1 (0.67%) |
| Dr. B.R.A. Institute Rotary Cancer Hospital, All India Institute of Medical Sciences | 1 (0.67%) |
| Max Institute of Cancer Care, Vaishali | 1 (0.67%) |
| Royal Victoria Hospital | 1 (0.67%) |
| Universidad de Costa Rica | 1 (0.67%) |
| University of Cape Town | 1 (0.67%) |
| Gender, no. (%) | - |
| Female | 63 (42.28%) |
| Male | 77 (51.68%) |
| Diverse | 0 |
| Not reported | 6 (4.03%) |
| Age | - |
| Median (IQR), y | 62 (53-74) |
| Not reported, no. (%) | 58 (38.93%) |
| Highest educational level, no. (%) | - |
| Elementary School Diploma | 23 (15.44%) |
| Middle School Diploma | 23 (15.44%) |
| High School Diploma | 10 (6.71%) |
| University Degree | 27 (18.12%) |
| Not reported | 76 (51.01%) |
| Which of these technical devices do you use at least once a week? | - |
| Median (IQR), total no. | 2 (1-3) |
| Smartphone, no. (%) | 59 (39.6%) |
| PC/laptop, no. (%) | 32 (21.48%) |
| Game console (eg, PlayStation, Switch), no. (%) | 3 (2.01%) |
| Tablet (eg, iPad), no. (%) | 17 (11.41%) |
| E-reader, no. (%) | 3 (2.01%) |
| Smartwatch, no. (%) | 10 (6.71%) |
| None, no. (%) | 6 (4.03%) |
| Not reported, no. (%) | 91 (61.07%) |
| What is your current general state of health?, no. (%) | - |
| Very poor | 1 (0.67%) |
| Poor | 8 (5.37%) |
| Sufficient | 19 (12.75%) |
| Good | 18 (12.08%) |
| Very good | 10 (6.71%) |
| Not reported | 102 (68.46%) |

Abbreviation: IQR, interquartile range.

### eTable 2. Overview of participating institutions, departments, and patients.

| Country/Institution | City | Department | Patients, no. (%) |
|---|---|---|---|
| Total | - | - | 13806 |
| Argentina | - | - | 421 (3.05%) |
| 1. Center for Medical Education and Clinical Research (CEMIC) | Autonomous City of Buenos Aires | Radiology | 329 (2.38%) |
| 2. Hospital Italiano de Buenos Aires | Autonomous City of Buenos Aires | Radiology | 92 (0.67%) |
| Australia | - | - | 221 (1.60%) |
| 3. Royal Victorian Eye and Ear Hospital | Melbourne | Ophthalmology | 100 (0.72%) |
| 4. The Alfred Hospital | Melbourne | Radiology | 121 (0.88%) |
| Austria | - | - | 225 (1.63%) |
| 5. Medical University of Vienna | Vienna | Orthopedics, Trauma Surgery | 225 (1.63%) |
| Azerbaijan | - | - | 100 (0.72%) |
| 6. Azerbaijan Medical University | Baku | Surgery | 100 (0.72%) |
| Brazil | - | - | 581 (4.21%) |
| 7. A.C.Camargo Cancer Center | São Paulo | Radiology | 130 (0.94%) |
| 8. Federal University of Rio Grande do Norte | Natal | Collective Health | 100 (0.72%) |
| 9. Pontifical Catholic University of Rio Grande do Sul | Porto Alegre | Radiology | 100 (0.72%) |
| 10. Universidade Federal de São Paulo | São Paulo | Radiology | 251 (1.82%) |
| Bulgaria | - | - | 255 (1.85%) |
| 11. Karidad Medical Health Center | Plovdiv | Cardiology | 255 (1.85%) |
| Canada | - | - | 816 (5.91%) |
| 12. Montreal General Hospital | Montreal | Radiology | 299 (2.17%) |
| 13. Royal Victoria Hospital | Montreal | Radiology | 457 (3.31%) |
| 14. Lachine Hospital | Montreal | Radiology | 60 (0.43%) |
| China | | | 1315 (9.52%) |
| 15. Jinling Hospital, Affiliated Hospital of Medical School, Nanjing University | Nanjing | Radiology | 1019 (7.38%) |
| 16. Renji Hospital, Shanghai Jiao Tong University School of Medicine | Shanghai | Rheumatology | 160 (1.16%) |
| 17. Zhongda Hospital, Southeast University | Nanjing | Radiology | 136 (0.99%) |
| Colombia | - | - | 100 (0.72%) |
| 18. Fundación Universitaria Sanitas | Bogotá D.C. | Primary care | 100 (0.72%) |
| Costa Rica | - | - | 258 (1.87%) |
| 19. Universidad de Costa Rica | San José | Radiology | 258 (1.87%) |
| Croatia | - | - | 100 (0.72%) |
| 20. Clinical Hospital Dubrava | Zagreb | Hematology | 100 (0.72%) |
| Czech Republic | - | - | 148 (1.07%) |
| 21. Masaryk University | Brno | Infectious diseases | 148 (1.07%) |
| Ecuador | - | - | 234 (1.69%) |
| 22. Universidad de Las Américas | Quito | Respiratory Medicine | 234 (1.69%) |
| France | - | - | 103 (0.75%) |
| 23. University of Bordeaux | Bordeaux | Urology | 103 (0.75%) |
| Germany | - | - | 2091 (15.15%) |
| 24. Charité – University Medicine Berlin, Campus Benjamin Franklin | Berlin | Radiology | 1639 (11.87%) |
| 25. Ludwig Maximilian University of Munich | Munich | Radiology | 134 (0.97%) |

**eTable 2. Overview of participating institutions, departments, and patients.**

| Country/Institution | City | Department | Patients, no. (%) |
|---|---|---|---|
| 26. Faculty of Health Sciences Brandenburg, Brandenburg Medical School Theodor Fontane | Neuruppin | Urology | 139 (1.01%) |
| 27. Technical University of Munich | Munich | Radiology | 165 (1.20%) |
| 28. University Hospital Aachen | Aachen | Radiology | 14 (0.10%) |
| Ghana | - | - | 172 (1.25%) |
| 29. RAAJ Specialist Scan | Cape Coast | Radiology | 50 (0.36%) |
| 30. Sonotech Medical Center | Accra | Radiology | 100 (0.72%) |
| 31. Wenchi Methodist Hospital | Wenchi | Radiology | 22 (0.16%) |
| Greece | - | - | 187 (1.35%) |
| 32. Aristotle University of Thessaloniki | Thessaloniki | Urology | 187 (1.35%) |
| Hungary | - | - | 145 (1.05%) |
| 33. Semmelweis University | Budapest | Otorhinolaryngology, Head and Neck Surgery | 145 (1.05%) |
| India | - | - | 300 (2.17%) |
| 34. Dr. B.R.A. Institute Rotary Cancer Hospital, All India Institute of Medical Sciences | New Delhi | Oncology | 100 (0.72%) |
| 35. Max Institute of Cancer Care, Vaishali | Ghaziabad | Oncology | 98 (0.71%) |
| 36. VPS Lakeshore Hospital | Kochi | Gastroenterology | 102 (0.74%) |
| Indonesia | - | - | 138 (1.00%) |
| 37. Muhammadiyah University of Palembang | Palembang | Neurology | 138 (1.00%) |
| Italy | - | - | 368 (2.67%) |
| 38. Azienda Ospedaliero - Universitaria di Cagliari | Cagliari | Radiology | 106 (0.77%) |
| 39. University of Naples "Federico II" | Naples | Radiology | 127 (0.92%) |
| 40. University of Pavia | Pavia | Gastroenterology | 135 (0.98%) |
| Japan | - | - | 556 (4.03%) |
| 41. Chiba University | Chiba | Cardiology | 111 (0.80%) |
| 42. Hiroshima City Hiroshima Citizens Hospital | Hiroshima | Dermatology | 200 (1.45%) |
| 43. Kyoto University Graduate School of Medicine | Kyoto | Ophthalmology | 138 (1.00%) |
| 44. Mie Chuo Medical Center | Tsu | Neurosurgery | 107 (0.78%) |
| Mexico | - | - | 1110 (8.04%) |
| 45. National Institute of Cardiology Ignacio Chavez | Mexico City | Cardiology | 249 (1.80%) |
| 46. Salvador Zubirán National Institute of Health Sciences and Nutrition | Mexico City | Gastroenterology, Unit of Liver Transplantation | 861 (6.24%) |
| Nepal | - | - | 125 (0.91%) |
| 47. KIST Medical College and Teaching Hospital | Kathmandu | Outpatient | 125 (0.91%) |
| Nigeria | - | - | 173 (1.25%) |
| 48. University of Ilorin Teaching Hospital | Ilorin | Anesthesiology | 173 (1.25%) |
| Poland | - | - | 469 (3.40%) |
| 49. Medical University of Warsaw | Warsaw | Neurology | 158 (1.14%) |
| 50. University of Warmia and Mazury | Olsztyn | Ophthalmology | 163 (1.18%) |
| 51. Wroclaw Medical University | Wroclaw | Otolaryngology, Head and Neck Surgery | 148 (1.07%) |
| Portugal | - | - | 348 (2.52%) |

**eTable 2. Overview of participating institutions, departments, and patients.**

| Country/Institution | City | Department | Patients, no. (%) |
|---|---|---|---|
| 52. Centro Hospitalar Vila Nova de Gaia/Espinho | Vila Nova de Gaia | Oncology | 91 (0.66%) |
| 53. Coimbra University and Medical School | Coimbra | Otorhinolaryngology, Head and Neck Surgery | 143 (1.04%) |
| 54. University of Algarve | Faro | Radiology | 114 (0.83%) |
| Republic of Moldova | - | - | 167 (1.21%) |
| 55. Nicolae Testemițanu State University of Medicine and Pharmacy | Chișinău | Respiratory Medicine, Allergology | 167 (1.21%) |
| Republic of North Macedonia | - | - | 104 (0.75%) |
| 56. Ss. Cyril and Methodius University | Skopje | Cardiac Surgery | 104 (0.75%) |
| Rwanda | - | - | 100 (0.72%) |
| 57. Byumba Level 2 Teaching Hospital | Byumba City | Outpatient | 100 (0.72%) |
| Slovenia | - | - | 189 (1.37%) |
| 58. Institute of Oncology Ljubljana | Ljubljana | Oncology | 189 (1.37%) |
| South Africa | - | - | 109 (0.79%) |
| 59. University of Cape Town | Cape Town | Orthopedics | 109 (0.79%) |
| Spain | - | - | 228 (1.65%) |
| 60. Hospital Universitario Fundación Jiménez Díaz | Madrid | Cardiology | 128 (0.93%) |
| 61. University Hospital Ramón y Cajal | Madrid | Radiology | 100 (0.72%) |
| Sweden | - | - | 394 (2.85%) |
| 62. Central Hospital Växjö | Växjö | Radiology | 183 (1.33%) |
| 63. Lund University | Malmö | Radiology | 144 (1.04%) |
| 64. Umeå University | Umeå | Outpatient | 67 (0.49%) |
| Switzerland | - | - | 200 (1.45%) |
| 65. Kantonsspital Glarus | Glarus | Radiology | 200 (1.45%) |
| Thailand | - | - | 128 (0.93%) |
| 66. Chiang Mai University | Chiang Mai | Radiology | 128 (0.93%) |
| The Netherlands | - | - | 43 (0.31%) |
| 67. Leiden University Medical Center | Leiden | Ophthalmology | 43 (0.31%) |
| Tunisia | - | - | 103 (0.75%) |
| 68. Centre Hospitalier Universitaire Hédi Chaker de Sfax | Sfax | Obstetrics, Gynecology | 103 (0.75%) |
| Turkey | - | - | 284 (2.06%) |
| 69. Hacettepe University | Ankara | Nuclear Medicine | 284 (2.06%) |
| Uganda | - | - | 71 (0.51%) |
| 70. Butabika National Referral Mental Hospital | Kampala | Radiology | 71 (0.51%) |
| United Arab Emirates | - | - | 95 (0.69%) |
| 71. Ajman University | Ajman | Oral surgery | 95 (0.69%) |
| United States | - | - | 100 (0.72%) |
| 72. University of Florida | Florida | Radiology | 100 (0.72%) |
| Vietnam | - | - | 432 (3.13%) |
| 73. Hanoi Medical University Hospital | Hanoi | Radiology | 308 (2.23%) |
| 74. Hue University | Hue | Radiology | 124 (0.90%) |

Abbreviation: NA, not available.

**eTable 3. Adjusted odds ratios for self-reported gender from regression analyses.**

| | Male (N=6973) | Female (N=6451) | Diverse (N=32) |
|---|---|---|---|
| Q1[a], no. (%) | 6099 (87.47%) | 5775 (89.52%) | 24 (75%) |
| Adjusted OR (95% CI), *P* value | 1 [Reference] | 0.84 (0.78-0.9), p<.001 | 0.54 (0.24-1.22), p=1 |
| Q2[a], no. (%) | 5970 (85.62%) | 5724 (88.73%) | 23 (71.88%) |
| Adjusted OR (95% CI), *P* value | 1 [Reference] | 0.75 (0.7-0.8), p<.001 | 0.49 (0.2-0.78), p=1 |
| Q3[a], no. (%) | 6079 (87.18%) | 5767 (89.4%) | 24 (75%) |
| Adjusted OR (95% CI), *P* value | 1 [Reference] | 0.77 (0.71-0.82), p<.001 | 0.56 (0.24-1.3), p=1 |
| Q4[a], no. (%) | 6072 (87.08%) | 5755 (89.21%) | 24 (75%) |
| Adjusted OR (95% CI), *P* value | 1 [Reference] | 0.8 (0.74-0.85), p<.001 | 0.36 (0.16-0.79), p=1 |
| Q5[a], no. (%) | 6069 (87.04%) | 5758 (89.26%) | 24 (75%) |
| Adjusted OR (95% CI), *P* value | 1 [Reference] | 0.76 (0.71-0.8), p<.001 | 0.42 (0.2-0.90), p=1 |
| Q6 [a], no. (%) | 6062 (86.94%) | 5753 (89.18%) | 24 (75%) |
| Adjusted OR (95% CI), *P* value | 1 [Reference] | 0.76 (0.71-0.81), p<.001 | 0.35 (0.16-0.78), p=1 |
| Q7[a], no. (%) | 5929 (85.03%) | 5630 (87.27%) | 24 (75%) |
| Adjusted OR (95% CI), *P* value | 1 [Reference] | 0.88 (0.82-0.94), p<.009 | 1.06 (0.5-2.24), p=1 |
| Q8[a], no. (%) | 6050 (86.76%) | 5733 (88.87%) | 24 (75%) |
| Adjusted OR (95% CI), *P* value | 1 [Reference] | 0.76 (0.7-0.81), p<.001 | 0.44 (0.2-1), p=1 |
| Q9[a], no. (%) | 5815 (83.39%) | 5601 (86.82%) | 24 (75%) |
| Adjusted OR (95% CI), *P* value | 1 [Reference] | 0.85 (0.84-0.86), p<.001 | 0.38 (0.17-0.84), p=1 |
| Q10[a], no. (%) | 5804 (83.24%) | 5591 (86.67%) | 24 (75%) |
| Adjusted OR (95% CI), *P* value | 1 [Reference] | 0.77 (0.72-0.83), p<.001 | 0.47 (0.21-1.03), p=1 |
| Q11[a], no. (%) | 5806 (83.26%) | 5596 (86.75%) | 24 (75%) |
| Adjusted OR (95% CI), *P* value | 1 [Reference] | 0.85 (0.84-0.86), p<.001 | 0.59 (0.58-0.6), p<.001 |
| Q12[b], no. (%) | 5657 (81.13%) | 5460 (84.64%) | 24 (75%) |
| Adjusted OR (95% CI), *P* value | 1 [Reference] | 1.02 (0.93-1.12), p=1 | 1.25 (0.47-1.34), p=1 |
| Q13[c], no. (%) | 5533 (79.35%) | 5362 (83.12%) | 24 (75%) |
| Adjusted OR 1 (95% CI), *P* value | 1 [Reference] | 1.02 (0.93-1.12), p=1 | 1.25 (0.47-3.34), p=1 |
| Adjusted OR 2 (95% CI), *P* value | 1 [Reference] | 0.97 (0.89-1.06), p=1 | 0.78 (0.3-2), p=1 |
| Q14[a], no. (%) | 5712 (81.92%) | 5493 (85.15%) | 24 (75%) |
| Adjusted OR (95% CI), *P* value | 1 [Reference] | 0.97 (0.89-1.06), p=1 | 0.74 (0.28-1.94), p=1 |
| Q15[a], no. (%) | 5673 (81.36%) | 5463 (84.68%) | 23 (71.88%) |
| Adjusted OR (95% CI), *P* value | 1 [Reference] | 0.79 (0.78-0.79), p<.001 | 0.54 (0.24-1.22), p=11 |
| Q16[a], no. (%) | 5631 (80.75%) | 5404 (83.77%) | 24 (75%) |
| Adjusted OR (95% CI), *P* value | 1 [Reference] | 0.81 (0.75-0.88), p<.001 | 0.82 (0.35-1.91), p=1 |
| Q17[a], no. (%) | 5688 (81.57%) | 5464 (84.7%) | 24 (75%) |
| Adjusted OR (95% CI), *P* value | 1 [Reference] | 0.9 (0.83-0.96), p=0.74 | 0.59 (0.28-1.27), p=1 |
| Q18[a], no. (%) | 5674 (81.37%) | 5481 (84.96%) | 24 (75%) |

**eTable 3. Adjusted odds ratios for self-reported gender from regression analyses.**

| | Male (N=6973) | Female (N=6451) | Diverse (N=32) |
|---|---|---|---|
| Adjusted OR (95% CI), *P* value | 1 [Reference] | 0.67 (0.62-0.72), p<.001 | 0.48 (0.23-1), p=.05 |
| Q19[a], no. (%) | 5741 (82.33%) | 5516 (85.51%) | 24 (75%) |
| Adjusted OR (95% CI), *P* value | 1 [Reference] | 0.73 (0.67-0.78), p<.001 | 0.56 (0.27-1.18), p=1 |
| Q20[a], no. (%) | 5734 (82.23%) | 5506 (85.35%) | 24 (75%) |
| Adjusted OR (95% CI), *P* value | 1 [Reference] | 0.7 (0.65-0.75), p<.001 | 0.56 (0.27-1.18), p=1 |

[a] Cumulative link mixed models were used for ordinal regression analysis.

[b] A binary mixed-effects regression analysis with the reference category "A high degree of accuracy, the decision path is clearly comprehensible (explainable AI)." compared to "A higher accuracy, the decision path however, is not comprehensible." was performed.

[c] Two binary mixed-effects regression analyses with the reference category "The AI gives a false alarm about as often as it misses a diagnosis." compared to "The AI misses almost no diagnosis, but often gives a false alarm." (adjusted OR 1) and "The AI almost never gives a false alarm, but sometimes misses a diagnosis." (adjusted OR 2) were performed.

Abbreviations: CI, confidence interval; OR, odds ratio; Q1, What are your general views on the use of artificial intelligence (AI) in medicine?; Q2, Artificial intelligence (AI) should be increasingly used in the healthcare sector.; Q3, How much confidence do you have that AI can improve healthcare?; Q4, How much do you trust an AI to provide reliable information about your health?; Q5, How much do you trust an AI to provide accurate information about your diagnosis?; Q6, How much do you trust an AI to provide accurate information about your response to therapy?; Q7, Which of the following statements about a potential application of AI in medicine do you most likely agree with?; Q8, I would trust a highly accurate AI to make a vital decision for me.; Q9, How would you rate it if a certified AI software analyzes X-ray images?; Q10, How would you rate it if a certified AI software diagnoses cancer?; Q11, How would you rate it if a certified AI software would be available to doctors as a second opinion?; Q12/Q13, Suppose an AI makes a diagnosis. What would you prefer?; Q14, Suppose an AI has about the same accuracy as doctors. Which situation for a diagnosis would you prefer?; Q15, What do you think of healthcare facilities (clinics, practices) using AI software to aid in diagnosis?; Q16, Would you prefer to visit healthcare facilities that use AI software? Q17, How concerned are you about AI compromising the protection of your personal data?; Q18, How concerned are you that the use of AI will reduce the contact between physicians and patients?; Q19, How concerned are you that AI could replace human doctors in the future?; Q20, How concerned are you that the use of AI will lead to higher healthcare costs?.

**eTable 4. Adjusted odds ratios for self-reported health status from regression analyses.**

| | Very poor (N=203) | Poor (N=1221) | Sufficient (N=4244) | Good (N=5439) | Very good (N=2553) |
|---|---|---|---|---|---|
| Q1[a], no. (%) | 183 (90.15%) | 1061 (86.9%) | 3699 (87.16%) | 4728 (86.93%) | 2227 (87.23%) |
| Adjusted OR (95% CI), *P* value | 0.15 (0.11-0.21), p<.001 | 0.54 (0.46-0.63), p<.001 | 0.56 (0.5-0.62), p<.001 | 0.71 (0.64-0.78), p<.001 | 1 [Reference] |
| Q2[a], no. (%) | 182 (89.66%) | 1043 (85.42%) | 3653 (86.07%) | 4638 (85.27%) | 2201 (86.21%) |
| Adjusted OR (95% CI), *P* value | 0.21 (0.15-0.3), p<.001 | 0.62 (0.53-0.72), p<.001 | 0.62 (0.55-0.69), p<.001 | 0.77 (0.67-0.85), p<.001 | 1 [Reference] |
| Q3[a], no. (%) | 181 (89.16%) | 1052 (86.16%) | 3685 (86.83%) | 4726 (86.89%) | 2226 (87.19%) |
| Adjusted OR (95% CI), *P* value | 0.21 (0.16-0.3), p<.001 | 0.57 (0.49-0.67), p<.001 | 0.62 (0.56-0.7), p<.001 | 0.75 (0.67-0.82), p<.001 | 1 [Reference] |
| Q4[a], no. (%) | 181 (89.16%) | 1048 (85.83%) | 3675 (86.59%) | 4722 (86.82%) | 2225 (87.15%) |
| Adjusted OR (95% CI), *P* value | 0.27 (0.19-0.37), p<.001 | 0.61 (0.52-0.71), p<.001 | 0.61 (0.54-0.68), p<.001 | 0.78 (0.71-0.86), p<.001 | 1 [Reference] |
| Q5[a], no. (%) | 180 (88.67%) | 1047 (85.75%) | 3676 (86.62%) | 4723 (86.84%) | 2225 (87.15%) |
| Adjusted OR (95% CI), *P* value | 0.31 (0.23-0.42), p<.001 | 0.6 (0.52-0.7), p<.001 | 0.7 (0.6-0.75), p<.001 | 0.85 (0.77-0.93), p=.24 | 1 [Reference] |
| Q6 [a], no. (%) | 181 (89.16%) | 1044 (85.5%) | 3671 (86.5%) | 4720 (86.78%) | 2223 (87.07%) |
| Adjusted OR (95% CI), *P* value | 0.28 (0.2-0.38), p<.001 | 0.59 (0.51-0.69), p<.001 | 0.62 (0.55-0.69), p<.001 | 0.7 (0.7-0.85), p<.001 | 1 [Reference] |
| Q7[a], no. (%) | 181 (89.16%) | 1022 (83.7%) | 3596 (84.73%) | 4607 (84.7%) | 2177 (85.27%) |
| Adjusted OR (95% CI), *P* value | 1.1 (0.8-1.5), p=1 | 0.93 (0.8-1.09), p=1 | 0.93 (0.83-1.03), p=1 | 0.98 (0.89-1.08), p=1 | 1 [Reference] |
| Q8[a], no. (%) | 182 (89.66%) | 1044 (85.5%) | 3670 (86.48%) | 4692 (86.27%) | 2219 (86.92%) |
| Adjusted OR (95% CI), *P* value | 0.45 (0.33-0.62), p<.001 | 0.74 (0.63-0.86), p=0.02 | 0.71 (0.64-0.79), p<.001 | 0.88 (0.8-0.97), p=1 | 1 [Reference] |
| Q9[a], no. (%) | 176 (86.7%) | 998 (81.74%) | 3542 (83.46%) | 4539 (83.45%) | 2185 (85.59%) |
| Adjusted OR (95% CI), *P* value | 0.2 (0.2-0.21), p<.001 | 0.61 (0.6-0.62), p<.001 | 0.59 (0.59-0.6), p<.001 | 0.77 (0.71-0.82), p<.001 | 1 [Reference] |
| Q10[a], no. (%) | 177 (87.19%) | 995 (81.49%) | 3527 (83.11%) | 4536 (83.4%) | 2184 (85.55%) |
| Adjusted OR (95% CI), *P* value | 0.35 (0.25-0.48), p<.001 | 0.7 (0.6-0.82), p=.002 | 0.67 (0.6-0.74), p<.001 | 0.83 (0.75-0.91), p=.04 | 1 [Reference] |
| Q11[a], no. (%) | 176 (86.7%) | 995 (81.49%) | 3535 (83.29%) | 4535 (83.38%) | 2185 (85.59%) |
| Adjusted OR (95% CI), *P* value | 0.33 (0.33-0.34), p<.001 | 0.78 (0.77-0.79), p<.001 | 0.74 (0.73-0.75), p<.001 | 0.89 (0.83-0.95), p=.29 | 1 [Reference] |
| Q12[b], no. (%) | 172 (84.73%) | 964 (78.95%) | 3452 (81.34%) | 4412 (81.12%) | 2141 (83.86%) |
| Adjusted OR (95% CI), *P* value | 1.27 (0.83-1.94), p=1 | 1.09 (0.89-1.33), p=1 | 1.34 (0.99-1.31), p=1 | 1.07 (0.94-1.21), p=1 | 1 [Reference] |
| Q13[c], no. (%) | 168 (82.76%) | 939 (76.9%) | 3394 (79.97%) | 4323 (79.48%) | 2095 (82.06%) |

**eTable 4. Adjusted odds ratios for self-reported health status from regression analyses.**

| | Very poor (N=203) | Poor (N=1221) | Sufficient (N=4244) | Good (N=5439) | Very good (N=2553) |
|---|---|---|---|---|---|
| Adjusted OR 1 (95% CI), *P* value | 1.27 (0.83-1.94), p=1 | 1.09 (0.89-1.33), p=1 | 1.14 (0.99-1.31), p=1 | 1.07 (0.94-1.21), p=1 | 1 [Reference] |
| Adjusted OR 2 (95% CI), *P* value | 1.1 (0.74-1.65), p=1 | 1.07 (0.89-1.29), p=1 | 1.08 (0.95-1.23), p=1 | 1.1 (0.98-1.23), p=1 | 1 [Reference] |
| Q14[a], no. (%) | 172 (84.73%) | 975 (79.85%) | 3476 (81.9%) | 4462 (82.04%) | 2144 (83.98%) |
| Adjusted OR (95% CI), *P* value | 1.14 (0.77-1.67), p=1 | 1.03 (0.85-1.67), p=1 | 0.86 (0.75-0.98), p=1 | 0.95 (0.84-1.07), p=1 | 1 [Reference] |
| Q15[a], no. (%) | 170 (83.74%) | 979 (80.18%) | 3463 (81.6%) | 4428 (81.41%) | 2119 (83%) |
| Adjusted OR (95% CI), *P* value | 0.31 (0.31-0.31), p<.001 | 0.68 (0.68-0.69), p<.001 | 0.66 (0.65-0.67), p<.001 | 0.84 (0.83-0.85), p<.001 | 1 [Reference] |
| Q16[a], no. (%) | 170 (83.74%) | 963 (78.87%) | 3424 (80.68%) | 4394 (80.79%) | 2108 (82.57%) |
| Adjusted OR (95% CI), *P* value | 0.84 (0.51-1.2), p=1 | 0.85 (0.72-1), p=1 | 0.78 (0.69-0.88), p=.01 | 0.9 (0.81-1), p=1 | 1 [Reference] |
| Q17[a], no. (%) | 172 (84.73%) | 978 (80.1%) | 3467 (81.69%) | 4433 (81.5%) | 2126 (83.27%) |
| Adjusted OR (95% CI), *P* value | 1.48 (1.07-2.04), p=1 | 1.29 (1.1-1.5), p=.35 | 1.77 (1.05-1.31), p=.99 | 1.07 (1.97-1.19), p=1 | 1 [Reference] |
| Q18[a], no. (%) | 172 (84.73%) | 980 (80.26%) | 3466 (81.67%) | 4435 (81.54%) | 2126 (83.27%) |
| Adjusted OR (95% CI), *P* value | 1.76 (1.26-2.46), p=.23 | 1.22 (1.04-1.42), p=1 | 1.09 (0.97-1.21), p=1 | 1.07 (0.97-1.18), p=1 | 1 [Reference] |
| Q19[a], no. (%) | 172 (84.73%) | 982 (80.43%) | 3485 (82.12%) | 4480 (82.37%) | 2162 (84.68%) |
| Adjusted OR (95% CI), *P* value | 1.94 (0.96-1.87), p=1 | 1.1 (0.94-1.28), p=1 | 1.1 (0.99-1.23), p=1 | 1.11 (1.01-1.23), p=1 | 1 [Reference] |
| Q20[a], no. (%) | 172 (84.73%) | 979 (80.18%) | 3477 (81.93%) | 4477 (82.31%) | 2159 (84.57%) |
| Adjusted OR (95% CI), *P* value | 1 (1.43-2.8), p=.01 | 1.15 (0.98-1.34), p=1 | 1.02 (0.92-1.14), p=1 | 1.02 (0.92-1.12), p=1 | 1 [Reference] |

[a] Cumulative link mixed models were used for ordinal regression analysis.
[b] A binary mixed-effects regression analysis with the reference category "A high degree of accuracy, the decision path is clearly comprehensible (explainable AI)." compared to "A higher accuracy, the decision path however, is not comprehensible." was performed.
[c] Two binary mixed-effects regression analyses with the reference category "The AI gives a false alarm about as often as it misses a diagnosis." compared to "The AI misses almost no diagnosis, but often gives a false alarm." (adjusted OR 1) and "The AI almost never gives a false alarm, but sometimes misses a diagnosis." (adjusted OR 2) were performed.
Abbreviations: CI, confidence interval; OR, odds ratio; Q1, What are your general views on the use of artificial intelligence (AI) in medicine?; Q2, Artificial intelligence (AI) should be increasingly used in the healthcare sector.; Q3, How much confidence do you have that AI can improve healthcare?; Q4, How much do you trust an AI to provide reliable information about your health?; Q5, How much do you trust an AI to provide accurate information about your diagnosis?; Q6, How much do you trust an AI to provide accurate information about your response to therapy?; Q7, Which of the following statements about a potential application of AI in medicine do you most likely agree with?; Q8, I would trust a highly accurate AI to make a vital decision for me.; Q9, How would you rate it if a certified AI software analyzes X-ray images?; Q10, How would you rate it if a certified AI software diagnoses cancer?; Q11, How would you rate it if a certified AI software would be available to doctors as a second opinion?; Q12/Q13, Suppose an AI makes a diagnosis. What would you prefer?; Q14, Suppose an AI has about the same accuracy as doctors. Which situation for a diagnosis would you prefer?; Q15, What do you think of healthcare facilities (clinics, practices) using AI software to aid in diagnosis?; Q16, Would you prefer to visit healthcare facilities that use AI software? Q17, How concerned are you about AI compromising the protection of your personal data?; Q18, How concerned are you that the use of AI will reduce the contact between physicians and patients?; Q19, How concerned are you that AI could replace human doctors in the future?; Q20, How concerned are you that the use of AI will lead to higher healthcare costs.

**eTable 5. Adjusted odds ratios for self-reported AI knowledge from regression analyses.**

| | No knowledge (N=1848) | Little knowledge (N=8097) | Good knowledge (N=3423) | Expert (N=211) |
|---|---|---|---|---|
| Q1[a], no. (%) | 1532 (82.9%) | 7175 (88.61%) | 3003 (87.73%) | 188 (89.1%) |
| Adjusted OR (95% CI), *P* value | 1 [Reference] | 1.75 (1.56-1.96), p<.001 | 3.17 (2.77-3.63), p<.001 | 7.11 (5.19-9.74), p<.001 |
| Q2[a], no. (%) | 1495 (80.9%) | 7055 (87.13%) | 2978 (87%) | 189 (89.57%) |
| Adjusted OR (95% CI), *P* value | 1 [Reference] | 1.38 (1.24-1.54), p<.001 | 1.48 (1.32-1.67), p<.001 | 3.8 (2.77-5.2), p<.001 |
| Q3[a], no. (%) | 1531 (82.85%) | 7155 (88.37%) | 2995 (87.5%) | 189 (89.57%) |
| Adjusted OR (95% CI), *P* value | 1 [Reference] | 1.46 (1.3-1.64), p<.001 | 2.53 (2.21-2.89), p<.001 | 5.11 (3.76-6.94), p<.001 |
| Q4[a], no. (%) | 1525 (82.52%) | 7143 (88.22%) | 2994 (87.47%) | 189 (89.57%) |
| Adjusted OR (95% CI), *P* value | 1 [Reference] | 1.51 (1.34-1.69), p<.001 | 2.41 (2.11-2.76), p<.001 | 3.26 (2.41-4.41), p<.001 |
| Q5[a], no. (%) | 1528 (82.68%) | 7141 (88.19%) | 2994 (87.47%) | 188 (89.1%) |
| Adjusted OR (95% CI), *P* value | 1 [Reference] | 1.43 (1.27-1.6), p<.001 | 2.27 (1.98-2.59), p<.001 | 3.96 (2.91-5.4), p<.001 |
| Q6 [a], no. (%) | 1527 (82.63%) | 7130 (88.06%) | 2993 (87.44%) | 189 (89.57%) |
| Adjusted OR (95% CI), *P* value | 1 [Reference] | 1.43 (1.27-1.6), p<.001 | 2.27 (1.99-2.6), p<.001 | 3.44 (2.53-4.67), p<.001 |
| Q7[a], no. (%) | 1493 (80.79%) | 6976 (86.16%) | 2930 (85.6%) | 184 (87.2%) |
| Adjusted OR (95% CI), *P* value | 1 [Reference] | 1.89 (1.68-2.12), p<.001 | 2.8 (2.45-3.2), p<.001 | 4.48 (3.26-6.15), p<.001 |
| Q8[a], no. (%) | 1523 (82.41%) | 7108 (87.79%) | 2987 (87.26%) | 189 (89.57%) |
| Adjusted OR (95% CI), *P* value | 1 [Reference] | 1.18 (1.06-1.32), p=.83 | 1.74 (1.53-1.98), p<.001 | 2.06 (1.52-2.8), p<.001 |
| Q9[a], no. (%) | 1484 (80.3%) | 6869 (84.83%) | 2904 (84.84%) | 183 (86.73%) |
| Adjusted OR (95% CI), *P* value | 1 [Reference] | 1.23 (1.22-1.24), p<.001 | 1.98 (1.95-2), p<.001 | 4.44 (3.31-5.97), p<.001 |
| Q10[a], no. (%) | 1477 (79.92%) | 6864 (84.77%) | 2895 (84.57%) | 183 (86.73%) |
| Adjusted OR (95% CI), *P* value | 1 [Reference] | 1.32 (1.17-1.48), p<.001 | 1.91 (1.67-2.19), p<.001 | 3.36 (2.46-4.59), p<.001 |
| Q11[a], no. (%) | 1473 (79.71%) | 6870 (84.85%) | 2901 (84.75%) | 182 (86.26%) |
| Adjusted OR (95% CI), *P* value | 1 [Reference] | 1.35 (1.34-1.37), p<.001 | 1.96 (1.93-1.98), p<.001 | 4.29 (4.23-4.35), p<.001 |
| Q12[b], no. (%) | 1423 (77%) | 6690 (82.62%) | 2848 (83.2%) | 180 (85.31%) |
| Adjusted OR (95% CI), *P* value | 1 [Reference] | 0.86 (0.75-1), p=1 | 0.72 (0.61-0.69), p=.04 | 1.06 (0.72-0.85), p=1 |
| Q13[c], no. (%) | 1400 (75.76%) | 6544 (80.82%) | 2799 (81.77%) | 176 (83.41%) |
| Adjusted OR 1 (95% CI), *P* value | 1 [Reference] | 0.86 (0.75-1), p=1 | 0.72 (0.61-0.69), p=.04 | 1.06 (0.72-0.85), p=1 |
| Adjusted OR 2 (95% CI), *P* value | 1 [Reference] | 1.08 (0.94-1.24), p=1 | 1.11 (0.95-1.3), p=1 | 0.97 (0.67-1.4), p=1 |
| Q14[a], no. (%) | 1440 (77.92%) | 6740 (83.24%) | 2866 (83.73%) | 183 (86.73%) |
| Adjusted OR (95% CI), *P* value | 1 [Reference] | 1.23 (1.06-1.43), p=1 | 1.3 (1.1-1.53), p=.7 | 1.65 (1.16-2.36), p=1 |
| Q15[a], no. (%) | 1410 (76.3%) | 6723 (83.03%) | 2846 (83.14%) | 180 (85.31%) |
| Adjusted OR (95% CI), *P* value | 1 [Reference] | 1.45 (1.44-1.46), p<.001 | 2.3 (2.28-2.32), p<.001 | 4.77 (3.51-6.48), p<.001 |
| Q16[a], no. (%) | 1401 (75.81%) | 6651 (82.14%) | 2827 (82.59%) | 180 (85.31%) |
| Adjusted OR (95% CI), *P* value | 1 [Reference] | 1.38 (1.21-1.56), p<.001 | 1.99 (1.72-2.3), p<.001 | 3.18 (2.29-4.42), p<.001 |
| Q17[a], no. (%) | 1414 (76.52%) | 6728 (83.09%) | 2852 (83.32%) | 182 (86.26%) |
| Adjusted OR (95% CI), *P* value | 1 [Reference] | 0.94 (0.84-1.06), p=1 | 1.15 (1-1.31), p=1 | 1.03 (0.75-1.39), p=1 |
| Q18[a], no. (%) | 1417 (76.68%) | 6730 (83.12%) | 2850 (83.26%) | 182 (86.26%) |

**eTable 5. Adjusted odds ratios for self-reported AI knowledge from regression analyses.**

| | No knowledge (N=1848) | Little knowledge (N=8097) | Good knowledge (N=3423) | Expert (N=211) |
|---|---|---|---|---|
| Adjusted OR (95% CI), *P* value | 1 [Reference] | 1.02 (0.91-1.15), p=1 | 1.45 (1.27-1.66), p<.001 | 1.46 (1.08-2), p=1 |
| Q19[a], no. (%) | 1460 (79%) | 6770 (83.61%) | 2868 (83.79%) | 183 (86.73%) |
| Adjusted OR (95% CI), *P* value | 1 [Reference] | 0.94 (0.84-1.01), p=1 | 1.3 (1.14-1.49), p=.03 | 1.84 (1.35-2.5), p=.03 |
| Q20[a], no. (%) | 1455 (78.73%) | 6763 (83.52%) | 2865 (83.7%) | 181 (85.78%) |
| Adjusted OR (95% CI), *P* value | 1 [Reference] | 1.09 (0.97-1.22), p=1 | 1.6 (1.4-1.8), p<.001 | 1.83 (1.34-2.5), p=.001 |

[a] Cumulative link mixed models were used for ordinal regression analysis.

[b] A binary mixed-effects regression analysis with the reference category "A high degree of accuracy, the decision path is clearly comprehensible (explainable AI)." compared to "A higher accuracy, the decision path however, is not comprehensible." was performed.

[c] Two binary mixed-effects regression analyses with the reference category "The AI gives a false alarm about as often as it misses a diagnosis." compared to "The AI misses almost no diagnosis, but often gives a false alarm." (adjusted OR 1) and "The AI almost never gives a false alarm, but sometimes misses a diagnosis." (adjusted OR 2) were performed.

Abbreviations: CI, confidence interval; OR, odds ratio; Q1, What are your general views on the use of artificial intelligence (AI) in medicine?; Q2, Artificial intelligence (AI) should be increasingly used in the healthcare sector.; Q3, How much confidence do you have that AI can improve healthcare?; Q4, How much do you trust an AI to provide reliable information about your health?; Q5, How much do you trust an AI to provide accurate information about your diagnosis?; Q6, How much do you trust an AI to provide accurate information about your response to therapy?; Q7, Which of the following statements about a potential application of AI in medicine do you most likely agree with?; Q8, I would trust a highly accurate AI to make a vital decision for me.; Q9, How would you rate it if a certified AI software analyzes X-ray images?; Q10, How would you rate it if a certified AI software diagnoses cancer?; Q11, How would you rate it if a certified AI software would be available to doctors as a second opinion?; Q12/Q13, Suppose an AI makes a diagnosis. What would you prefer?; Q14, Suppose an AI has about the same accuracy as doctors. Which situation for a diagnosis would you prefer?; Q15, What do you think of healthcare facilities (clinics, practices) using AI software to aid in diagnosis?; Q16, Would you prefer to visit healthcare facilities that use AI software? Q17, How concerned are you about AI compromising the protection of your personal data?; Q18, How concerned are you that the use of AI will reduce the contact between physicians and patients?; Q19, How concerned are you that AI could replace human doctors in the future?; Q20, How concerned are you that the use of AI will lead to higher healthcare costs?.

**eTable 6 Adjusted odds ratios for age and number of technical devices used weekly from regression analyses.**

| | **Age (N=12452)** | **Total number tech devices (N=13752)** |
|---|---|---|
| Q1[a], no. (%) | 11898 (95.55%) | 11898 (86.52%) |
| Adjusted OR (95% CI), *P* value | 0.99 (0.78-1.26), p=1 | 1.17 (1.13-1.21), p<.001 |
| Q2[a], no. (%) | 11717 (94.10%) | 11717 (85.20%) |
| Adjusted OR (95% CI), *P* value | 1.07 (0.84-1.36), p=1 | 1.14 (1.1-1.18), p<.001 |
| Q3[a], no. (%) | 11870 (95.33%) | 11870 (86.31%) |
| Adjusted OR (95% CI), *P* value | 0.77 (0.61-0.98), p=1 | 1.15 (1.11-1.19), p<.001 |
| Q4[a], no. (%) | 11851 (95.17%) | 11851 (86.18%) |
| Adjusted OR (95% CI), *P* value | 0.95 (0.75-1.2), p=1 | 1.13 (1.09-1.17), p<.001 |
| Q5[a], no. (%) | 11851 (95.17%) | 11851 (86.18%) |
| Adjusted OR (95% CI), *P* value | 1.43 (1.27-1.6), p=1 | 1.1 (1.06-1.14), p<.001 |
| Q6 [a], no. (%) | 11839 (95.08%) | 11839 (86.09%) |
| Adjusted OR (95% CI), *P* value | 0.88 (0.7-1.12), p=1 | 1.07 (1.04-1.11), p=.0.2 |
| Q7[a], no. (%) | 11583 (93.02%) | 11583 (84.23%) |
| Adjusted OR (95% CI), *P* value | 1.62 (1.28-2.06), p<.02 | 1.13 (1.09-1.17), p<.001 |
| Q8[a], no. (%) | 11807 (94.82%) | 11807 (85.86%) |
| Adjusted OR (95% CI), *P* value | 1.41 (1.11-1.78), p=1 | 1.18 (1.06-1.02), p=1 |
| Q9[a], no. (%) | 11440 (91.87%) | 11440 (83.19%) |
| Adjusted OR (95% CI), *P* value | 0.89 (0.88-0.9), p<.001 | 1.09 (1.08-1.11), p<.001 |
| Q10[a], no. (%) | 11419 (91.70%) | 11419 (83.04%) |
| Adjusted OR (95% CI), *P* value | 1.32 (1.17-1.48), p=.13 | 1.32 (1.17-1.13), p<.001 |
| Q11[a], no. (%) | 11426 (91.76%) | 11426 (83.09%) |
| Adjusted OR (95% CI), *P* value | 1.05 (1.03-1.06), p<.001 | 1.15 (1.14-1.17), p<.001 |
| Q12[b], no. (%) | 11141 (89.47%) | 11141 (81.01%) |
| Adjusted OR (95% CI), *P* value | 1.62 (1.18-2.15), p=.76 | 0.91 (0.87-0.95), p=.02 |
| Q13[c], no. (%) | 10919 (87.69%) | 10919 (79.40%) |
| Adjusted OR 1 (95% CI), *P* value | 1.12 (1.18-2.21), p=.76 | 0.91 (0.87-0.95), p=.02 |
| Adjusted OR 2 (95% CI), *P* value | 2.12 (1.59-2.83), p<.001 | 0.92 (0.88-0.96), p=.01 |
| Q14[a], no. (%) | 11229 (90.18%) | 11229 (81.65%) |
| Adjusted OR (95% CI), *P* value | 0.5 (0.37-0.66), p<.001 | 0.98 (0.94-1.03), p=1 |
| Q15[a], no. (%) | 11159 (89.62%) | 11159 (81.14%) |
| Adjusted OR (95% CI), *P* value | 0.91 (0.9-0.92), p<.001 | 1.16 (1.15-1.17), p<.001 |
| Q16[a], no. (%) | 11059 (88.81%) | 11059 (80.42%) |
| Adjusted OR (95% CI), *P* value | 0.83 (0.64-1.07), p=1 | 1.05 (1.02-1.09), p=1 |
| Q17[a], no. (%) | 11176 (89.75%) | 11176 (81.27%) |
| Adjusted OR (95% CI), *P* value | 0.62 (0.49-0.79), p=.03 | 1.09 (1.06-1.13), p<.001 |
| Q18[a], no. (%) | 11179 (89.78%) | 11179 (81.29%) |
| Adjusted OR (95% CI), *P* value | 0.36 (0.29-0.46), p<.001 | 1.03 (1-1.07), p=1 |
| Q19[a], no. (%) | 11281 (90.60%) | 11281 (82.03%) |
| Adjusted OR (95% CI), *P* value | 0.39 (0.3-0.49), p<.001 | 1.06 (1.03-1.1), p=.12 |
| Q20[a], no. (%) | 11264 (90.46%) | 11264 (81.91%) |
| Adjusted OR (95% CI), *P* value | 0.5 (0.4-0.64), p<.001 | 1.06 (1.03-1.1), p=.21 |

[a] Cumulative link mixed models were used for ordinal regression analysis.
[b] A binary mixed-effects regression analysis with the reference category "A high degree of accuracy, the decision path is clearly comprehensible (explainable AI)." compared to "A higher accuracy, the decision path however, is not comprehensible." was performed.
[c] Two binary mixed-effects regression analyses with the reference category "The AI gives a false alarm about as often as it misses a diagnosis." compared to "The AI misses almost no diagnosis, but often gives a false alarm." (adjusted OR 1) and "The AI almost never gives a false alarm, but sometimes misses a diagnosis." (adjusted OR 2) were performed.
Abbreviations: CI, confidence interval; OR, odds ratio; Q1, What are your general views on the use of artificial intelligence (AI) in medicine?; Q2, Artificial intelligence (AI) should be increasingly used in the healthcare sector.; Q3, How much confidence do you have that AI can improve healthcare?; Q4, How much do you trust an AI to provide reliable information about your health?; Q5, How much do you trust an AI to provide accurate information about your diagnosis?; Q6, How much do you trust an AI to provide accurate information about your response to therapy?; Q7, Which of the following statements about a potential application of AI in medicine do you most likely agree with?; Q8, I would trust a highly accurate AI to make a vital decision for me.; Q9, How would you rate it if a certified AI software analyzes X-ray images?; Q10, How would you rate it if a certified AI software diagnoses cancer?; Q11, How would you rate it if a certified AI software would be available to doctors as a second opinion?; Q12/Q13, Suppose an AI makes a diagnosis. What would you prefer?; Q14, Suppose an AI has about the same accuracy as doctors. Which situation for a diagnosis would you prefer?; Q15, What do you think of healthcare facilities (clinics,

practices) using AI software to aid in diagnosis?; Q16, Would you prefer to visit healthcare facilities that use AI software? Q17, How concerned are you about AI compromising the protection of your personal data?; Q18, How concerned are you that the use of AI will reduce the contact between physicians and patients?; Q19, How concerned are you that AI could replace human doctors in the future?; Q20, How concerned are you that the use of AI will lead to higher healthcare costs?.

**eTable 7. Adjusted odds ratios for highest educational level from regression analyses.**

| | Elementary School (N=1192) | Middle School (N=2844) | High School (N=4142) | University (N=5403) |
|---|---|---|---|---|
| Q1[a], no. (%) | 961 (80.62%) | 2427 (85.34%) | 3783 (91.33%) | 4727 (87.49%) |
| Adjusted OR (95% CI), *P* value | 1 [Reference] | 0.93 (0.81-1.08), p=1 | 1.03 (0.89-1.19), p=1 | 1.27 (1.1-1.48), p=1 |
| Q2[a], no. (%) | 922 (77.35%) | 2356 (82.84%) | 3748 (90.49%) | 4691 (86.82%) |
| Adjusted OR (95% CI), *P* value | 1 [Reference] | 0.88 (0.76-1.01), p=1 | 1.02 (0.89-1.17), p=1 | 1.05 (0.91-1.2), p=1 |
| Q3[a], no. (%) | 955 (80.12%) | 2423 (85.2%) | 3777 (91.19%) | 4715 (87.27%) |
| Adjusted OR (95% CI), *P* value | 1 [Reference] | 0.9 (0.78-1.04), p=1 | 0.94 (0.81-1.1), p=1 | 1.15 (1-1.33), p=1 |
| Q4[a], no. (%) | 951 (79.78%) | 2411 (84.77%) | 3774 (91.12%) | 4715 (87.27%) |
| Adjusted OR (95% CI), *P* value | 1 [Reference] | 0.86 (0.76-1.01), p=1 | 0.98 (0.93-1.06), p=1 | 1.03 (0.89-1.19), p=1 |
| Q5[a], no. (%) | 952 (79.87%) | 2411 (84.77%) | 3774 (91.12%) | 4714 (87.25%) |
| Adjusted OR (95% CI), *P* value | 1 [Reference] | 0.86 (0.74-0.99), p=1 | 0.86 (0.74-1), p=1 | 0.97 (0.84-1.12), p=1 |
| Q6 [a], no. (%) | 944 (79.19%) | 2414 (84.88%) | 3770 (91.02%) | 4711 (87.19%) |
| Adjusted OR (95% CI), *P* value | 1 [Reference] | 0.84 (0.73-0.97), p=1 | 0.81 (0.7-0.94), p=1 | 0.93 (0.81-1.08), p=1 |
| Q7[a], no. (%) | 915 (76.76%) | 2361 (83.02%) | 3695 (89.21%) | 4612 (85.36%) |
| Adjusted OR (95% CI), *P* value | 1 [Reference] | 1.09 (0.94-1.26), p=1 | 1.38 (1.19-1.6), p=.005 | 1.68 (1.45-1.95), p<.001 |
| Q8[a], no. (%) | 942 (79.03%) | 2404 (84.53%) | 3766 (90.92%) | 4695 (86.9%) |
| Adjusted OR (95% CI), *P* value | 1 [Reference] | 0.84 (0.73-0.97), p=1 | 0.78 (0.68-0.9), p=.17 | 0.81 (0.85-0.93), p=.93 |
| Q9[a], no. (%) | 898 (75.34%) | 2306 (81.08%) | 3700 (89.33%) | 4536 (83.95%) |
| Adjusted OR (95% CI), *P* value | 1 [Reference] | 0.88 (0.88-0.9), p<.001 | 0.98 (0.97-0.99), p=1 | 1.22 (1.21-1.24), p<.001 |
| Q10[a], no. (%) | 896 (75.17%) | 2297 (80.77%) | 3693 (89.16%) | 4533 (83.9%) |
| Adjusted OR (95% CI), *P* value | 1 [Reference] | 0.86 (0.74-1), p=1 | 0.91 (0.79-1.06), p=1 | 0.99 (0.85-1.14), p=1 |
| Q11[a], no. (%) | 895 (75.08%) | 2301 (80.91%) | 3690 (89.09%) | 4540 (84.03%) |
| Adjusted OR (95% CI), *P* value | 1 [Reference] | 0.98 (0.97-1), p=1 | 1.08 (1.07-1.1), p<.001 | 1.37 (1.35-1.39), p<.001 |
| Q12[b], no. (%) | 859 (72.06%) | 2206 (77.57%) | 3602 (86.96%) | 4474 (82.81%) |
| Adjusted OR (95% CI), *P* value | 1 [Reference] | 1.03 (0.86-1.24), p=1 | 1.09 (0.9-1.31), p=1 | 1 (0.83-1.2), p=1 |
| Q13[c], no. (%) | 822 (68.96%) | 2161 (75.98%) | 3549 (85.68%) | 4387 (81.2%) |
| Adjusted OR 1 (95% CI), *P* value | 1 [Reference] | 1.03 (0.86-1.24), p=1 | 1.09 (0.9-1.31), p=1 | 1 (0.82-1.2), p=1 |
| Adjusted OR 2 (95% CI), *P* value | 1 [Reference] | 0.92 (0.77-1.1), p=1 | 0.97 (0.81-1.56), p=1 | 1 (0.84-1.2), p=1 |
| Q14[a], no. (%) | 872 (73.15%) | 2253 (79.22%) | 3636 (87.78%) | 4468 (82.69%) |
| Adjusted OR (95% CI), *P* value | 1 [Reference] | 0.86 (0.72-1.03), p=1 | 0.82 (0.68-0.99), p=1 | 0.76 (0.64-0.92), p=1 |
| Q15[a], no. (%) | 874 (73.32%) | 2254 (79.25%) | 3600 (86.91%) | 4431 (82.01%) |
| Adjusted OR (95% CI), *P* value | 1 [Reference] | 0.81 (0.8-0.81), p<.001 | 0.93 (0.92-0.94), p<.001 | 1.18 (1.17-1.19), p<.001 |
| Q16[a], no. (%) | 854 (71.64%) | 2234 (78.55%) | 3564 (86.05%) | 4407 (81.57%) |
| Adjusted OR (95% CI), *P* value | 1 [Reference] | 0.84 (0.72-1), p=1 | 0.91 (0.77-1.07), p=1 | 1.16 (0.98-1.37), p=1 |
| Q17[a], no. (%) | 873 (73.24%) | 2248 (79.04%) | 3611 (87.18%) | 4444 (82.25%) |
| Adjusted OR (95% CI), *P* value | 1 [Reference] | 1.08 (0.93-1.25), p=1 | 1.03 (0.89-1.2), p=1 | 1.03 (0.89-1.19), p=1 |
| Q18[a], no. (%) | 871 (73.07%) | 2255 (79.29%) | 3607 (87.08%) | 4446 (82.29%) |

**eTable 7. Adjusted odds ratios for highest educational level from regression analyses.**

| | Elementary School (N=1192) | Middle School (N=2844) | High School (N=4142) | University (N=5403) |
|---|---|---|---|---|
| Adjusted OR (95% CI), *P* value | 1 [Reference] | 0.91 (0.78-1.06), p=1 | 0.86 (0.74-1), p=1 | 0.87 (0.74-1.01), p=1 |
| Q19[a], no. (%) | 880 (73.83%) | 2270 (79.82%) | 3655 (88.24%) | 4476 (82.84%) |
| Adjusted OR (95% CI), *P* value | 1 [Reference] | 1.01 (0.87-1.17), p=1 | 0.94 (0.8-1.09), p=1 | 1.13 (0.97-1.3), p=1 |
| Q20[a], no. (%) | 879 (73.74%) | 2271 (79.85%) | 3657 (88.29%) | 4457 (82.49%) |
| Adjusted OR (95% CI), *P* value | 1 [Reference] | 1.14 (0.99-1.33), p=1 | 1.06 (0.91-1.23), p=1 | 1.27 (1.11-1.5), p=.27 |

[a] Cumulative link mixed models were used for ordinal regression analysis.
[b] A binary mixed-effects regression analysis with the reference category "A high degree of accuracy, the decision path is clearly comprehensible (explainable AI)." compared to "A higher accuracy, the decision path however, is not comprehensible." was performed.
[c] Two binary mixed-effects regression analyses with the reference category "The AI gives a false alarm about as often as it misses a diagnosis." compared to "The AI misses almost no diagnosis, but often gives a false alarm." (adjusted OR 1) and "The AI almost never gives a false alarm, but sometimes misses a diagnosis." (adjusted OR 2) were performed.
Abbreviations: CI, confidence interval; OR, odds ratio; Q1, What are your general views on the use of artificial intelligence (AI) in medicine?; Q2, Artificial intelligence (AI) should be increasingly used in the healthcare sector.; Q3, How much confidence do you have that AI can improve healthcare?; Q4, How much do you trust an AI to provide reliable information about your health?; Q5, How much do you trust an AI to provide accurate information about your diagnosis?; Q6, How much do you trust an AI to provide accurate information about your response to therapy?; Q7, Which of the following statements about a potential application of AI in medicine do you most likely agree with?; Q8, I would trust a highly accurate AI to make a vital decision for me.; Q9, How would you rate it if a certified AI software analyzes X-ray images?; Q10, How would you rate it if a certified AI software diagnoses cancer?; Q11, How would you rate it if a certified AI software would be available to doctors as a second opinion?; Q12/Q13, Suppose an AI makes a diagnosis. What would you prefer?; Q14, Suppose an AI has about the same accuracy as doctors. Which situation for a diagnosis would you prefer?; Q15, What do you think of healthcare facilities (clinics, practices) using AI software to aid in diagnosis?; Q16, Would you prefer to visit healthcare facilities that use AI software? Q17, How concerned are you about AI compromising the protection of your personal data?; Q18, How concerned are you that the use of AI will reduce the contact between physicians and patients?; Q19, How concerned are you that AI could replace human doctors in the future?; Q20, How concerned are you that the use of AI will lead to higher healthcare costs?

### eTable 8. Absolute survey results for each item stratified by gender.

| | Male (N=6973) | Female (N=6451) | Diverse (N=32) |
|---|---|---|---|
| Q1, no. (%) | 6864 (98.44%) | 6318 (97.94%) | 31 (96.88%) |
| Extremely negative | 150 (2.19%) | 141 (2.23%) | 2 (6.45%) |
| Rather negative | 417 (6.08%) | 596 (9.43%) | 5 (16.13%) |
| Neutral | 2240 (32.63%) | 2070 (32.76%) | 6 (19.35%) |
| Rather positive | 2836 (41.32%) | 2593 (41.04%) | 11 (35.48%) |
| Extremely positive | 1221 (17.79%) | 918 (14.53%) | 7 (22.58%) |
| Q2, no. (%) | 6708 (96.20%) | 6302 (97.69%) | 29 (90.63%) |
| Completely disagree | 167 (2.49%) | 252 (4%) | 3 (10.34%) |
| Tend to disagree | 454 (6.77%) | 590 (9.36%) | 7 (24.14%) |
| Neutral | 1727 (25.75%) | 1657 (26.29%) | 4 (13.79%) |
| Tend to agree | 2549 (38 %) | 2503 (39.72%) | 6 (20.69%) |
| Agree completely | 1811 (27 %) | 1300 (20.63%) | 9 (31.03%) |
| Q3, no. (%) | 6861 (98.39%) | 6357 (98.54%) | 30 (93.75%) |
| Very little | 219 (3.19 %) | 296 (4.66%) | 3 (10%) |
| Little | 668 (9.74 %) | 850 (13.37%) | 4 (13.33%) |
| Medium | 2448 (35.68%) | 2349 (36.95%) | 10 (33.33%) |
| Much | 2361 (34.41%) | 2122 (33.38%) | 4 (13.33%) |
| Very much | 1165 (16.98%) | 740 (11.64%) | 9 (30%) |
| Q4, no. (%) | 6847 (98.19%) | 6344 (98.34%) | 30 (93.75%) |
| Very little | 219 (3.2%) | 331 (5.22%) | 3 (10%) |
| Little | 888 (12.97%) | 938 (14.79%) | 6 (20%) |
| Medium | 2551 (37.26%) | 2492 (39.28%) | 11 (36.67%) |
| Much | 2427 (35.45%) | 2079 (32.77%) | 5 (16.67%) |
| Very much | 762 (11.13%) | 504 (7.94%) | 5 (16.67%) |
| Q5, no. (%) | 6838 (98.06%) | 6342 (98.31%) | 30 (93.75%) |
| Very little | 243 (3.55%) | 343 (5.41%) | 3 (10%) |
| Little | 875 (12.8%) | 1007 (15.88%) | 6 (20%) |
| Medium | 2522 (36.88%) | 2473 (38.99%) | 12 (40%) |
| Much | 2412 (35.27%) | 2015 (31.77%) | 5 (16.67%) |
| Very much | 786 (11.49%) | 504 (7.95%) | 4 (13.33%) |
| Q6, no. (%) | 6826 (97.89%) | 6337 (98.23%) | 30 (93.75%) |
| Very little | 244 (3.57%) | 355 (5.6%) | 6 (20%) |
| Little | 915 (13.4%) | 1013 (15.99%) | 4 (13.33%) |
| Medium | 2591 (37.96%) | 2566 (40.49%) | 11 (36.67%) |
| Much | 2339 (34.27%) | 1927 (30.41%) | 4 (13.33%) |
| Very much | 737 (10.8%) | 476 (7.51%) | 5 (16.67%) |
| Q7, no. (%) | 6656 (95.45%) | 6185 (95.88%) | 28 (87.50%) |
| No use of AI independent of the disease. | 690 (10.37%) | 756 (12.22%) | 3 (10.71%) |
| Use only for minor illnesses (eg, cold). | 1698 (25.51%) | 1876 (30.33%) | 5 (17.86%) |
| Use also for moderately severe diseases (eg, appendicitis). | 1872 (28.13%) | 1549 (25.04%) | 10 (35.71%) |
| Use also for severe diseases (eg, cancer, traffic accidents). | 2396 (36%) | 2004 (32.4%) | 10 (35.71%) |
| Q8, no. (%) | 6812 (97.69%) | 6310 (97.81%) | 29 (90.63%) |
| Completely disagree | 629 (9.23%) | 765 (12.12%) | 7 (24.14%) |
| Tend to disagree | 1197 (17.57%) | 1380 (21.87%) | 5 (17.24%) |
| Neutral | 1942 (28.51%) | 1650 (26.15%) | 3 (10.34%) |
| Tend to agree | 2257 (33.13%) | 1992 (31.57%) | 8 (17.24%) |
| Agree completely | 787 (11.55%) | 523 (8.29%) | 6 (24.14%) |

### eTable 8. Absolute survey results for each item stratified by gender.

| | Male (N=6973) | Female (N=6451) | Diverse (N=32) |
|---|---|---|---|
| Q9, no. (%) | 6486 (93.02%) | 6145 (95.26%) | 30 (93.75%) |
| Extremely negative | 152 (2.34%) | 220 (3.58%) | 3 (10%) |
| Rather negative | 516 (7.96%) | 684 (11.13%) | 5 (16.67%) |
| Neutral | 1886 (29.08 %) | 1698 (27.63%) | 8 (26.67%) |
| Rather positive | 2651 (40.87%) | 2542 (41.37%) | 8 (26.67%) |
| Extremely positive | 1281 (19.75%) | 1001 (16.29%) | 6 (20%) |
| Q10, no. (%) | 6466 (92.73%) | 6134 (95.09%) | 30 (93.75%) |
| Extremely negative | 206 (3.19%) | 346 (5.64%) | 3 (10%) |
| Rather negative | 724 (11.2%) | 820 (13.37%) | 6 (20%) |
| Neutral | 1853 (28.66%) | 1781 (29.03%) | 6 (20%) |
| Rather positive | 2491 (38.52%) | 2311 (37.68%) | 10 (33.33%) |
| Extremely positive | 1192 (18.43%) | 876 (14.28%) | 5 (16.67%) |
| Q11, no. (%) | 6472 (92.82%) | 6142 (95.21%) | 30 (93.75%) |
| Extremely negative | 141 (2.18%) | 190 (3.09%) | 3 (10%) |
| Rather negative | 346 (5.35 %) | 473 (7.7%) | 3 (10%) |
| Neutral | 1489 (23.01%) | 1403 (22.84%) | 5 (16.67%) |
| Rather positive | 2672 (41.29%) | 2576 (41.94%) | 10 (33.33%) |
| Extremely positive | 1824 (28.18%) | 1500 (24.42%) | 9 (30%) |
| Q12, no. (%) | 6284 (90.12%) | 5964 (92.45%) | 30 (93.75%) |
| A high degree of accuracy, the decision path is clearly comprehensible (explainable AI). | 4333 (68.95%) | 4271 (71.61%) | 21 (70%) |
| A higher accuracy, the decision path however, is not comprehensible. | 1951 (31.05%) | 1693 (28.39%) | 9 (30%) |
| Q13, no. (%) | 6117 (87.72%) | 5832 (90.4%) | 30 (93.75%) |
| The AI misses almost no diagnosis, but often gives a false alarm. | 2792 (45.64%) | 2761 (47.34%) | 17 (56.67%) |
| The AI almost never gives a false alarm, but sometimes misses a diagnosis. | 2260 (36.95%) | 2077 (35.61%) | 9 (30%) |
| The AI gives a false alarm about as often as it misses a diagnosis. | 1065 (17.41%) | 994 (17.04%) | 4 (13.33%) |
| Q14, no. (%) | 6341 (90.94%) | 5976 (92.64%) | 30 (93.75%) |
| Physicians make the diagnosis alone. | 379 (5.98%) | 435 (7.28%) | 4 (13.33%) |
| AI and physicians make the diagnosis together. Doctors make the final decision. | 4715 (74.36%) | 4301 (71.97%) | 18 (60%) |
| AI and physicians make the diagnosis together. Both have equal authority. | 787 (12.41%) | 875 (14.64%) | 4 (13.33%) |
| AI and physicians make the diagnosis together. The AI makes the final decision. | 160 (2.52%) | 143 (2.39%) | 5 (16.67%) |
| AI makes the diagnosis alone. | 290 (4.57%) | 222 (3.71%) | 2 (6.67%) |

### eTable 8. Absolute survey results for each item stratified by gender.

| | Male (N=6973) | Female (N=6451) | Diverse (N=32) |
|---|---|---|---|
| Q15, no. (%) | 6329 (90.76%) | 5990 (92.85%) | 29 (90.63%) |
| Extremely negative | 140 (2.21%) | 177 (2.95%) | 1 (3.45%) |
| Rather negative | 420 (6.64%) | 455 (7.6%) | 4 (13.79%) |
| Neutral | 1728 (27.3%) | 1773 (29.6%) | 9 (31.03%) |
| Rather positive | 2609 (41.22%) | 2502 (41.77%) | 7 (24.14%) |
| Extremely positive | 1432 (22.63%) | 1083 (18.08%) | 8 (27.59%) |
| Q16, no. (%) | 6263 (89.82%) | 5912 (91.64%) | 30 (93.75%) |
| Always prefer facilities without AI. | 324 (5.17%) | 431 (7.29%) | 3 (10%) |
| Tend to prefer facilities without AI. | 1321 (21.09%) | 1420 (24.02%) | 5 (16.67%) |
| Rather prefer facilities with AI. | 3584 (57.22%) | 3296 (55.75%) | 14 (46.67%) |
| Always prefer facilities with AI. | 1034 (16.51%) | 765 (12.94%) | 8 (26.67%) |
| Q17, no. (%) | 6339 (90.91%) | 5993 (92.9%) | 30 (93.75%) |
| Very worried | 1051 (16.58%) | 1208 (20.16%) | 8 (26.67%) |
| Somewhat concerned | 2131 (33.62%) | 2161 (36.06%) | 9 (30%) |
| Neutral | 1908 (30.1%) | 1601 (26.71%) | 4 (13.33%) |
| Rather unconcerned | 856 (13.5%) | 731 (12.2%) | 5 (16.67%) |
| Unconcerned | 393 (6.2%) | 292 (4.87%) | 4 (13.33%) |
| Q18, no. (%) | 6320 (90.64%) | 6012 (93.19%) | 30 (93.75%) |
| Very worried | 1305 (20.65%) | 1737 (28.89%) | 8 (26.67%) |
| Somewhat concerned | 2337 (36.98%) | 2251 (37.44%) | 12 (40%) |
| Neutral | 1659 (26.25%) | 1217 (20.24%) | 6 (20%) |
| Rather unconcerned | 746 (11.8%) | 613 (10.2%) | 2 (6.67%) |
| Unconcerned | 273 (4.32%) | 194 (3.23%) | 2 (6.67%) |
| Q19, no. (%) | 6392 (91.67%) | 6043 (93.68%) | 30 (93.75%) |
| Very worried | 1587 (24.83%) | 2185 (36.16%) | 9 (30%) |
| Somewhat concerned | 2085 (32.62%) | 1844 (30.51%) | 12 (40%) |
| Neutral | 1440 (22.53%) | 1077 (17.82%) | 6 (20%) |
| Rather unconcerned | 875 (13.69%) | 649 (10.74%) | 1 (3.33%) |
| Unconcerned | 505 (7.9%) | 288 (4.77%) | 2 (6.67%) |
| Q20, no. (%) | 6381 (91.51%) | 6035 (93.55%) | 30 (93.75%) |
| Very worried | 1412 (22.13%) | 1841 (30.51%) | 6 (20%) |
| Somewhat concerned | 1976 (30.97%) | 1899 (31.47%) | 11 (36.67%) |
| Neutral | 1804 (28.27%) | 1470 (24.36%) | 8 (26.67%) |
| Rather unconcerned | 779 (12.21%) | 547 (9.06%) | 4 (13.33%) |
| Unconcerned | 410 (6.43%) | 278 (4.61%) | 1 (3.33%) |

Abbreviations: Q1, What are your general views on the use of artificial intelligence (AI) in medicine?; Q2, Artificial intelligence (AI) should be increasingly used in the healthcare sector.; Q3, How much confidence do you have that AI can improve healthcare?; Q4, How much do you trust an AI to provide reliable information about your health?; Q5, How much do you trust an AI to provide accurate information about your diagnosis?; Q6, How much do you trust an AI to provide accurate information about your response to therapy?; Q7, Which of the following statements about a potential application of AI in medicine do you most likely agree with?; Q8, I would trust a highly accurate AI to make a vital decision for me.; Q9, How would you rate it if a certified AI software analyzes X-ray images?; Q10, How would you rate it if a certified AI software diagnoses cancer?; Q11, How would you rate it if a certified AI software would be available to doctors as a second opinion?; Q12/Q13, Suppose an AI makes a diagnosis. What would you prefer?; Q14, Suppose an AI has about the same accuracy as doctors. Which situation for a diagnosis would you prefer?; Q15, What do you think of healthcare facilities (clinics, practices) using AI software to aid in diagnosis?; Q16, Would you prefer to visit healthcare facilities that use AI software? Q17, How concerned are you about AI compromising the protection of your personal data?; Q18, How concerned are you that the use of AI will reduce the contact between physicians and patients?; Q19, How concerned are you that AI could replace human doctors in the future?; Q20, How concerned are you that the use of AI will lead to higher healthcare costs?.

**eTable 9. Absolute survey results for each item stratified by self-reported health status.**

| | **Very poor (N=203)** | **Poor (N=1221)** | **Sufficient (N=4244)** | **Good (N=5439)** | **Very good (N=2553)** |
|---|---|---|---|---|---|
| Q1, no. (%) | 199 (98.03%) | 1185 (97.05%) | 4160 (98.02%) | 5351 (98.38%) | 2538 (99.41%) |
| Extremely negative | 53 (26.63%) | 78 (6.58%) | 77 (1.85%) | 54 (1.01%) | 33 (1.3%) |
| Rather negative | 58 (29.15%) | 231 (19.49%) | 282 (6.78%) | 318 (5.94%) | 134 (5.28%) |
| Neutral | 52 (26.13%) | 390 (32.91%) | 1700 (40.87%) | 1628 (30.42%) | 608 (23.96%) |
| Rather positive | 22 (11.06%) | 324 (27.34%) | 1592 (38.27%) | 2547 (47.6%) | 1035 (40.78%) |
| Extremely positive | 14 (7.04%) | 162 (13.67%) | 509 (12.27%) | 804 (15.03%) | 718 (28.29%) |
| Q2, no. (%) | 198 (97.54%) | 1169 (95.74%) | 4114 (96.94%) | 5258 (96.67%) | 2501 (97.96%) |
| Completely disagree | 64 (32.32%) | 106 (9.07%) | 113 (2.75%) | 89 (1.69%) | 54 (2.16%) |
| Tend to disagree | 49 (24.75%) | 231 (19.76%) | 303 (7.37%) | 327 (6.22%) | 140 (5.6%) |
| Neutral | 39 (19.7%) | 295 (25.24%) | 1334 (32.43%) | 1284 (24.42%) | 475 (18.99%) |
| Tend to agree | 22 (11.11%) | 297 (25.41%) | 1560 (37.92%) | 2296 (43.67%) | 961 (38.42%) |
| Agree completely | 24 (12.12%) | 240 (20.53%) | 804 (19.54%) | 1262 (24%) | 871 (34.83%) |
| Q3, no. (%) | 195 (96.06%) | 1190 (97.46%) | 4164 (98.11%) | 5377 (98.86%) | 2534 (99.26%) |
| Very little | 57 (29.23%) | 86 (7.23%) | 157 (3.77%) | 147 (2.73%) | 77 (3.04%) |
| Little | 49 (25.13%) | 290 (24.37%) | 462 (11.1%) | 521 (9.69%) | 212 (8.37%) |
| Medium | 53 (27.18%) | 385 (32.35%) | 1829 (43.92%) | 1884 (35.04%) | 713 (28.14%) |
| Much | 24 (12.31%) | 288 (24.2%) | 1238 (29.73%) | 2113 (39.3%) | 884 (34.89%) |
| Very much | 12 (6.15%) | 141 (11.85%) | 428 (10.28%) | 712 (13.24%) | 648 (25.57%) |
| Q4, no. (%) | 196 (96.55%) | 1184 (96.97%) | 4144 (97.64%) | 5375 (98.82%) | 2530 (99.1%) |
| Very little | 48 (24.49%) | 82 (6.39%) | 177 (4.27%) | 156 (2.9%) | 99 (3.91%) |
| Little | 57 (29.08%) | 272 (22.97%) | 619 (14.94%) | 647 (12.04%) | 255 (10.08%) |
| Medium | 57 (29.08%) | 433 (36.57%) | 1775 (42.83%) | 2046 (38.07%) | 797 (31.5%) |
| Much | 22 (11.22%) | 290 (24.49%) | 1275 (30.77%) | 2032 (37.8%) | 949 (37.51%) |
| Very much | 12 (6.12%) | 107 (9.04%) | 298 (7.19%) | 494 (9.19%) | 430 (17%) |
| Q5, no. (%) | 194 (95.57%) | 1183 (96.89%) | 4143 (97.62%) | 5373 (98.79%) | 2531 (99.14%) |
| Very little | 35 (18.04%) | 71 (6%) | 185 (4.47%) | 183 (3.41%) | 120 (4.74%) |
| Little | 64 (32.99%) | 285 (24.09%) | 648 (15.64%) | 644 (11.99%) | 268 (10.59%) |
| Medium | 62 (31.96%) | 437 (36.94%) | 1709 (41.25%) | 2039 (37.95%) | 807 (31.88%) |
| Much | 23 (11.86%) | 290 (24.51%) | 1282 (30.94%) | 2005 (37.32%) | 900 (35.56%) |
| Very much | 10 (5.15%) | 98 (8.28%) | 321 (7.75%) | 502 (9.34%) | 436 (17.23%) |
| Q6, no. (%) | 195 (96.06%) | 1176 (96.31%) | 4139 (97.53%) | 5368 (98.69%) | 2529 (99.06%) |
| Very little | 38 (19.49%) | 77 (6.55%) | 190 (4.59%) | 191 (3.56%) | 115 (4.55%) |
| Little | 62 (31.79%) | 269 (22.87%) | 648 (15.66%) | 691 (12.87%) | 276 (10.91%) |
| Medium | 61 (31.28%) | 442 (37.59%) | 1791 (43.27%) | 2130 (39.68%) | 809 (31.99%) |
| Much | 24 (12.31%) | 295 (25.09%) | 1204 (29.09%) | 1897 (35.34%) | 909 (35.94%) |
| Very much | 10 (5.13%) | 93 (7.91%) | 306 (7.39%) | 459 (8.55%) | 420 (16.61%) |
| Q7, no. (%) | 195 (96.06%) | 1146 (93.86%) | 4033 (95.03%) | 5222 (96.01%) | 2473 (96.87%) |
| No use of AI independent of the disease. | 15 (7.69%) | 138 (12.04%) | 474 (11.75%) | 527 (10.09%) | 339 (13.71%) |
| Use only for minor illnesses (eg, cold). | 71 (36.41%) | 316 (27.57%) | 1191 (29.53%) | 1472 (28.19%) | 595 (24.06%) |

**eTable 9. Absolute survey results for each item stratified by self-reported health status.**

| | Very poor (N=203) | Poor (N=1221) | Sufficient (N=4244) | Good (N=5439) | Very good (N=2553) |
|---|---|---|---|---|---|
| Use also for moderately severe diseases (eg, appendicitis). | 51 (26.15%) | 315 (27.49%) | 1081 (26.8%) | 1444 (27.65%) | 598 (24.18%) |
| Use also for severe diseases (eg, cancer, traffic accidents). | 58 (29.74%) | 377 (32.9%) | 1287 (31.91%) | 1779 (34.07%) | 941 (38.05%) |
| Q8, no. (%) | 196 (96.55%) | 1172 (95.99%) | 4129 (97.29%) | 5338 (98.14%) | 2526 (98.94%) |
| Completely disagree | 52 (26.53%) | 154 (13.14%) | 438 (10.61%) | 489 (9.16%) | 261 (10.33%) |
| Tend to disagree | 65 (33.16%) | 315 (26.88%) | 781 (18.91%) | 1018 (19.07%) | 414 (16.39%) |
| Neutral | 29 (14.8%) | 305 (26.02%) | 1331 (32.24%) | 1399 (26.21%) | 478 (18.92%) |
| Tend to agree | 35 (17.86%) | 271 (23.12%) | 1258 (30.47%) | 1920 (35.97%) | 868 (34.36%) |
| Agree completely | 16 (8.16%) | 127 (10.84%) | 321 (7.77%) | 512 (9.59%) | 405 (16.03%) |
| Q9, no. (%) | 190 (93.6%) | 1119 (91.65%) | 3950 (93.07%) | 5127 (94.26%) | 2478 (97.06%) |
| Extremely negative | 42 (22.11%) | 96 (8.58%) | 102 (2.58%) | 82 (1.6%) | 51 (2.06%) |
| Rather negative | 52 (27.37%) | 196 (17.52%) | 413 (10.46%) | 397 (7.74%) | 161 (6.5%) |
| Neutral | 50 (26.32%) | 297 (26.54%) | 1387 (35.11%) | 1368 (26.68%) | 541 (21.83%) |
| Rather positive | 28 (14.74%) | 358 (31.99%) | 1492 (37.77%) | 2341 (45.66%) | 1039 (41.93%) |
| Extremely positive | 18 (9.47%) | 172 (15.37%) | 556 (14.08%) | 939 (18.31%) | 686 (27.68%) |
| Q10, no. (%) | 191 (94.09%) | 1116 (91.4%) | 3930 (92.6%) | 5121 (94.15%) | 2475 (96.94%) |
| Extremely negative | 41 (21.47%) | 110 (9.86%) | 163 (4.15%) | 153 (2.99%) | 91 (3.68%) |
| Rather negative | 41 (21.47%) | 197 (17.65%) | 558 (14.2%) | 536 (10.47%) | 235 (9.49%) |
| Neutral | 61 (31.94%) | 309 (27.69%) | 1279 (32.54%) | 1460 (28.51%) | 595 (24.04%) |
| Rather positive | 34 (17.8%) | 340 (30.47%) | 1423 (36.21%) | 2109 (41.18%) | 959 (38.75%) |
| Extremely positive | 14 (7.33%) | 160 (14.34%) | 507 (12.9%) | 863 (16.85%) | 595 (24.04%) |
| Q11, no. (%) | 190 (93.6%) | 1116 (91.4%) | 3941 (92.86%) | 5117 (94.08%) | 2477 (97.02%) |
| Extremely negative | 41 (21.58%) | 76 (6.81%) | 83 (2.11%) | 78 (1.52%) | 61 (2.46%) |
| Rather negative | 52 (27.37%) | 178 (15.95%) | 230 (5.84%) | 255 (4.98%) | 115 (4.64%) |
| Neutral | 40 (21.05%) | 143 (12.81%) | 1133 (28.75%) | 1064 (20.79%) | 469 (18.93%) |
| Rather positive | 35 (18.42%) | 373 (33.42%) | 1658 (42.07%) | 2307 (45.09%) | 958 (38.68%) |
| Extremely positive | 22 (11.58%) | 246 (22.04%) | 837 (21.24%) | 1413 (27.61%) | 876 (35.37%) |
| Q12, no. (%) | 184 (90.64%) | 1075 (88.04%) | 3828 (90.2%) | 4953 (91.06%) | 2418 (94.71%) |
| A high degree of accuracy, the decision path is clearly | 140 (76.09%) | 786 (73.12%) | 2548 (66.56%) | 3532 (71.31%) | 1745 (72.17%) |

**eTable 9. Absolute survey results for each item stratified by self-reported health status.**

| | Very poor (N=203) | Poor (N=1221) | Sufficient (N=4244) | Good (N=5439) | Very good (N=2553) |
|---|---|---|---|---|---|
| comprehensible (explainable AI). | | | | | |
| A higher accuracy, the decision path however, is not comprehensible. | 44 (23.91%) | 189 (17.58%) | 1280 (33.44%) | 1421 (28.69%) | 653 (27.01%) |
| Q13, no. (%) | 177 (87.19%) | 1037 (84.93%) | 3760 (88.6%) | 4842 (89.02%) | 2363 (92.56%) |
| The AI misses almost no diagnosis, but often gives a false alarm. | 96 (54.24%) | 470 (45.32%) | 1633 (43.43%) | 2272 (46.92%) | 1192 (50.44%) |
| The AI almost never gives a false alarm, but sometimes misses a diagnosis. | 51 (28.81%) | 343 (33.08%) | 1425 (37.9%) | 1779 (36.74%) | 827 (35%) |
| The AI gives a false alarm about as often as it misses a diagnosis. | 30 (16.95%) | 224 (21.6%) | 702 (18.67%) | 791 (16.34%) | 344 (14.56%) |
| Q14, no. (%) | 184 (90.64%) | 1076 (88.12%) | 3842 (90.53%) | 5018 (92.26%) | 2423 (94.91%) |
| Physicians make the diagnosis alone. | 8 (4.35%) | 74 (6.88%) | 293 (7.63%) | 302 (6.02%) | 144 (5.94%) |
| AI and physicians make the diagnosis together. Doctors make the final decision. | 9 (4.89%) | 30 (2.79%) | 2790 (72.62%) | 3701 (73.75%) | 1727 (71.28%) |
| AI and physicians make the diagnosis together. Both have equal authority. | 27 (14.67%) | 158 (14.68%) | 540 (14.06%) | 687 (13.69%) | 300 (12.38%) |
| AI and physicians make the | 138 (75%) | 783 (72.77%) | 84 (2.19%) | 125 (2.49%) | 68 (2.81%) |

**eTable 9. Absolute survey results for each item stratified by self-reported health status.**

| | Very poor (N=203) | Poor (N=1221) | Sufficient (N=4244) | Good (N=5439) | Very good (N=2553) |
|---|---|---|---|---|---|
| diagnosis together. The AI makes the final decision. | | | | | |
| AI makes the diagnosis alone. | 2 (1.09%) | 31 (2.88%) | 135 (3.51%) | 203 (4.05%) | 184 (7.59%) |
| Q15, no. (%) | 182 (89.66%) | 1096 (89.76%) | 3860 (90.95%) | 5005 (92.02%) | 2406 (94.24%) |
| Extremely negative | 57 (31.32%) | 84 (7.66%) | 79 (2.05%) | 50 (1%) | 47 (1.95%) |
| Rather negative | 42 (23.08%) | 179 (16.33%) | 325 (8.42%) | 251 (5.01%) | 90 (3.74%) |
| Neutral | 35 (19.23%) | 309 (28.19%) | 1331 (34.48%) | 1367 (27.31%) | 525 (21.82%) |
| Rather positive | 30 (16.48%) | 346 (31.57%) | 1504 (38.96%) | 2281 (45.57%) | 1024 (42.56%) |
| Extremely positive | 18 (9.89%) | 178 (16.24%) | 621 (16.09%) | 1056 (21.1%) | 720 (29.93%) |
| Q16, no. (%) | 182 (89.66%) | 1075 (88.04%) | 3805 (89.66%) | 4953 (91.06%) | 2386 (93.46%) |
| Always prefer facilities without AI. | 18 (9.89%) | 108 (10.05%) | 247 (6.49%) | 258 (5.21%) | 134 (5.62%) |
| Tend to prefer facilities without AI. | 25 (13.74%) | 271 (25.21%) | 933 (24.52%) | 1089 (21.99%) | 466 (19.53%) |
| Rather prefer facilities with AI. | 99 (54.4%) | 545 (50.7%) | 2188 (57.5%) | 2862 (57.78%) | 1280 (53.65%) |
| Always prefer facilities with AI. | 40 (21.98%) | 151 (14.05%) | 437 (11.48%) | 744 (15.02%) | 506 (21.21%) |
| Q17, no. (%) | 184 (90.64%) | 1094 (89.6%) | 3863 (91.02%) | 5007 (92.06%) | 2412 (94.48%) |
| Very worried | 25 (13.59%) | 184 (16.82%) | 611 (15.82%) | 928 (18.53%) | 552 (22.89%) |
| Somewhat concerned | 61 (33.15%) | 327 (29.89%) | 1344 (34.79%) | 1806 (36.07%) | 842 (34.91%) |
| Neutral | 38 (20.65%) | 335 (30.62%) | 1211 (31.35%) | 1417 (28.3%) | 567 (23.51%) |
| Rather unconcerned | 38 (20.65%) | 159 (14.53%) | 474 (12.27%) | 341 (6.81%) | 297 (12.31%) |
| Unconcerned | 22 (11.96%) | 89 (8.14%) | 223 (5.77%) | 215 (4.29%) | 154 (6.38%) |
| Q18, no. (%) | 183 (90.15%) | 1096 (89.76%) | 3861 (90.98%) | 5012 (92.15%) | 2409 (94.36%) |
| Very worried | 25 (13.66%) | 271 (24.73%) | 878 (22.74%) | 1215 (24.24%) | 702 (29.14%) |
| Somewhat concerned | 55 (30.05%) | 358 (32.66%) | 1468 (38.02%) | 1984 (39.59%) | 797 (33.08%) |
| Neutral | 37 (20.22%) | 254 (23.18%) | 1007 (26.08%) | 1127 (22.49%) | 512 (21.25%) |
| Rather unconcerned | 41 (22.4%) | 153 (13.96%) | 394 (10.2%) | 534 (10.65%) | 265 (11%) |
| Unconcerned | 25 (13.66%) | 60 (5.47%) | 114 (2.95%) | 152 (3.03%) | 133 (5.52%) |
| Q19, no. (%) | 184 (90.64%) | 1097 (89.84%) | 3881 (91.45%) | 5058 (93%) | 2443 (95.69%) |
| Very worried | 43 (23.37%) | 333 (30.36%) | 1107 (28.52%) | 1536 (30.37%) | 806 (32.99%) |
| Somewhat concerned | 40 (21.74%) | 330 (30.08%) | 1256 (32.36%) | 1664 (32.9%) | 697 (28.53%) |
| Neutral | 31 (16.85%) | 217 (19.78%) | 886 (22.83%) | 898 (17.75%) | 453 (18.54%) |

**eTable 9. Absolute survey results for each item stratified by self-reported health status.**

| | Very poor (N=203) | Poor (N=1221) | Sufficient (N=4244) | Good (N=5439) | Very good (N=2553) |
|---|---|---|---|---|---|
| Rather unconcerned | 40 (21.74%) | 146 (13.31%) | 439 (11.31%) | 608 (12.02%) | 326 (13.34%) |
| Unconcerned | 30 (16.3%) | 71 (6.47%) | 139 (3.58%) | 261 (1.16%) | 161 (6.59%) |
| Q20, no. (%) | 184 (90.64%) | 1095 (89.68%) | 3870 (91.19%) | 5053 (92.9%) | 2444 (95.73%) |
| Very worried | 38 (20.65%) | 275 (25.11%) | 993 (25.66%) | 1282 (25.37%) | 717 (29.34%) |
| Somewhat concerned | 34 (18.48%) | 321 (29.32%) | 1261 (32.58%) | 1656 (32.77%) | 692 (28.31%) |
| Neutral | 32 (17.39%) | 189 (17.26%) | 1071 (27.67%) | 1343 (26.58%) | 588 (24.06%) |
| Rather unconcerned | 38 (20.65%) | 128 (11.69%) | 366 (9.46%) | 532 (10.53%) | 284 (11.62%) |
| Unconcerned | 42 (22.83%) | 82 (7.49%) | 178 (4.6%) | 240 (4.75%) | 163 (6.67%) |

Abbreviations: Q1, What are your general views on the use of artificial intelligence (AI) in medicine?; Q2, Artificial intelligence (AI) should be increasingly used in the healthcare sector.; Q3, How much confidence do you have that AI can improve healthcare?; Q4, How much do you trust an AI to provide reliable information about your health?; Q5, How much do you trust an AI to provide accurate information about your diagnosis?; Q6, How much do you trust an AI to provide accurate information about your response to therapy?; Q7, Which of the following statements about a potential application of AI in medicine do you most likely agree with?; Q8, I would trust a highly accurate AI to make a vital decision for me.; Q9, How would you rate it if a certified AI software analyzes X-ray images?; Q10, How would you rate it if a certified AI software diagnoses cancer?; Q11, How would you rate it if a certified AI software would be available to doctors as a second opinion?; Q12/Q13, Suppose an AI makes a diagnosis. What would you prefer?; Q14, Suppose an AI has about the same accuracy as doctors. Which situation for a diagnosis would you prefer?; Q15, What do you think of healthcare facilities (clinics, practices) using AI software to aid in diagnosis?; Q16, Would you prefer to visit healthcare facilities that use AI software? Q17, How concerned are you about AI compromising the protection of your personal data?; Q18, How concerned are you that the use of AI will reduce the contact between physicians and patients?; Q19, How concerned are you that AI could replace human doctors in the future?; Q20, How concerned are you that the use of AI will lead to higher healthcare costs?.

**eTable 10. Absolute survey results for each item stratified by self-reported AI knowledge.**

| | No knowledge (N=1848) | Little knowledge (N=8097) | Good knowledge (N=3423) | Expert (N=211) |
|---|---|---|---|---|
| Q1, no. (%) | 1755 (94.97%) | 8054 (99.47%) | 3402 (99.39%) | 210 (99.53%) |
| Extremely negative | 78 (4.44%) | 122 (1.51%) | 84 (2.47%) | 7 (3.33%) |
| Rather negative | 151 (8.6%) | 669 (8.31%) | 196 (5.76%) | 9 (4.29%) |
| Neutral | 859 (48.95%) | 2879 (35.75%) | 620 (18.22%) | 19 (9.05%) |
| Rather positive | 398 (22.68%) | 3396 (42.17%) | 1647 (48.41%) | 80 (38.1%) |
| Extremely positive | 269 (15.33%) | 988 (12.27%) | 855 (25.13%) | 95 (45.24%) |
| Q2, no. (%) | 1703 (92.15%) | 7898 (97.54%) | 3367 (98.36%) | 210 (99.53%) |
| Completely disagree | 90 (5.28%) | 205 (2.6%) | 115 (3.42%) | 8 (3.81%) |
| Tend to disagree | 137 (8.04%) | 692 (8.76%) | 212 (6.3%) | 9 (4.29%) |
| Neutral | 714 (41.93%) | 2183 (27.64%) | 493 (14.64%) | 26 (12.38%) |
| Tend to agree | 388 (22.78%) | 3259 (41.26%) | 1409 (41.85%) | 63 (30%) |
| Agree completely | 374 (21.96%) | 1559 (19.74%) | 1138 (33.8%) | 104 (49.52%) |
| Q3, no. (%) | 1756 (95.02%) | 8021 (99.06%) | 3396 (99.21%) | 211 (100%) |
| Very little | 135 (7.69%) | 269 (3.35%) | 106 (3.12%) | 7 (3.32%) |
| Little | 283 (16.12%) | 959 (11.96%) | 256 (7.54%) | 14 (6.64%) |
| Medium | 708 (40.32%) | 3247 (40.48%) | 873 (25.71%) | 27 (12.8%) |
| Much | 386 (21.98%) | 2651 (33.05%) | 1404 (41.34%) | 73 (34.6%) |
| Very much | 244 (13.9%) | 895 (11.16%) | 757 (22.29%) | 90 (42.65%) |
| Q4, no. (%) | 1748 (94.59%) | 8008 (98.9%) | 3390 (99.04%) | 210 (99.53%) |
| Very little | 143 (8.18%) | 284 (3.55%) | 116 (3.42%) | 6 (2.86%) |
| Little | 411 (23.51%) | 1075 (13.42%) | 324 (9.56%) | 21 (10%) |
| Medium | 613 (35.07%) | 3407 (42.54%) | 1033 (30.47%) | 52 (24.76%) |
| Much | 354 (20.25%) | 2674 (33.39%) | 1432 (42.24%) | 77 (36.67%) |
| Very much | 227 (12.99%) | 568 (7.09%) | 485 (14.31%) | 54 (25.71%) |
| Q5, no. (%) | 1750 (94.7%) | 7999 (98.79%) | 3393 (99.12%) | 208 (98.58%) |
| Very little | 138 (7.89%) | 315 (3.94%) | 130 (3.83%) | 4 (1.92%) |
| Little | 418 (23.89%) | 1119 (13.99%) | 324 (9.55%) | 25 (12.02%) |
| Medium | 610 (34.86%) | 3366 (42.08%) | 1044 (30.77%) | 42 (20.19%) |
| Much | 364 (20.8%) | 2598 (32.48%) | 1423 (41.94%) | 75 (36.06%) |
| Very much | 220 (12.57%) | 601 (7.51%) | 472 (13.91%) | 62 (29.81%) |
| Q6, no. (%) | 1750 (94.7%) | 7985 (98.62%) | 3390 (99.04%) | 210 (99.53%) |
| Very little | 149 (8.51%) | 312 (3.91%) | 140 (4.13%) | 6 (2.86%) |
| Little | 428 (24.46%) | 1142 (14.3%) | 328 (9.68%) | 26 (12.38%) |
| Medium | 593 (33.89%) | 3482 (43.61%) | 1098 (32.39%) | 50 (23.81%) |
| Much | 255 (14.57%) | 2482 (31.08%) | 1392 (41.06%) | 74 (35.24%) |
| Very much | 225 (12.86%) | 567 (7.1%) | 432 (12.74%) | 54 (25.71%) |
| Q7, no. (%) | 1701 (92.05%) | 6686 (82.57%) | 3319 (96.96%) | 204 (96.68%) |
| No use of AI independent of the disease. | 455 (26.75%) | 736 (11.01%) | 276 (8.32%) | 11 (5.39%) |
| Use only for minor illnesses (eg, cold). | 595 (34.98%) | 2316 (34.64%) | 678 (20.43%) | 45 (22.06%) |
| Use also for moderately severe diseases (eg, appendicitis). | 277 (16.28%) | 2251 (33.67%) | 911 (27.45%) | 33 (16.18%) |
| Use also for severe diseases (eg, cancer, traffic accidents). | 374 (21.99%) | 2482 (37.12%) | 1454 (43.81%) | 115 (56.37%) |
| Q8, no. (%) | 1745 (94.43%) | 7954 (98.23%) | 3383 (98.83%) | 210 (99.53%) |
| Completely disagree | 202 (11.58%) | 805 (10.12%) | 348 (10.29%) | 25 (11.9%) |
| Tend to disagree | 271 (15.53%) | 1674 (21.05%) | 605 (17.88%) | 38 (18.1%) |
| Neutral | 644 (36.91%) | 2247 (28.25%) | 706 (20.87%) | 27 (12.86%) |

**eTable 10. Absolute survey results for each item stratified by self-reported AI knowledge.**

| | No knowledge (N=1848) | Little knowledge (N=8097) | Good knowledge (N=3423) | Expert (N=211) |
|---|---|---|---|---|
| Tend to agree | 367 (21.03%) | 2622 (32.96%) | 1269 (37.51%) | 146 (69.52%) |
| Agree completely | 261 (14.96%) | 606 (%) | 455 (13.45%) | 46 (21.9%) |
| Q9, no. (%) | 1698 (91.88%) | 7642 (94.38%) | 3248 (94.89%) | 205 (97.16%) |
| Extremely negative | 90 (5.3%) | 188 (2.46%) | 88 (2.71%) | 6 (29.93%) |
| Rather negative | 161 (9.48%) | 799 (10.46%) | 224 (6.9%) | 9 (4.39%) |
| Neutral | 657 (38.69%) | 2336 (30.57%) | 614 (18.9%) | 25 (12.2%) |
| Rather positive | 459 (27.03%) | 3225 (32.2%) | 1490 (45.87%) | 122 (59.51%) |
| Extremely positive | 331 (19.49%) | 1094 (14.32%) | 832 (25.62%) | 103 (50.24%) |
| Q10, no. (%) | 1691 (91.5%) | 7630 (94.23%) | 3239 (94.62%) | 204 (96.68%) |
| Extremely negative | 113 (6.68%) | 305 (4%) | 117 (3.61%) | 10 (4.9%) |
| Rather negative | 296 (17.5%) | 959 (12.57%) | 295 (9.11%) | 15 (7.35%) |
| Neutral | 534 (31.58%) | 2398 (31.43%) | 721 (22.26%) | 32 (15.69%) |
| Rather positive | 430 (25.43%) | 2911 (38.15%) | 1433 (44.24%) | 65 (31.86%) |
| Extremely positive | 318 (18.81%) | 1057 (13.85%) | 673 (20.78%) | 82 (40.2%) |
| Q11, no. (%) | 1684 (91.13%) | 7639 (94.34%) | 3246 (94.83%) | 203 (96.21%) |
| Extremely negative | 76 (4.51%) | 173 (2.26%) | 81 (2.5%) | 8 (3.94%) |
| Rather negative | 132 (4.84%) | 513 (6.72%) | 170 (5.24%) | 18 (8.87%) |
| Neutral | 613 (36.4%) | 1842 (24.11%) | 461 (14.2%) | 19 (9.36%) |
| Rather positive | 500 (29.69%) | 3357 (43.95%) | 1393 (42.91%) | 50 (24.63%) |
| Extremely positive | 363 (21.56%) | 1754 (22.96%) | 1141 (35.15%) | 121 (59.61%) |
| Q12, no. (%) | 1611 (87.18%) | 7397 (91.35%) | 3182 (92.96%) | 201 (95.26%) |
| A high degree of accuracy, the decision path is clearly comprehensible (explainable AI). | 985 (61.14%) | 5155 (69.69%) | 2394 (75.24%) | 150 (74.63%) |
| A higher accuracy, the decision path however, is not comprehensible. | 626 (38.86%) | 2242 (30.31%) | 788 (24.76%) | 51 (25.37%) |
| Q13, no. (%) | 1584 (85.71%) | 7212 (89.07%) | 3118 (91.09%) | 194 (91.94%) |
| The AI misses almost no diagnosis, but often gives a false alarm. | 628 (39.65%) | 3362 (46.62%) | 1506 (48.3%) | 118 (60.82%) |
| The AI almost never gives a false alarm, but sometimes misses a diagnosis. | 631 (39.84%) | 2592 (35.94%) | 1131 (36.27%) | 58 (29.9%) |
| The AI gives a false alarm about as often as it misses a diagnosis. | 325 (20.52%) | 1258 (17.44%) | 481 (15.43%) | 18 (9.28%) |
| Q14, no. (%) | 1643 (88.91%) | 7484 (92.43%) | 3204 (93.6%) | 204 (96.68%) |
| Physicians make the diagnosis alone. | 246 (14.97%) | 424 (5.67%) | 150 (4.68%) | 5 (2.45%) |
| AI and physicians make the diagnosis together. Doctors make the final decision. | 968 (58.92%) | 5590 (74.69%) | 2422 (75.59%) | 153 (75%) |
| AI and physicians make the diagnosis | 205 (12.48%) | 1049 (14.02%) | 437 (13.64%) | 19 (9.31%) |

### eTable 10. Absolute survey results for each item stratified by self-reported AI knowledge.

| | No knowledge (N=1848) | Little knowledge (N=8097) | Good knowledge (N=3423) | Expert (N=211) |
|---|---|---|---|---|
| together. Both have equal authority. | | | | |
| AI and physicians make the diagnosis together. The AI makes the final decision. | 75 (4.56%) | 165 (2.2%) | 70 (2.18%) | 4 (1.96%) |
| AI makes the diagnosis alone. | 149 (9.07%) | 256 (3.42%) | 125 (3.9%) | 23 (11.27%) |
| Q15, no. (%) | 1616 (87.45%) | 7466 (92.21%) | 3186 (93.08%) | 202 (95.73%) |
| Extremely negative | 70 (4.33%) | 163 (2.18%) | 74 (2.32%) | 8 (3.96%) |
| Rather negative | 221 (13.68%) | 503 (6.74%) | 146 (4.58%) | 9 (4.46%) |
| Neutral | 578 (35.77%) | 2351 (31.49%) | 592 (18.58%) | 24 (11.88%) |
| Rather positive | 448 (27.72%) | 3196 (73.56%) | 1460 (45.83%) | 54 (26.73%) |
| Extremely positive | 299 (18.5%) | 1254 (39.24%) | 914 (28.69%) | 107 (52.97%) |
| Q16, no. (%) | 1601 (86.63%) | 7366 (90.97%) | 3156 (92.2%) | 201 (95.26%) |
| Always prefer facilities without AI. | 188 (11.74%) | 398 (5.4%) | 153 (4.85%) | 10 (4.98%) |
| Tend to prefer facilities without AI. | 438 (27.36%) | 1831 (24.86%) | 490 (15.53%) | 19 (9.45%) |
| Rather prefer facilities with AI. | 684 (42.72%) | 4230 (57.43%) | 1903 (60.3%) | 105 (52.24%) |
| Always prefer facilities with AI. | 291 (18.18%) | 907 (12.31%) | 610 (19.33%) | 67 (33.33%) |
| Q17, no. (%) | 1622 (87.77%) | 7472 (92.28%) | 3190 (93.19%) | 204 (96.68%) |
| Very worried | 363 (22.38%) | 1267 (16.96%) | 597 (18.71%) | 48 (23.53%) |
| Somewhat concerned | 427 (26.33%) | 2804 (37.53%) | 1062 (33.29%) | 67 (32.84%) |
| Neutral | 592 (36.5%) | 2143 (28.68%) | 777 (24.36%) | 39 (19.12%) |
| Rather unconcerned | 147 (9.06%) | 909 (12.17%) | 521 (16.33%) | 29 (14.22%) |
| Unconcerned | 93 (5.73%) | 349 (4.67%) | 233 (7.3%) | 21 (10.29%) |
| Q18, no. (%) | 1625 (87.93%) | 7470 (92.26%) | 3189 (93.16%) | 203 (96.21%) |
| Very worried | 470 (28.92%) | 1819 (24.35%) | 721 (22.61%) | 45 (22.17%) |
| Somewhat concerned | 555 (34.15%) | 2947 (39.45%) | 1077 (33.77%) | 69 (33.99%) |
| Neutral | 410 (25.23%) | 1714 (22.95%) | 762 (23.89%) | 38 (18.72%) |
| Rather unconcerned | 121 (7.45%) | 772 (10.33%) | 451 (14.14%) | 39 (19.21%) |
| Unconcerned | 69 (4.25%) | 219 (2.93%) | 178 (5.58%) | 12 (5.91%) |
| Q19, no. (%) | 1668 (90.26%) | 7512 (92.78%) | 3205 (93.63%) | 205 (97.16%) |
| Very worried | 563 (33.75%) | 2304 (30.67%) | 866 (27.02%) | 47 (22.93%) |
| Somewhat concerned | 474 (28.42%) | 2490 (33.15%) | 954 (29.77%) | 60 (29.27%) |
| Neutral | 393 (23.56%) | 1511 (20.11%) | 632 (19.72%) | 28 (13.66%) |
| Rather unconcerned | 152 (9.11%) | 833 (11.09%) | 522 (16.29%) | 44 (21.46%) |
| Unconcerned | 86 (5.16%) | 374 (4.98%) | 231 (7.21%) | 26 (12.68%) |
| Q20, no. (%) | 1662 (89.94%) | 7505 (92.69%) | 3203 (93.57%) | 201 (95.26%) |
| Very worried | 537 (32.31%) | 1953 (26.02%) | 713 (22.26%) | 48 (23.88%) |
| Somewhat concerned | 503 (30.26%) | 2478 (33.02%) | 907 (28.32%) | 56 (27.86%) |
| Neutral | 444 (26.71%) | 2009 (26.77%) | 383 (11.96%) | 33 (16.42%) |
| Rather unconcerned | 111 (6.68%) | 734 (9.78%) | 467 (14.58%) | 26 (12.94%) |
| Unconcerned | 67 (4.03%) | 331 (4.41%) | 378 (11.8%) | 28 (13.9%) |

Abbreviations: Q1, What are your general views on the use of artificial intelligence (AI) in medicine?; Q2, Artificial intelligence (AI) should be increasingly used in the healthcare sector.; Q3, How much confidence do you have that AI can improve healthcare?; Q4, How much do you trust an AI to provide reliable information about your health?; Q5, How much do you trust an

AI to provide accurate information about your diagnosis?; Q6, How much do you trust an AI to provide accurate information about your response to therapy?; Q7, Which of the following statements about a potential application of AI in medicine do you most likely agree with?; Q8, I would trust a highly accurate AI to make a vital decision for me.; Q9, How would you rate it if a certified AI software analyzes X-ray images?; Q10, How would you rate it if a certified AI software diagnoses cancer?; Q11, How would you rate it if a certified AI software would be available to doctors as a second opinion?; Q12/Q13, Suppose an AI makes a diagnosis. What would you prefer?; Q14, Suppose an AI has about the same accuracy as doctors. Which situation for a diagnosis would you prefer?; Q15, What do you think of healthcare facilities (clinics, practices) using AI software to aid in diagnosis?; Q16, Would you prefer to visit healthcare facilities that use AI software? Q17, How concerned are you about AI compromising the protection of your personal data?; Q18, How concerned are you that the use of AI will reduce the contact between physicians and patients?; Q19, How concerned are you that AI could replace human doctors in the future?; Q20, How concerned are you that the use of AI will lead to higher healthcare costs?.

### eTable 11. Absolute survey results for each item stratified by median age and median number of technical devices used weekly.

| | Age ≤ median of 48 years (N=6287) | Age > median of 48 years (N=6165) | Technical devices used ≤ median of two (N=9611) | Technical devices > median of two (N=4141) |
|---|---|---|---|---|
| Q1, no. (%) | 6171 (98.15%) | 6049 (98.12%) | 9390 (97.7%) | 4105 (99.13%) |
| Extremely negative | 158 (2.56%) | 117 (1.93%) | 220 (2.34%) | 76 (1.85%) |
| Rather negative | 441 (7.15%) | 479 (7.92%) | 793 (8.45%) | 238 (5.8%) |
| Neutral | 1747 (28.31%) | 2245 (37.11%) | 3369 (35.88%) | 1025 (24.97%) |
| Rather positive | 2622 (42.49%) | 2431 (40.19%) | 3592 (38.25%) | 1962 (47.8%) |
| Extremely positive | 1203 (19.49%) | 777 (12.85%) | 1416 (15.08%) | 804 (19.59%) |
| Q2, no. (%) | 6147 (97.77%) | 5936 (96.29%) | 9245 (96.19%) | 4062 (98.09%) |
| Completely disagree | 232 (3.77%) | 158 (2.66%) | 318 (3.44%) | 110 (2.71%) |
| Tend to disagree | 508 (8.26%) | 468 (7.88%) | 797 (8.62%) | 261 (6.43%) |
| Neutral | 1308 (21.28%) | 1825 (30.74%) | 2606 (28.19%) | 835 (10.56%) |
| Tend to agree | 2420 (39.37%) | 2266 (38.17%) | 3393 (36.7%) | 1169 (28.78%) |
| Agree completely | 1679 (27.31%) | 1219 (20.54%) | 2131 (23.05%) | 1087 (26.76%) |
| Q3, no. (%) | 6232 (99.13%) | 6038 (97.94%) | 9439 (98.21%) | 4097 (98.94%) |
| Very little | 244 (3.92%) | 230 (3.81%) | 415 (4.4%) | 113 (2.76%) |
| Little | 692 (11.1%) | 731 (12.11%) | 1201 (12.72%) | 343 (8.37%) |
| Medium | 2035 (32.65%) | 2404 (39.81%) | 3593 (38.07%) | 1300 (31.73%) |
| Much | 2233 (35.83%) | 1937 (32.08%) | 2941 (31.16%) | 1625 (39.66%) |
| Very much | 1028 (16.5%) | 736 (12.19%) | 1289 (13.66%) | 716 (17.48%) |
| Q4, no. (%) | 6225 (99.01%) | 6013 (97.53%) | 9401 (97.82%) | 4100 (99.01%) |
| Very little | 269 (4.32%) | 243 (4.04%) | 454 (4.83%) | 110 (2.68%) |
| Little | 785 (12.61%) | 930 (15.47%) | 1392 (14.81%) | 466 (11.37%) |
| Medium | 2314 (37.17%) | 2331 (38.77%) | 3673 (39.07%) | 1473 (35.93%) |
| Much | 2182 (35.05%) | 2025 (33.68%) | 2967 (31.56%) | 1620 (39.51%) |
| Very much | 675 (10.84%) | 484 (8.05%) | 915 (9.73%) | 431 (10.51%) |
| Q5, no. (%) | 6225 (99.01%) | 6009 (97.47%) | 9398 (97.78%) | 4091 (98.79%) |
| Very little | 267 (4.29%) | 273 (4.45%) | 459 (4.88%) | 137 (3.35%) |
| Little | 837 (13.45%) | 928 (15.44%) | 1448 (15.41%) | 470 (11.49%) |
| Medium | 2330 (37.43%) | 2285 (38.03%) | 3606 (38.37%) | 1484 (36.27%) |
| Much | 2127 (34.17%) | 2008 (33.42%) | 2927 (31.14%) | 1589 (38.84%) |
| Very much | 664 (10.67%) | 515 (8.57%) | 958 (10.19%) | 411 (10.05%) |
| Q6, no. (%) | 6221 (98.95%) | 5997 (97.27%) | 9385 (97.65%) | 4088 (98.72%) |
| Very little | 271 (4.36%) | 292 (4.87%) | 470 (5.01%) | 143 (3.5%) |
| Little | 838 (13.47%) | 952 (15.87%) | 1441 (15.35%) | 520 (12.72%) |
| Medium | 2416 (38.84%) | 2366 (39.45%) | 3758 (40.04%) | 1507 (36.86%) |
| Much | 2060 (33.11%) | 1907 (31.8%) | 2797 (29.8%) | 1547 (37.84%) |
| Very much | 636 (10.22%) | 480 (8%) | 919 (9.79%) | 371 (9.08%) |
| Q7, no. (%) | 6114 (97.25%) | 5817 (94.36%) | 9149 (95.19%) | 3985 (96.23%) |
| No use of AI independent of the disease. | 665 (10.88%) | 669 (11.5%) | 1239 (13.54%) | 259 (6.5%) |
| Use only for minor illnesses (eg, cold). | 1797 (29.39%) | 1571 (27.01%) | 2675 (26.24%) | 684 (17.16%) |
| Use also for moderately severe diseases (eg, appendicitis). | 1652 (27.02%) | 1510 (25.96%) | 2415 (26.4%) | 1095 (27.48%) |
| Use also for severe diseases (eg, cancer, traffic accidents). | 2000 (32.71%) | 2067 (35.53%) | 2820 (30.82%) | 1647 (41.33%) |
| Q8, no. (%) | 6206 (98.71%) | 5979 (96.98%) | 9348 (97.26%) | 4083 (98.6%) |
| Completely disagree | 640 (10.31%) | 657 (10.99%) | 958 (10.25%) | 452 (11.07%) |

**eTable 11. Absolute survey results for each item stratified by median age and median number of technical devices used weekly.**

| | Age ≤ median of 48 years (N=6287) | Age > median of 48 years (N=6165) | Technical devices used ≤ median of two (N=9611) | Technical devices > median of two (N=4141) |
|---|---|---|---|---|
| Tend to disagree | 1252 (20.17%) | 1111 (18.58%) | 1718 (18.38%) | 891 (21.82%) |
| Neutral | 1520 (25.49%) | 1816 (30.37%) | 2717 (29.07%) | 939 (23%) |
| Tend to agree | 2097 (33.79%) | 1869 (31.26%) | 2913 (31.16%) | 1460 (35.76%) |
| Agree completely | 697 (11.23%) | 526 (8.8%) | 1042 (11.15%) | 341 (8.35%) |
| Q9, no. (%) | 6133 (97.55%) | 5682 (92.17%) | 9020 (93.85%) | 3923 (94.74%) |
| Extremely negative | 204 (3.33%) | 151 (2.66%) | 294 (3.26%) | 84 (2.14%) |
| Rather negative | 607 (9.9%) | 521 (9.17%) | 899 (9.97%) | 322 (8.21%) |
| Neutral | 1536 (25.04%) | 1802 (31.71%) | 2760 (30.6%) | 909 (23.17%) |
| Rather positive | 2522 (41.12%) | 2320 (40.83%) | 3467 (38.44%) | 1821 (46.42%) |
| Extremely positive | 1264 (20.61%) | 888 (15.63%) | 1600 (17.74%) | 787 (20.06%) |
| Q10, no. (%) | 6125 (97.42%) | 5664 (91.87%) | 8985 (93.49%) | 3925 (94.78%) |
| Extremely negative | 286 (4.67%) | 232 (4.1%) | 441 (4.91%) | 121 (3.08%) |
| Rather negative | 741 (12.1%) | 708 (12.5%) | 1158 (12.89%) | 419 (10.68%) |
| Neutral | 1710 (27.92%) | 1673 (29.54%) | 2715 (30.22%) | 1009 (25.71%) |
| Rather positive | 2295 (37.47%) | 2178 (38.45%) | 3183 (35.43%) | 1709 (43.54%) |
| Extremely positive | 1093 (17.84%) | 873 (15.41%) | 1488 (16.56%) | 667 (16.99%) |
| Q11, no. (%) | 6119 (97.33%) | 5676 (92.07%) | 8989 (93.53%) | 3929 (94.88%) |
| Extremely negative | 192 (3.14%) | 124 (2.18%) | 265 (2.95%) | 77 (1.96%) |
| Rather negative | 462 (7.55%) | 303 (4.34%) | 629 (7%) | 204 (5.19%) |
| Neutral | 1303 (21.29%) | 1387 (24.44%) | 2346 (26.1%) | 617 (15.7%) |
| Rather positive | 2450 (40.04%) | 2453 (43.22%) | 3623 (40.3%) | 1741 (44.31%) |
| Extremely positive | 1712 (27.98%) | 1409 (24.82%) | 2126 (23.65%) | 1290 (32.83%) |
| Q12, no. (%) | 6014 (95.66%) | 5463 (88.61%) | 8691 (90.43%) | 3834 (92.59%) |
| A high degree of accuracy, the decision path is clearly comprehensible (explainable AI). | 4327 (71.95%) | 3702 (67.76%) | 5892 (67.79%) | 2897 (75.56%) |
| A higher accuracy, the decision path however, is not comprehensible. | 1687 (28.05%) | 1761 (32.24%) | 2799 (32.21%) | 937 (24.44%) |
| Q13, no. (%) | 5948 (94.61%) | 5286 (85.74%) | 8483 (88.26%) | 3753 (90.63%) |
| The AI misses almost no diagnosis, but often gives a false alarm. | 2885 (48.5%) | 2358 (44.61%) | 3719 (43.84%) | 1952 (52.01%) |
| The AI almost never gives a false alarm, but sometimes misses a diagnosis. | 2079 (34.95%) | 1965 (37.17%) | 3224 (38.01%) | 1199 (31.95%) |
| The AI gives a false alarm about as often as it misses a diagnosis. | 984 (16.54%) | 963 (18.22%) | 1540 (18.15%) | 568 (15.13%) |
| Q14, no. (%) | 5939 (94.46%) | 5579 (90.49%) | 8752 (91.06%) | 3861 (93.24%) |
| Physicians make the diagnosis alone. | 337 (5.67%) | 420 (7.53%) | 678 (7.75%) | 149 (3.86%) |
| AI and physicians make the diagnosis together. Doctors make the final decision. | 4290 (72.23%) | 4159 (74.55%) | 6107 (69.78%) | 3082 (79.82%) |

**eTable 11. Absolute survey results for each item stratified by median age and median number of technical devices used weekly.**

| | **Age ≤ median of 48 years (N=6287)** | **Age > median of 48 years (N=6165)** | **Technical devices used ≤ median of two (N=9611)** | **Technical devices > median of two (N=4141)** |
|---|---|---|---|---|
| AI and physicians make the diagnosis together. Both have equal authority. | 886 (14.92%) | 680 (12.19%) | 1254 (14.33%) | 464 (12.02%) |
| AI and physicians make the diagnosis together. The AI makes the final decision. | 139 (2.34%) | 439 (7.87%) | 265 (3.03%) | 52 (1.35%) |
| AI makes the diagnosis alone. | 287 (4.83%) | 181 (3.24%) | 448 (5.12%) | 114 (2.95%) |
| Q15, no. (%) | 5940 (94.48%) | 5572 (90.38%) | 8761 (91.16%) | 3854 (93.07%) |
| Extremely negative | 181 (3.05%) | 122 (2.19%) | 259 (2.96%) | 61 (1.58%) |
| Rather negative | 384 (6.46%) | 445 (7.99%) | 701 (8%) | 188 (4.88%) |
| Neutral | 1518 (25.56%) | 1720 (30.87%) | 2733 (31.2%) | 853 (22.13%) |
| Rather positive | 2472 (41.62%) | 2300 (41.28%) | 3390 (38.69%) | 1820 (47.22%) |
| Extremely positive | 1385 (23.32%) | 985 (17.68%) | 1677 (19.14%) | 932 (24.18%) |
| Q16, no. (%) | 5890 (93.69%) | 5504 (89.28%) | 8653 (90.03%) | 3807 (91.93%) |
| Always prefer facilities without AI. | 364 (6.18%) | 354 (6.43%) | 620 (7.17%) | 152 (3.99%) |
| Tend to prefer facilities without AI. | 1187 (20.15%) | 1368 (24.85%) | 2130 (24.62%) | 660 (17.34%) |
| Rather prefer facilities with AI. | 3361 (57.06%) | 3088 (56.1%) | 4559 (52.69%) | 2445 (64.22%) |
| Always prefer facilities with AI. | 978 (16.6%) | 694 (12.61%) | 1344 (15.53%) | 550 (14.45%) |
| Q17, no. (%) | 5938 (94.45%) | 5593 (90.72%) | 8772 (91.27%) | 3859 (93.19%) |
| Very worried | 1094 (18.42%) | 1022 (18.27%) | 1672 (19.06%) | 644 (16.69%) |
| Somewhat concerned | 2073 (34.91%) | 1962 (35.08%) | 3042 (34.68%) | 1360 (35.24%) |
| Neutral | 1630 (27.45%) | 1623 (29.02%) | 2605 (29.7%) | 983 (25.47%) |
| Rather unconcerned | 779 (13.12%) | 698 (12.48%) | 994 (11.33%) | 623 (16.14%) |
| Unconcerned | 362 (6.1%) | 288 (5.15%) | 459 (5.23%) | 249 (6.45%) |
| Q18, no. (%) | 5940 (94.48%) | 5592 (90.71%) | 8771 (91.26%) | 3860 (93.21%) |
| Very worried | 1334 (22.46%) | 1530 (27.36%) | 2229 (25.41%) | 880 (22.8%) |
| Somewhat concerned | 2078 (34.98%) | 2201 (39.36%) | 3227 (36.79%) | 1462 (37.88%) |
| Neutral | 1503 (25.3%) | 1167 (20.87%) | 2115 (24.11%) | 838 (21.71%) |
| Rather unconcerned | 730 (12.29%) | 543 (9.71%) | 880 (10.03%) | 512 (13.26%) |
| Unconcerned | 295 (4.97%) | 151 (2.7%) | 320 (3.65%) | 168 (4.35%) |
| Q19, no. (%) | 6037 (96.02%) | 5600 (90.84%) | 8868 (92.27%) | 3866 (93.36%) |
| Very worried | 1597 (26.45%) | 1921 (34.3%) | 2764 (31.17%) | 1088 (28.14%) |
| Somewhat concerned | 1850 (30.64%) | 1838 (32.82%) | 2763 (31.16%) | 1244 (32.18%) |
| Neutral | 1327 (21.98%) | 1033 (18.45%) | 1900 (21.43%) | 688 (17.80%) |
| Rather unconcerned | 833 (13.8%) | 585 (10.45%) | 982 (11.07%) | 585 (15.13%) |
| Unconcerned | 430 (7.12%) | 223 (3.98%) | 459 (5.18%) | 261 (6.75%) |
| Q20, no. (%) | 6030 (95.91%) | 5589 (90.66%) | 8851 (92.09%) | 3862 (93.26%) |
| Very worried | 1518 (25.17%) | 1545 (27.64%) | 2436 (27.52%) | 886 (22.94%) |
| Somewhat concerned | 1883 (31.23%) | 1794 (32.1%) | 5690 (64.29%) | 1133 (29.34%) |
| Neutral | 1486 (24.64%) | 1517 (27.14%) | 2340 (26.44%) | 1005 (26.02%) |

### eTable 11. Absolute survey results for each item stratified by median age and median number of technical devices used weekly.

| | Age ≤ median of 48 years (N=6287) | Age > median of 48 years (N=6165) | Technical devices used ≤ median of two (N=9611) | Technical devices > median of two (N=4141) |
|---|---|---|---|---|
| Rather unconcerned | 725 (12.02%) | 503 (9%) | 800 (9.04%) | 557 (14.42%) |
| Unconcerned | 418 (6.93%) | 230 (4.12%) | 429 (4.85%) | 281 (7.28%) |

Abbreviations: Q1, What are your general views on the use of artificial intelligence (AI) in medicine?; Q2, Artificial intelligence (AI) should be increasingly used in the healthcare sector.; Q3, How much confidence do you have that AI can improve healthcare?; Q4, How much do you trust an AI to provide reliable information about your health?; Q5, How much do you trust an AI to provide accurate information about your diagnosis?; Q6, How much do you trust an AI to provide accurate information about your response to therapy?; Q7, Which of the following statements about a potential application of AI in medicine do you most likely agree with?; Q8, I would trust a highly accurate AI to make a vital decision for me.; Q9, How would you rate it if a certified AI software analyzes X-ray images?; Q10, How would you rate it if a certified AI software diagnoses cancer?; Q11, How would you rate it if a certified AI software would be available to doctors as a second opinion?; Q12/Q13, Suppose an AI makes a diagnosis. What would you prefer?; Q14, Suppose an AI has about the same accuracy as doctors. Which situation for a diagnosis would you prefer?; Q15, What do you think of healthcare facilities (clinics, practices) using AI software to aid in diagnosis?; Q16, Would you prefer to visit healthcare facilities that use AI software? Q17, How concerned are you about AI compromising the protection of your personal data?; Q18, How concerned are you that the use of AI will reduce the contact between physicians and patients?; Q19, How concerned are you that AI could replace human doctors in the future?; Q20, How concerned are you that the use of AI will lead to higher healthcare costs?.

**eTable 12. Absolute survey results for each item stratified by highest educational level.**

| | Elementary School (N=1192) | Middle School (N=2844) | High School (N=4142) | University (N=5403) |
|---|---|---|---|---|
| Q1, no. (%) | 1151 (96.56%) | 2767 (97.29%) | 4058 (97.97%) | 5368 (99.35%) |
| Extremely negative | 36 (3.13%) | 67 (2.42%) | 617 (15.2%) | 122 (2.27%) |
| Rather negative | 91 (7.91%) | 253 (9.14%) | 1729 (42.61%) | 333 (6.2%) |
| Neutral | 496 (43.49%) | 1074 (38.81%) | 1304 (32.13%) | 1454 (27.09%) |
| Rather positive | 357 (31.02%) | 1005 (36.32%) | 1729 (42.61%) | 2416 (45.01%) |
| Extremely positive | 171 (14.86%) | 368 (13.3%) | 617 (15.2%) | 1043 (19.43%) |
| Q2, no. (%) | 1095 (91.86%) | 2697 (94.83%) | 4055 (97.9%) | 5317 (98.41%) |
| Completely disagree | 53 (4.84%) | 95 (3.52%) | 111 (2.74%) | 159 (2.99%) |
| Tend to disagree | 82 (7.49%) | 257 (9.53%) | 352 (8.68%) | 358 (6.73%) |
| Neutral | 377 (34.43%) | 838 (31.07%) | 1029 (25.38%) | 1151 (21.65%) |
| Tend to agree | 333 (30.41%) | 965 (35.78%) | 1588 (39.16%) | 2225 (41.85%) |
| Agree completely | 250 (22.83%) | 542 (20.1%) | 975 (24.04%) | 1424 (26.78%) |
| Q3, no. (%) | 1084 (90.94%) | 2793 (98.21%) | 4091 (98.77%) | 5354 (99.09%) |
| Very little | 70 (6.46%) | 114 (4.08%) | 164 (4.01%) | 168 (3.14%) |
| Little | 156 (14.39%) | 395 (14.14%) | 503 (12.3%) | 468 (8.74%) |
| Medium | 454 (41.88%) | 1109 (39.71%) | 1467 (35.86%) | 1805 (33.71%) |
| Much | 293 (27.03%) | 853 (30.54%) | 1463 (35.76%) | 1316 (24.58%) |
| Very much | 172 (15.07%) | 322 (11.53%) | 464 (11.34%) | 997 (18.62%) |
| Q4, no. (%) | 1140 (95.64%) | 2775 (97.57%) | 4085 (98.62%) | 5353 (99.07%) |
| Very little | 72 (6.32%) | 145 (5.23%) | 170 (4.16%) | 165 (3.08%) |
| Little | 173 (15.18%) | 396 (14.27%) | 566 (13.86%) | 698 (13.04%) |
| Medium | 474 (41.58%) | 1190 (42.88%) | 1517 (37.14%) | 1900 (35.49%) |
| Much | 286 (25.09%) | 825 (29.73%) | 1501 (36.74%) | 1946 (36.35%) |
| Very much | 135 (11.84%) | 219 (7.89%) | 331 (8.1%) | 644 (12.03%) |
| Q5, no. (%) | 1139 (95.55%) | 2771 (97.43%) | 4086 (98.65%) | 5347 (98.96%) |
| Very little | 76 (6,67%) | 136 (4.91%) | 179 (4.38%) | 187 (3.5%) |
| Little | 172 (15.1%) | 414 (14.94%) | 607 (14.86%) | 704 (13.17%) |
| Medium | 455 (39.95%) | 1174 (42.37%) | 1496 (36.61%) | 1910 (35.72%) |
| Much | 287 (25.2%) | 808 (29.16%) | 1484 (36.32%) | 1903 (35.59%) |
| Very much | 149 (13.08%) | 239 (8.63%) | 320 (7.83%) | 643 (12.03%) |
| Q6, no. (%) | 1132 (94.97%) | 2774 (97.54%) | 4080 (98.5%) | 5342 (98.87%) |
| Very little | 67 (5.92%) | 152 (5.48%) | 172 (4.22%) | 205 (3.84%) |
| Little | 174 (15.37%) | 409 (14.74%) | 630 (15.44%) | 727 (13.81%) |
| Medium | 460 (40.64%) | 1193 (43.01%) | 1586 (38.87%) | 1966 (36.8%) |
| Much | 301 (26.59%) | 777 (28.01%) | 1382 (33.87%) | 1850 (34.63%) |
| Very much | 130 (11.48%) | 241 (8.69%) | 310 (7.6%) | 594 (11.12%) |
| Q7, no. (%) | 1090 (91.44%) | 2697 (94.83%) | 3996 (96.48%) | 5224 (96.69%) |
| No use of AI independent of the disease. | 242 (22.2%) | 378 (4.02%) | 437 (10.94%) | 421 (8.06%) |
| Use only for minor illnesses (eg, cold). | 314 (28.81%) | 809 (30%) | 1123 (28.1%) | 1379 (26.4%) |
| Use also for moderately severe diseases (eg, appendicitis). | 149 (13.67%) | 723 (26.81%) | 1125 (28.15%) | 1366 (26.15%) |
| Use also for severe diseases (eg, cancer, traffic accidents). | 285 (26.15%) | 787 (29.18%) | 1301 (32.56%) | 2058 (39.4%) |
| Q8, no. (%) | 1128 (94.63%) | 2758 (96.98%) | 4079 (98.48%) | 5322 (%) |
| Completely disagree | 138 (12.23%) | 323 (11.71%) | 427 (10.47%) | 501 (9.41%) |
| Tend to disagree | 160 (14.18%) | 577 (20.92%) | 825 (20.23%) | 1018 (19.13%) |
| Neutral | 373 (33.07%) | 785 (28.46%) | 1097 (26.89%) | 1367 (25.69%) |
| Tend to agree | 318 (28.19%) | 776 (28.14%) | 1359 (33.32%) | 1881 (35.34%) |

**eTable 12. Absolute survey results for each item stratified by highest educational level.**

| | Elementary School (N=1192) | Middle School (N=2844) | High School (N=4142) | University (N=5403) |
|---|---|---|---|---|
| Agree completely | 139 (12.32%) | 297 (10.77%) | 371 (9.1%) | 555 (10.43%) |
| Q9, no. (%) | 1061 (89.01%) | 2632 (92.55%) | 4001 (96.6%) | 5102 (94.43%) |
| Extremely negative | 45 (4.24%) | 86 (3.27%) | 114 (2.85%) | 121 (2.37%) |
| Rather negative | 104 (9.8%) | 350 (13.3%) | 368 (9.2%) | 391 (7.66%) |
| Neutral | 368 (34.68%) | 830 (31.53%) | 1152 (28.79%) | 1266 (24.81%) |
| Rather positive | 342 (32.23%) | 347 (13.18%) | 1701 (42.51%) | 2249 (44.08%) |
| Extremely positive | 202 (19.04%) | 419 (15.92%) | 666 (16.65%) | 1075 (21.07%) |
| Q10, no. (%) | 1058 (88.76%) | 2620 (92.12%) | 3994 (96.43%) | 5092 (94.24%) |
| Extremely negative | 56 (5.29%) | 680 (25.95%) | 175 (4.38%) | 183 (3.59%) |
| Rather negative | 116 (10.96%) | 382 (14.58%) | 444 (11.12%) | 620 (12.18%) |
| Neutral | 362 (34.22%) | 806 (30.76%) | 1165 (29.17%) | 1346 (26.43%) |
| Rather positive | 337 (31.85%) | 911 (34.77%) | 1560 (39.06%) | 2037 (40%) |
| Extremely positive | 187 (17.67%) | 385 (14.69%) | 650 (16.27%) | 906 (17.79%) |
| Q11, no. (%) | 1052 (88.26%) | 2623 (92.23%) | 3991 (96.35%) | 5106 (94.5%) |
| Extremely negative | 38 (3.61%) | 66 (2.52%) | 97 (2.43%) | 133 (2.6%) |
| Rather negative | 59 (5.61%) | 186 (7.09%) | 283 (7.09%) | 298 (5.84%) |
| Neutral | 320 (30.42%) | 727 (27.72%) | 875 (21.92%) | 998 (19.55%) |
| Rather positive | 405 (38.5%) | 1082 (41.25%) | 1741 (43.62%) | 2083 (40.8%) |
| Extremely positive | 230 (21.86%) | 562 (21.43%) | 995 (24.93%) | 1594 (31.22%) |
| Q12, no. (%) | 1004 (84.23%) | 2493 (87.66%) | 3885 (93.89%) | 5010 (92.73%) |
| A high degree of accuracy, the decision path is clearly comprehensible (explainable AI). | 666 (66.33%) | 1743 (69.92%) | 2773 (71.38%) | 3524 (70.34%) |
| A higher accuracy, the decision path however, is not comprehensible. | 338 (33.67%) | 750 (30.08%) | 1112 (28.62%) | 1486 (29.66%) |
| Q13, no. (%) | 958 (80.37%) | 2441 (85.83%) | 3816 (92.13%) | 4900 (90.69%) |
| The AI misses almost no diagnosis, but often gives a false alarm. | 393 (41.02%) | 1110 (45.47%) | 1749 (45.83%) | 2381 (48.59%) |
| The AI almost never gives a false alarm, but sometimes misses a diagnosis. | 375 (39.14%) | 897 (36.75%) | 1376 (36.06%) | 1751 (35.73%) |
| The AI gives a false alarm about as often as it misses a diagnosis. | 190 (19.83%) | 434 (17.78%) | 691 (18.11%) | 768 (15.67%) |
| Q14, no. (%) | 1028 (86.24%) | 2537 (89.21%) | 3888 (93.87%) | 5017 (92.86%) |
| Physicians make the diagnosis alone. | 82 (7.98%) | 134 (5.28%) | 252 (6.48%) | 212 (4.23%) |
| AI and physicians make the diagnosis together. Doctors make the final decision. | 632 (61.48%) | 1731 (68.23%) | 2818 (72.48%) | 3917 (78.07%) |
| AI and physicians make the diagnosis together. Both have equal authority. | 161 (15.66%) | 351 (13.84%) | 598 (15.38%) | 589 (11.74%) |
| AI and physicians make the diagnosis together. | 39 (3.79%) | 84 (3.31%) | 92 (2.37%) | 94 (1.87%) |

**eTable 12. Absolute survey results for each item stratified by highest educational level.**

| | Elementary School (N=1192) | Middle School (N=2844) | High School (N=4142) | University (N=5403) |
|---|---|---|---|---|
| The AI makes the final decision. | | | | |
| AI makes the diagnosis alone. | 114 (11.09%) | 237 (9.34%) | 128 (3.29%) | 205 (4.09%) |
| Q15, no. (%) | 1033 (86.66%) | 2563 (90.12%) | 3895 (94.04%) | 4982 (92.21%) |
| Extremely negative | 33 (3.19%) | 63 (2.46%) | 101 (2.59%) | 118 (2.37%) |
| Rather negative | 51 (4.94%) | 222 (8.66%) | 232 (5.96%) | 372 (7.47%) |
| Neutral | 406 (39.3%) | 918 (35.82%) | 1130 (29.01%) | 1077 (21.62%) |
| Rather positive | 337 (32.62%) | 923 (36.01%) | 1693 (43.47%) | 2208 (44.32%) |
| Extremely positive | 206 (19.94%) | 437 (17.05%) | 739 (18.97%) | 1207 (24.23%) |
| Q16, no. (%) | 1009 (84.65%) | 2538 (89.24%) | 3843 (92.78%) | 4941 (91.45%) |
| Always prefer facilities without AI. | 102 (10.11%) | 203 (8%) | 226 (5.88%) | 228 (4.61%) |
| Tend to prefer facilities without AI. | 267 (26.46%) | 729 (28.72%) | 905 (23.55%) | 861 (17.43%) |
| Rather prefer facilities with AI. | 455 (45.09%) | 1248 (49.17%) | 2204 (57.35%) | 3027 (61.26%) |
| Always prefer facilities with AI. | 185 (18.33%) | 358 (14.11%) | 508 (13.22%) | 825 (16.7%) |
| Q17, no. (%) | 1032 (86.58%) | 2559 (89.98%) | 3907 (94.33%) | 4991 (92.37%) |
| Very worried | 190 (18.41%) | 434 (16.96%) | 802 (20.53%) | 855 (17.13%) |
| Somewhat concerned | 318 (30.81%) | 875 (34.19%) | 1424 (36.45%) | 1739 (34.84%) |
| Neutral | 350 (33.91%) | 751 (29.35%) | 1038 (26.57%) | 1411 (28.27%) |
| Rather unconcerned | 108 (10.47%) | 335 (13.09%) | 445 (11.39%) | 714 (14.31%) |
| Unconcerned | 66 (6.4%) | 164 (6.41%) | 198 (5.07%) | 272 (5.45%) |
| Q18, no. (%) | 1030 (86.41%) | 2565 (90.19%) | 3902 (94.21%) | 4994 (92.43%) |
| Very worried | 249 (24.17%) | 635 (24.76%) | 1099 (28.17%) | 1076 (21.55%) |
| Somewhat concerned | 352 (34.17%) | 806 (31.42%) | 1420 (36.39%) | 1960 (39.25%) |
| Neutral | 279 (27.09%) | 640 (24.95%) | 872 (22.35%) | 1135 (22.73%) |
| Rather unconcerned | 102 (9.9%) | 266 (10.37%) | 389 (9.97%) | 627 (12.56%) |
| Unconcerned | 49 (4.76%) | 116 (4.52%) | 122 (3.13%) | 196 (3.92%) |
| Q19, no. (%) | 1039 (87.16%) | 2580 (90.72%) | 3950 (95.36%) | 5024 (92.99%) |
| Very worried | 341 (32.82%) | 815 (31.59%) | 1367 (34.61%) | 1277 (25.42%) |
| Somewhat concerned | 300 (28.87%) | 748 (28.99%) | 1251 (31.67%) | 1659 (33.02%) |
| Neutral | 235 (22.62%) | 549 (21.28%) | 793 (20.08%) | 971 (19.33%) |
| Rather unconcerned | 111 (10.68%) | 295 (11.43%) | 372 (9.42%) | 775 (14.43%) |
| Unconcerned | 52 (5%) | 153 (5.93%) | 167 (4.23%) | 342 (6.81%) |
| Q20, no. (%) | 1038 (87.08%) | 2582 (90.79%) | 3951 (95.39%) | 5002 (92.58%) |
| Very worried | 302 (29.09%) | 670 (25.95%) | 1225 (31%) | 1076 (21.51%) |
| Somewhat concerned | 284 (27.36%) | 758 (29.36%) | 1253 (31.71%) | 1640 (32.79%) |
| Neutral | 301 (29%) | 747 (28.93%) | 977 (24.73%) | 1289 (25.77%) |
| Rather unconcerned | 99 (9.54%) | 271 (10.5%) | 338 (8.55%) | 641 (12.81%) |
| Unconcerned | 52 (5.01%) | 136 (5.27%) | 158 (4%) | 356 (7.12%) |

Abbreviations: Q1, What are your general views on the use of artificial intelligence (AI) in medicine?; Q2, Artificial intelligence (AI) should be increasingly used in the healthcare sector.; Q3, How much confidence do you have that AI can improve healthcare?; Q4, How much do you trust an AI to provide reliable information about your health?; Q5, How much do you trust an AI to provide accurate information about your diagnosis?; Q6, How much do you trust an AI to provide accurate information about your response to therapy?; Q7, Which of the following statements about a potential application of AI in medicine do you most likely agree with?; Q8, I would trust a highly accurate AI to make a vital decision for me.; Q9, How would you rate it if a certified AI software analyzes X-ray images?; Q10, How would you rate it if a certified AI software diagnoses cancer?; Q11, How would you rate it if a certified AI software would be available to doctors as a second opinion?; Q12/Q13, Suppose an AI makes a diagnosis. What would you prefer?; Q14, Suppose an AI has about the same accuracy as doctors. Which situation for a diagnosis would you prefer?; Q15, What do you think of healthcare facilities (clinics, practices) using AI software to aid in diagnosis?; Q16, Would you prefer to visit healthcare facilities that use AI software? Q17, How concerned are you about AI compromising the protection of your personal data?; Q18, How concerned are you that the use of AI will reduce the contact between physicians and patients?; Q19, How concerned are you that AI could replace human doctors in the future?; Q20, How concerned are you that the use of AI will lead to higher healthcare costs?.

### eTable 13. STROBE statement.

| | Item No | Recommendation | Location |
|---|---|---|---|
| **Title and abstract** | 1 | (*a*) Indicate the study's design with a commonly used term in the title or the abstract | Page 1 |
| | | (*b*) Provide in the abstract an informative and balanced summary of what was done and what was found | Page 4 |
| **Introduction** | | | |
| Background/rationale | 2 | Explain the scientific background and rationale for the investigation being reported | Page 5 |
| Objectives | 3 | State specific objectives, including any prespecified hypotheses | Page 5 |
| **Methods** | | | |
| Study design | 4 | Present key elements of study design early in the paper | Pages 6-7 |
| Setting | 5 | Describe the setting, locations, and relevant dates, including periods of recruitment, exposure, follow-up, and data collection | Page 6 |
| Participants | 6 | (*a*) Give the eligibility criteria, and the sources and methods of selection of participants | Page 6 |
| Variables | 7 | Clearly define all outcomes, exposures, predictors, potential confounders, and effect modifiers. Give diagnostic criteria, if applicable | Pages 7-8 |
| Data sources/ measurement | 8* | For each variable of interest, give sources of data and details of methods of assessment (measurement). Describe comparability of assessment methods if there is more than one group | Pages 7-8 |
| Bias | 9 | Describe any efforts to address potential sources of bias | Pages 8-9 |
| Study size | 10 | Explain how the study size was arrived at | Page 6 |
| Quantitative variables | 11 | Explain how quantitative variables were handled in the analyses. If applicable, describe which groupings were chosen and why | Pages 6-8 |
| Statistical methods | 12 | (*a*) Describe all statistical methods, including those used to control for confounding | Page 8 |
| | | (*b*) Describe any methods used to examine subgroups and interactions | Page 8 |
| | | (*c*) Explain how missing data were addressed | Page 8 |
| | | (*d*) If applicable, describe analytical methods taking account of sampling strategy | N/A |
| | | (*e*) Describe any sensitivity analyses | Page 8 |
| **Results** | | | |
| Participants | 13* | (a) Report numbers of individuals at each stage of study—eg numbers potentially eligible, examined for eligibility, confirmed eligible, included in the study, completing follow-up, and analysed | Page 9 |
| | | (b) Give reasons for non-participation at each stage | Page 9 |
| | | (c) Consider use of a flow diagram | N/A |
| Descriptive data | 14* | (a) Give characteristics of study participants (eg demographic, clinical, social) and information on exposures and potential confounders | Page 9 |
| | | (b) Indicate number of participants with missing data for each variable of interest | Pages 9-12 |
| Outcome data | 15* | Report numbers of outcome events or summary measures | Pages 9-12 |
| Main results | 16 | (*a*) Give unadjusted estimates and, if applicable, confounder-adjusted estimates and their precision (eg, 95% confidence interval). Make clear which confounders were adjusted for and why they were included | Pages 9-12 |

**eTable 13. STROBE statement.**

| | Item No | Recommendation | Location |
|---|---|---|---|
| | | (*b*) Report category boundaries when continuous variables were categorised | N/A |
| | | (*c*) If relevant, consider translating estimates of relative risk into absolute risk for a meaningful time period | N/A |
| Other analyses | 17 | Report other analyses done—eg analyses of subgroups and interactions, and sensitivity analyses | Pages 9-12 |
| **Discussion** | | | |
| Key results | 18 | Summarise key results with reference to study objectives | Pages 12-13 |
| Limitations | 19 | Discuss limitations of the study, taking into account sources of potential bias or imprecision. Discuss both direction and magnitude of any potential bias | Page 14 |
| Interpretation | 20 | Give a cautious overall interpretation of results considering objectives, limitations, multiplicity of analyses, results from similar studies, and other relevant evidence | Pages 12-14 |
| Generalisability | 21 | Discuss the generalisability (external validity) of the study results | Page 14 |
| **Other information** | | | |
| Funding | 22 | Give the source of funding and the role of the funders for the present study and, if applicable, for the original study on which the present article is based | Page 15 |

[*] Give information separately for exposed and unexposed groups.
Abbreviation: N/A, not applicable.

### eTable 14. Institutional review board approval information.

| Institution | Approval number |
|---|---|
| Alfred Health | 305/23 |
| All India Institute of Medical Sciences | IEC-209/11.04.2023, OP-32/16.05.2023 |
| A.C.Camargo Cancer Center | 6.443.601 (CAEE N ° 74036923.5.0000.5432) |
| Aristotle University of Thessaloniki | 75078/2023 |
| ARS Algarve/Coimbra University and Medical School | 36/2023 |
| Center for Medical Education and Clinical Research (CEMIC) | 9256 |
| Centro Hospitalar Vila Nova de Gaia-Espinho | 145/2023 |
| Charité – University Medicine Berlin | EA4/213/22 |
| Chiba University Hospital | HK202305-06 |
| Faculty of Medicine, Chiang Mai University | RAD-2566-09468 |
| Federal University of Rio Grande do Norte | 6.274.288 (CAEE N° 69991023.4.0000.0253) |
| Hospital Universitario Fundación Jiménez Díaz | PIC097-23 |
| Institute of Oncology Ljubljana | ERIDEK-0037/2023 |
| Fundación Universitaria Sanitas - Instituto Global de Excelencia Clínica | CEIFUS 1454-23 |
| KIST Medical College and Teaching Hospital | 2079/80/94 |
| Leiden University Medical Center | 23-3015 |
| Linnaeus University, Umeå University, Lund University | 2023-03578-01 |
| McGill University Health Center | 2023-9537 |
| Mie Chuo Medical Center | MCERB-202301 |
| Muhammadiyah University of Palembang | 113/EC/KBHKI/FK-UMP/V/2023 |
| Ramón y Cajal University Hospital | 139/23 |
| Royal Victorian Eye and Ear Hospital | 23-1580HL |
| Ulster University | 23/0039 |
| Universidad de Las Américas | 2023-EXC-003 |
| Universidade Federal de São Paulo | 6.416.852 (CAEE N° 70812723.0.0000.5505) |
| University Hospital Brno | 13-080323/EK |
| University Hospital Dubrava | 2023/1602-09 |
| Algarve University Hospital Center | UAIF 062/2023 |
| University of Cape Town | 223/2023 |
| Universidad de Costa Rica | CEC-UCR-356-2023 |
| University of Medicine and Pharmacy, Hue University | H2023/022 |
| Fondazione IRCCS Policlinico San Matteo | 0020473/23 |
| University of Salerno | 13/23 |

### eTable 15. Baseline characteristics of the pilot study group.

| | Total (N=100) |
|---|---|
| Gender, no. (%) | - |
| Female | 36 (36%) |
| Male | 64 (64%) |
| Diverse | 0 |
| Not reported | 0 |
| Age | - |
| Median (IQR), y | 59 (40-67) |
| Not reported, no. (%) | 17 (17%) |
| Highest educational level, no. (%) | - |
| Elementary School Diploma | 12 (12%) |
| Middle School Diploma | 30 (30%) |
| High School Diploma | 18 (18%) |
| University Degree | 38 (38%) |
| Not reported | 2 (2%) |
| Which of these technical devices do you use at least once a week? | - |
| Median (IQR), total no. | 2 (2-3) |
| Smartphone, no. (%) | 96 (96%) |
| PC/laptop, no. (%) | 71 (71%) |
| Game console (eg, PlayStation, Switch), no. (%) | 8 (8%) |
| Tablet (eg, iPad), no. (%) | 33 (33%) |
| E-reader, no. (%) | 8 (8%) |
| Smartwatch, no. (%) | 23 (23%) |
| None, no. (%) | 2 (2%) |
| Not reported, no. | 0 |
| What is your current general state of health?, no. (%) | - |
| Very poor | 0 |
| Poor | 13 (13%) |
| Sufficient | 47 (47%) |
| Good | 27 (27%) |
| Very good | 13 (13%) |
| Not reported | 3 (3%) |
| How would you rate your knowledge of artificial intelligence (AI)?, no (%) | - |
| No knowledge (never heard of AI) | 8 (8%) |
| Little knowledge (eg, documentary seen on television) | 64 (64%) |
| Good knowledge (eg, read several articles about AI) | 25 (25%) |
| Expert (eg, involved in AI development) | 2 (2%) |
| Not reported | 1 (1%) |

Abbreviation: IQR, interquartile range.
